# Cancer-associated *Lactobacillus iners* are genetically distinct and associated with chemoradiation resistance in cervical cancer

**DOI:** 10.1101/2022.04.26.22274346

**Authors:** Lauren E. Colbert, Tatiana Karpinets, Molly B. El Alam, Erica J. Lynn, Julie Sammouri, David Lo, Jacob H Elnaggar, Rui Wang, Timothy A Harris, Kyoko Yoshida-Court, Katarina Tomasic, Julianna K. Bronk, Ananta V. Yanamandra, Adilene V. Olvera, Lily G. Carlin, Travis Sims, Andrea Y. Delgado Medrano, Travis Solley, Patricia J. Eifel, Anuja Jhingran, Melissa Joyner, Lilie Lin, Lois M. Ramondetta, Andrew M. Futreal, Kathleen M. Schmeler, Geena Mathew, Stephanie Dorta-Estremera, Jianhua Zhang, Xiaogang Wu, Nadim J. Ajami, Cullen Taniguchi, Joseph F. Petrosino, Jennifer Wargo, K. Jagannadha Sastry, Pablo C. Okhuysen, Ann H. Klopp

**Author notes:** CORRESPONDING AUTHOR Lauren Colbert, Department of Radiation Oncology, 1515 Holcombe Blvd, Houston, Texas 77006.

## Abstract

This study identifies a novel pathotype of cervical cancer-associated *Lactobacillus iners* (*L. iners*) that results in chemoradiation resistance *in vitro* and is associated with poor patient survival. Cervical cancer affects over half a million women a year around the world. Treatment for women with locally advanced cancer is delivered with definitive chemoradiation (CRT) but is curative for only 60% of patients. There are few validated molecular markers to identify patients who will respond poorly to treatment. Tumor microbiome features are associated with treatment resistance in patients with colon and pancreatic cancers, and thus we investigated their role in the response of cervical cancer to therapy. We identified a strong association between poor clinical response to CRT and tumors dominated by *L. iners*. Cancer-associated *L. iners* promoted *in vitro* resistance of cervical cancer cells and modified the local tumor immunologic microenvironment, while non-cancer-associated *L. iners* did not. Assembly of genomes from cancer-derived *L. iners* also demonstrated pathogenic, metabolic, and immune functions not found in healthy patients.

## INTRODUCTION

Cervical cancer remains one of the most common malignancies among women worldwide, with an annual incidence of more than 500,000 cases and an annual death rate of more than 250,000 women per year (Bray *et al*., 2018; Arbyn *et al*., 2020). Despite the successful introduction of HPV vaccination programs, vaccination rates remain low even among targeted populations, and cervical cancer remains a significant global burden. Chemoradiation (CRT) is the standard of care for treating cervical cancer, but 20-40% of patients will experience an incurable relapse (Yuh *et al*., 2009). The molecular factors associated with response to CRT are poorly understood.

The vaginal microbiome is well characterized and generally dominated by members of the *Lactobacillus* genus, which have beneficial roles in the vaginal microenvironment (Verhelst *et al*., 2005; Petrova *et al*., 2017). Beneficial bacteria in the *Lactobacillus* genus are characterized by lactic acid and hydrogen peroxide production, which maintain a healthy pH (Eschenbach *et al*., 1989) and provide other benefits such as immune regulation, maintenance of epithelial integrity, and clearance of other pathogenic viruses and bacteria; However, not all vaginal *Lactobacilli* are beneficial. *Lactobacillus iners (L. iners)*, for example, is a unique species known by microbiologists as a “wolf in sheep’s clothing” due to its tendency to appear in pathogenic or dysbiotic states and share similarities with other non-*Lactobacillus* bacteria, such as Gardnerella. Unlike its usually beneficial Lactobacilli counterparts (Petrova *et al*., 2017), *L. iners* has a small, adaptive, heterogeneous genome and is the smallest genome among known lactobacilli. As opposed to other members of its genus, it often produces limited amounts of lactic acid and little to no hydrogen peroxide (Choi *et al*., 2006; Witkin *et al*., 2013). Curiously, *L. iners* also produces unique inerolysin, a cytotoxic pore-forming toxin (Rampersaud *et al*., 2011).

The cervical microbiome can also play a crucial role in preventing the development and progression of cervical dysplasia and cervical cancer (Chase *et al*., 2015). There is a clear association between the cervical microbiome and clearance of HPV viral infection, although mechanistic studies are lacking Choi *et al*., 2006; Rampersaud *et al*., 2011; Witkin *et al*., 2013; Homburg *et al*., 2017). In contrast to dysplasia, the cervical microbiome in cervical cancer has not been as well defined. This analysis aimed to identify microbial features specific to the cervical cancer microbiome that could serve as potential therapeutic targets in the future.

## RESULTS

A total of 78 patients were enrolled in this prospective, longitudinal study (Fig. 1A) at two institutions. Patient characteristics are summarized in Supplemental Table 1. 82% of tumors were squamous carcinomas, 14% were adenocarcinoma, and 4% were adenosquamous carcinomas. The initial cohort consisted of 43 patients, and an additional 35 patients were later enrolled in a validation cohort. Median follow-up times were 33.2 months for the initial cohort and 21.7 months for the more recently collected validation cohort. 74 cervical microbiome samples were collected from the patients at baseline; 186 serial samples were collected during and after treatment. 146 tumor and 79 peripheral blood samples were also collected at baseline and post-treatment time points, respectively, for immune characterization (supplementary table 2).

**Figure 1.**
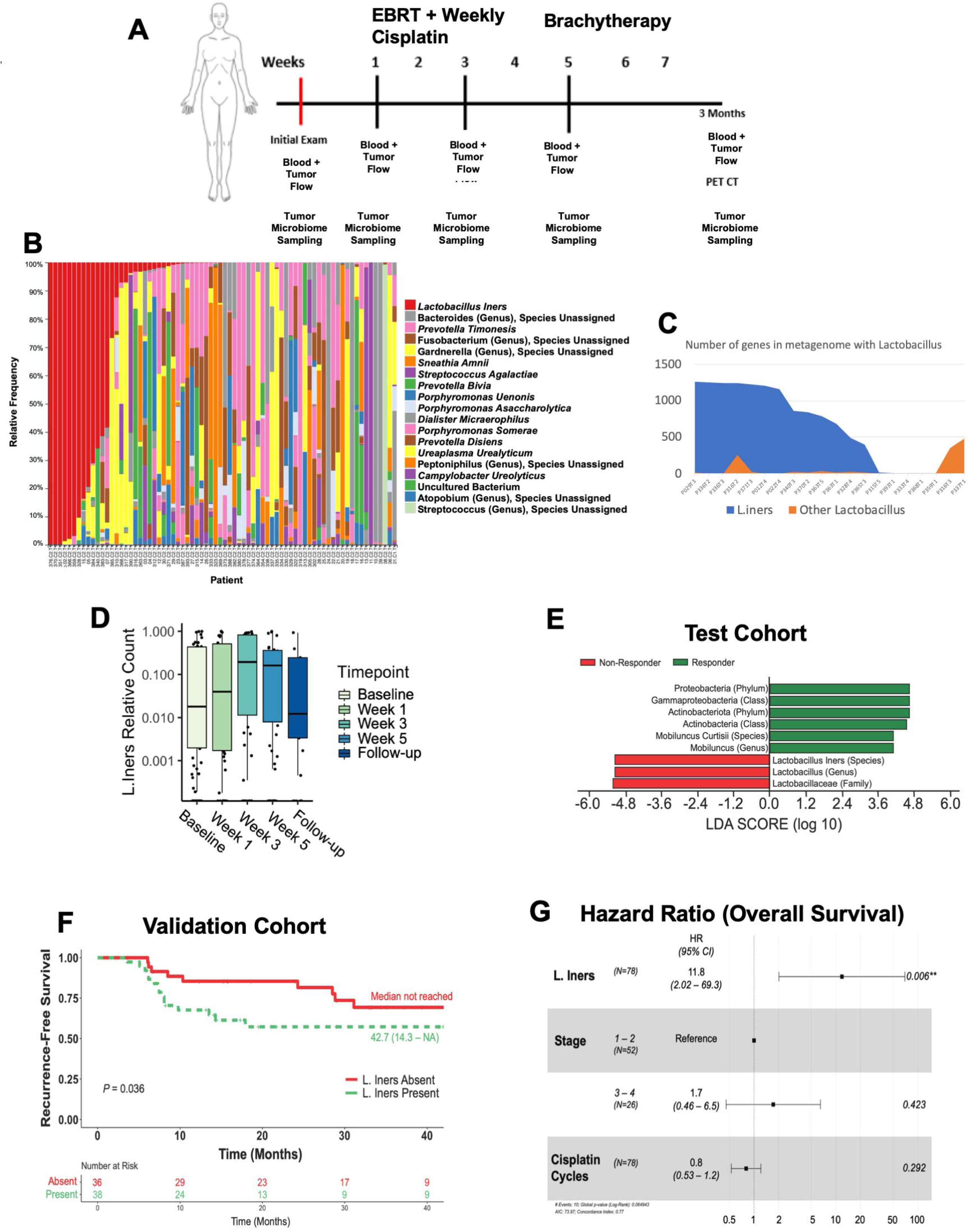
*L. iners* dominates the cervical cancer microbiome and is associated with poor outcomes. A) Study schema of sample collections over time for all patients enrolled on study. B) Stacked bar plot of species-level ASVs from 16s ribosomal RNA sequencing. C) Percent of assembled metagenomes assigned to members of the lactobacillus genus by lactobacillus species from shotgun metagenome sequencing. D) Distribution of relative counts of *L. iners* over time for all patients demonstrating no significant change in relative counts of *L. iners* through CRT. E) LEFSe for differentiating responders and non-responders to radiation in test cohort of 43 patients demonstrates *L. iners* is associated with non-response to radiation. F) Recurrence-free survival in an additional validation cohort of 35 patients at two instututions. G) Hazard ratio for overall survival for entire cohort demonstrates increased *L. iners* relative count is significantly associated with decreased overall survival after adjustment for all significant variables on univariate survival analysis.

### The microbiome of some cervical cancers is dominated by *L. iners* before and throughout treatment

Across all patients, the top 5 most common genera identified from 16s sequencing were Lactobacillus, Prevotella, Bacteroides, Streptococcus, and Gardnerella. On the species level, these were identified as *L. iners, Prevotella timonensis, Prevotella bivia, Streptococcus agalactiae*, and a variety of *Bacteroides* and *Gardnerella* species that could not be further characterized with 16s sequencing (Figure 1B). *L. iners* was present in 48% of patients (38/78) at baseline (Fig. 1B).

To further characterize these organisms, shotgun metagenomic sequencing was performed on baseline samples. Metagenome assemblies were available for 44 patients. Analysis of the genomic composition from samples containing *L. iners* demonstrated that nearly all Lactobacilli-derived genes identified belonged to *L. iners* (Fig. 1C). Thirteen patients had metagenomes with contigs annotated by *L. iners* and the same number of patients had contigs annotated by other *Lactobacillus*, such as *L. gasseri, L. jensenii, L. crispatus* and others. In 8 patients, *L. iners* coexisted with other Lactobacillus species.

To determine if the cervical microbiome within tumors changed during CRT, we evaluated paired, longitudinally collected cervical samples. Individual patient microbiome composition did not shift significantly during CRT (supplemental figure 1). *L. iners* relative counts did not change significantly during CRT (Fig. 1D), nor did richness or diversity (p>0.05 for all). There was a slight shift by week 12 after CRT towards less richness of rare species (Observed features mean 43.9 [SD 24.0] at baseline vs. 63.1 [24.7] at week 12; p=0.02).

### *Lactobacillus iners* is associated with poor treatment response, decreased recurrence-free and overall survival

To identify bacteria associated with treatment response, we performed linear effect size discriminant analysis (LEfSe) in our initial test cohort of 43 patients using 16s sequencing analysis to identify organisms associated with response. We found that a higher number of relative counts of *L. iners* was significantly associated with non-response to CRT (Fig. 1E; LDA score >4) and short-term survival. To validate this finding, we enrolled an additional 35 patients with locally advanced cervical cancer for microbiome analysis. In this validation cohort, the presence of any *L. iners* at baseline was also associated with worse response (residual FDG-avidity post CRT) and significantly lower recurrence-free survival rates (Fig. 1F; p=0.036).

To evaluate interactions with established clinical variables, we performed multivariable analysis on the full cohort. A higher relative count of *L. iners* at baseline was significantly associated with decreased RFS (Table 1; HR 3.71 [95% CI 1.04-13.25]; p=0.04) and overall survival (HR 10.39 [95% CI 1.79-60.25]; p=0.01) in univariate analysis. Stage and cycles of cisplatin received were also associated with RFS (Table 1). On multivariate survival analysis adjusting for clinical factors, increased *L. iners* remained independently associated with decreased RFS (HR 3.95 [1.17-13.25]; p=0.02) and decreased OS (Fig. 1G; HR 11.8 [2.02-69.3]; p=0.006).

**Table 1.** Univariate and Multivariate Survival Analysis. Cox proportional hazards models for recurrence-free and overall survival. Covariates included are the relative counts of *L. iners* before, during, and after chemoradiotherapy and patient clinical and demographic characteristics.

|  |  | Univariate Model for RFS |  | Univariate Model for OS |  | Multivariate Model for OS |  |
| --- | --- | --- | --- | --- | --- | --- | --- |
|  |  | HR (95%CI) | P | HR (95%CI) | P | HR (95%CI) | P |
| **L. Iners** | **Baseline** | 3.71(1.04-13.25) | **0.04** | 10.39(1.79-60.25) | **0.01** | 11.8(2.02-69.41) | 0.006 |
|  | **Week 1** | 2.64(0.64-10.93) | 0.18 | 7.09(0.91-55.57) | 0.06 | — | — |
|  | **Week 3** | 2.96(0.82-10.6) | 0.10 | 7.25(1.07-49.27) | **0.04** | — | — |
|  | **Week 5** | 3.1(0.7-13.81) | 0.14 | 7.97(0.95-66.89) | 0.06 | — | — |
|  | **Follow-up** | 0.52(0.01-32.02) | 0.76 | 17.53(0.18-1727.55) | 0.22 | — | — |
| **Institution** | **Lyndon B. Johnson Hospital** | — | — | — | — | — | — |
|  | **M.D. Anderson Cancer Center** | 1.63(0.69-3.84) | 0.26 | 1.11(0.29-4.31) | 0.88 | — | — |
| **Age** |  | 0.98(0.95-1.01) | 0.17 | 0.99(0.94-1.04) | 0.64 | — | — |
| **Race** | **Asian/Black/Other** | - | - | - | - | — | — |
|  | **Hispanic** | 0.57(0.18-1.80) | 0.34 | 0.47(0.09-2.55) | 0.38 | — | — |
|  | **White** | 0.69(0.22-2.15) | 0.53 | 0.39(0.06-2.41) | 0.31 | — | — |
| **BMI** |  | 0.96(0.9-1.03) | 0.24 | 0.93(0.83-1.05) | 0.24 | — | — |
| **Smoking Status** | **Never** | — | — | — | — | — | — |
|  | **Former** | 1.2(0.55-2.50) | 0.68 | 1.16(0.32-4.14) | 0.82 | — | — |
|  | **Current** | 0.4(0.05-3.00) | 0.37 | 0.00(0.00-NA) | 0.99 | — | — |
| **Histology** | **Adenocarcinoma** | — | — | — | — | — | — |
|  | **Adenosquamous Carcinoma** | 4.74(0.66-34.3) | 0.12 | 3.28(0.27-40.01) | 0.35 | — | — |
|  | **Squamous Cell Carcinoma** | 2.41(0.56-10.27) | 0.23 | 0.85(0.17-4.2) | 0.84 | — | — |
| **LVSI** | **No** | — | — | — | — | — | — |
|  | **Yes** | 1.47(0.37-5.89) | 0.59 | 0.89(0.09-8.66) | 0.92 | — | — |
|  | **Unknown** | 1.08(0.43-2.72) | 0.86 | 0.77(0.19-3.1) | 0.71 | — | — |
| **Stage** | **1 and 2** | — | — | — | — | — | — |
|  | **3 and 4** | 2.93(1.37-6.25) | **0.01** | 1.8(0.5-6.55) | 0.37 | 1.72(0.46-6.49) | 0.423 |
| **Grade** | **Other/Unknown** | — | — | — | — | — | — |
|  | **1** | 0.00(0.00-NA) | 0.99 | 0.00 (0.00-NA) | 0.99 | — | — |
|  | **2** | 0.45(0.17-1.21) | 0.12 | 0.38(0.09-1.72) | 0.21 | — | — |
|  | **3** | 0.75(0.30-1.86) | 0.53 | 0.43(0.09-1.94) | 0.27 | — | — |
| **HPV** | **HPV 16/18** | — | — | — | — | — | — |
|  | **Negative/Other** | 0.94(0.43-2.00) | 0.87 | 2.1(0.49-8.7) | 0.33 | — | — |
| **Cisplatin Cycles** |  | 0.77(0.6-0.98) | **0.03** | 0.82(0.55-1.21) | 0.31 | 0.80(0.53-1.21) | 0.292 |
| **Radiation Dose** |  | 1(1-1) | 0.54 | 1(1-1) | 0.17 | — | — |
| **Antibiotic Use** | **No** | — | — | — | — | — | — |
|  | **Yes** | 1.63(0.76-3.47) | 0.21 | 0.6(0.15-2.38) | 0.47 | — | — |
| **Nodes** | **None** | — | — | — | — | — | — |
|  | **Para-Aortic** | 0.90(0.19-4.20) | 0.89 | 0.80(0.09-7.30) | 0.84 | — | — |
|  | **Pelvic Node** | 1.50(0.63-3.50) | 0.37 | 0.86(2.10-3.60) | 0.84 | — | — |
| **Tumor Dimension** |  | 1.2(0.98-1.47) | 0.08 | 1.16(0.78-1.74) | 0.46 | — | — |

We evaluated relationships between the presence of *L. iners*, and clinical variables and found that the presence of lymphovascular space invasion (LVSI; p=0.03) and HPV18 rather than HPV16 (p=0.01) was associated with the presence of *L. iners* (supplemental table 3). Neither of these factors were significantly associated with RFS or OS on univariate analysis. Antibiotic use during the study period was also evaluated and was not associated with *L. iners* presence. Using the mclust algorithm (Scrucca *et al*., 2016) on the top 25 genera and species in baseline samples showed 3 distinct clusters in each. These clusters were not associated with recurrence-free or overall survival as well as clinical response.

### Cancer-derived *L. iners* acquires additional functions and metabolic capabilities

*L. iners* is a common organism in a healthy cervical microbiome but can easily undergo horizontal gene transfer (Macklaim *et al*., 2011). We hypothesized that *L. iners* in patients with cervical cancer may have undergone similar genomic changes, which may account for its association with poor response to radiation. We performed shotgun metagenomic sequencing on cervical microbiome specimens from 84 patients to evaluate this. A metagenome assembly was produced for 44 patients, of which 13 patients contained at least one *L. iners* contig. All contigs annotated by *L. iners* were combined to represent *L. iners* pan-genome; and contigs that were annotated by other *Lactobacillus*, such as *L. gasseri, L. jensenii, L. crispatus* and others, were combined to represent a cervical cancer Lactobacillus “pan-genome” for comparative analysis. Within the Lactobacillus pangenome, a total of 787 unique KEGG orthologous groups (KOs) were identified for Lactobacillus species using KEGG annotation. Of these, 52% (n=406) were unique to *L. iners, 39%* (n=307) were overlapping between *L. iners* and other/ unidentified *Lactobacillus* species, and only 9% (71) were unique to non-*L. iners* lactobacilli (Fig. 2A). We then compared the cancer-derived *L. iners* pangenome to the complete genomes of *L. iners* KY (Kwak *et al*., 2020), an *L. iners* strain isolated from a healthy individual (Fig. 2B). Interestingly, only 81% (n=593) of KOs aligned between cancer-derived *L. iners* and healthy *L. iners*, and 16% (n=120) of KOs were unique to our cancer-associated strains (Fig. 2B). Only 2% (n=18) KOs were unique to healthy *L. iners*, suggesting the cancer-derived strains acquired additional functions. We also compared our cancer-derived pangenome to assembled contigs of *L. iners* in the cervical metagenome of 14 dysplasia patients. In this comparison, there were an additional 100 KOs (14%) in common between dysplasia-associated *L. iners* that were not identified in healthy *L. iners* (Fig. 2C). This suggests that unique *L. iners* genotypes may be associated with different stages of the cervical dysplasia to cancer spectrum. Annotation of 120 cervical cancer *L. iners* specific KOs using KEGG mapper and Brite ontology revealed an enhanced pathogenic phenotype of *L. iners* from cervical cancer versus a healthy individual. Most of the specific functions were found in several samples (Fig. 2D) revealing common features of the pathogenic phenotype in cervical cancer *L. iners*, such as enhanced CRISPR-Cas system, competence, restriction-modification system, toxin-antitoxin system, antibiotic resistance, transport of lantibiotics, phage-associated proteins, and additional phosphotransferase components. Many of these genes were in common across patients identified in the pan-genome group and unique to genomes assembled from cancer patients (Fig. 2D).

**Figure 2.**
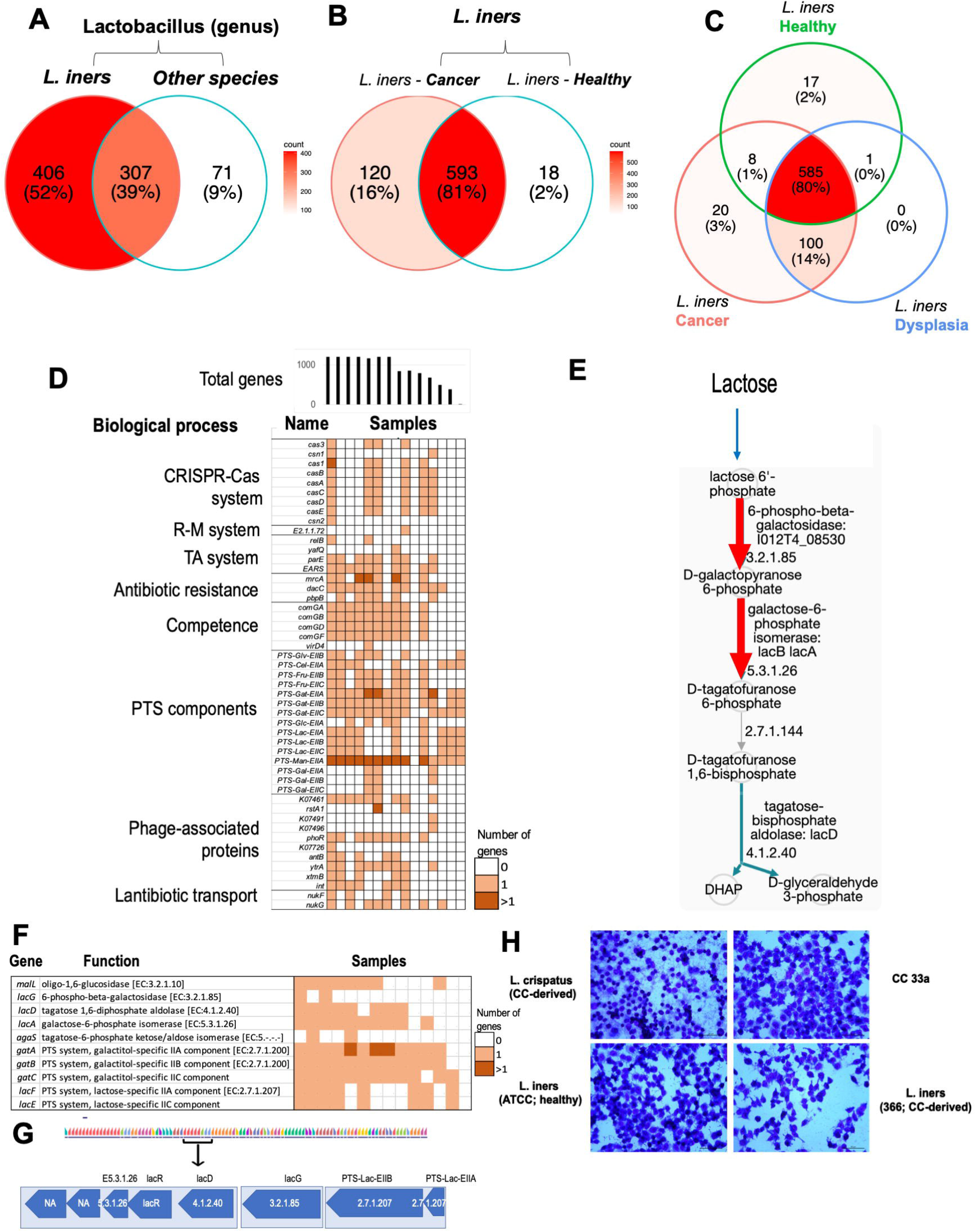
Cancer-derived *L. iners* are distinct from other Lactobacillus and non-cancer derived *L. iners*. A) Total genes assigned to Lactobacillus metagenomes across all patients shows predominantly *L. iners* associated genes. B) For all genes assigned to the Lactobacillus genus, only 9% are potentially assigned to a species other than *L*,. *iners*. C) For all genes assigned to *L. iners*, approximately 20% are found only in cancer-associated *L. iners* and not in *L*. iners associated with healthy controls. C) *L. iners* assemblies from XX patients with dysplasia demonstrates cancer-derived *L. iners* also have more overlap than with healthy *L. iners*. D) Molecular functions specific for cervical cancer *L. iners* are common across different patients and associated with bacterial immunity and pathogenic phenotypes, R-M: Restriction-modification system; PTS: PhosphoTransferase System; TA: Toxin-antitoxin system. E) Pathway of lactose and galactose degradation I predicted in cervical cancer isolated *L. iners* by Pathologic. The pathway is initially characterized in the well-known pathogen *Staphylococcus aureus* that causes skin and soft tissue infections. Enzymes specific for the pathway are labelled by red arrow, Other enzymes are labelled by blue (if enzyme identified) or grey (not identified) in the organism. F) Enzymes involved in the lactose and galactose degradation I and associated PTS components are common in L. iners assemblies produced by WGS. H) Cancer-derived *L. iners* induces cell detachment from the monolayer in c-33a cell line. This does not occur with healthy *L. iners*, cancer-derived *L. crispatus*, or the cell line alone.

Next, we aimed to isolate *L. iners* from cervical swabs of patients with cervical cancer for genomic and functional analysis. *L. iners* was cultured under anaerobic isolation techniques. Two patient-derived strains were isolated and sequenced (Pt1, Pt2; Supplemental table 4). Near-complete assemblies (99.6% and 90.5% completeness) were produced for these strains and then compared with sequenced strains from non-cancer patients: *L. iners* KY (complete genome; healthy patient) and ATCC 55195 (draft genome, 98.7% completeness; bacterial vaginosis patient). Encoded functions (KOs) and biological processes (Brite ontology) were annotated (Supplemental Fig. 2). The comparison of two cancer-derived with ATCC 55195 confirmed similarity of 2 patient-derived *L. iners* with each other and notable differences from ATCC 55195 (Supplemental Fig. 2A). The two cancer-derived strains demonstrated more significant enrichment of functions involved in bacterial immunity and virulence as compared to non-cancer derived strains (Supplemental Fig. 2B).

We further used the Pathway Tools software (Karp *et al*., 2020) to generate PGDBs (Pathway Genome Databases) of the cancer-derived and non-cancer derived *L. iners* strains, in addition to a near complete assembly of a strain derived from a third patient’s WGS data, referred as I012T4 (97% completeness). Results of a comparative analysis of the generated PGDBs is provided in Supplementary tables (Supplemental table 5). Curation of metabolic pathways common and distinct among the strains (Supplemental table 5 the three cancer-derived strains of *L iners* utilize lactose by a distinct lactose and galactose degradation pathway (Supplemental tables 5 versus the healthy individual. The galactose degradation pathway was initially characterized in pathogenic *Staphylococcus aureus* (Bissett and Anderson, 1973, 1980; Button *et al*., 1973) and specifically functions to convert lactose to Galactose-6-phosphate, D-fructose, and in turn to D-glyceraldehyde-3P, upregulating the pentose phosphate pathway (Fig. 2E). Importantly, enzymes of this pathway are found not only in the three cancer-derived strains, but also in most cancer patient samples with *L. iners* from WGS assemblies with a 2.5 fold enrichment versus healthy individuals (Fig. 2F). The pathway was not found in the complete genome of *L. iners* KY isolated from a healthy individual. The enzymes and associated components of phosphotransferase system represent 10 out of 120 cervical cancer specific KOs in th*e L. iners* pan-genome (Fig. 2B) indicating importance of the pathway for survival of *L iners* in the cervical cancer environment. In addition, metabolites produced in the pathway associate (consumed and/or produced) in many reactions predicted for the organism (Supplemental Fig. 3). Mapping of the pathways enzymes to the patient-derived *L. iners* genome revealed that the genes were transcribed from 3 consecutive operons of an assembled contig (Fig. 2G).

Considering the enhanced pathogenic functions described above (Fig. 2D) and known adhesive capability of the bacterium to vaginal epithelial cells (McMillan *et al*., 2013), we examined an ability of cancer-derived *L. iners* to infect the epithelial cells of the cervix (Fig 2H). Compared to the non-infected (NI) controls, cervical cancer-derived *L. crispatus* and non-cancer-derived *L. iners* had little effect on the C-33 A. In contrast, cancer-derived *L. iners* demonstrated increased cytotoxicity to epithelial cells of the cervix by inducing cell detachment from the monolayer.

### *Cancer-derived L. iners* induces treatment resistance in cervical cancer cell lines

We hypothesized that cancer-derived *L. iners* may directly drive resistance to radiation and chemotherapy. We performed cell survival and clonogenic assays in cervical cancer cells co-cultured with supernatants from these various *L. iners* and *L. crispatus* strains. HeLa cells treated with 40% cancer-derived *L. iners* supernatants had increased survival after radiation alone (Fig. 3A; p<0.001) and after combination chemotherapy and radiation treatment (Fig 3B; p<0.001) compared to supernatants from healthy *L. iners*. Clonogenic assays also showed increased survival in the presence of cancer-derived *L. iners* supernatants vs. healthy *L. iners* supernatants (Fig. 3C). Cancer-derived *L. crispatus* strains did not show any difference versus healthy *L. crispatus* (Fig. 3D).

**Fig 3.**
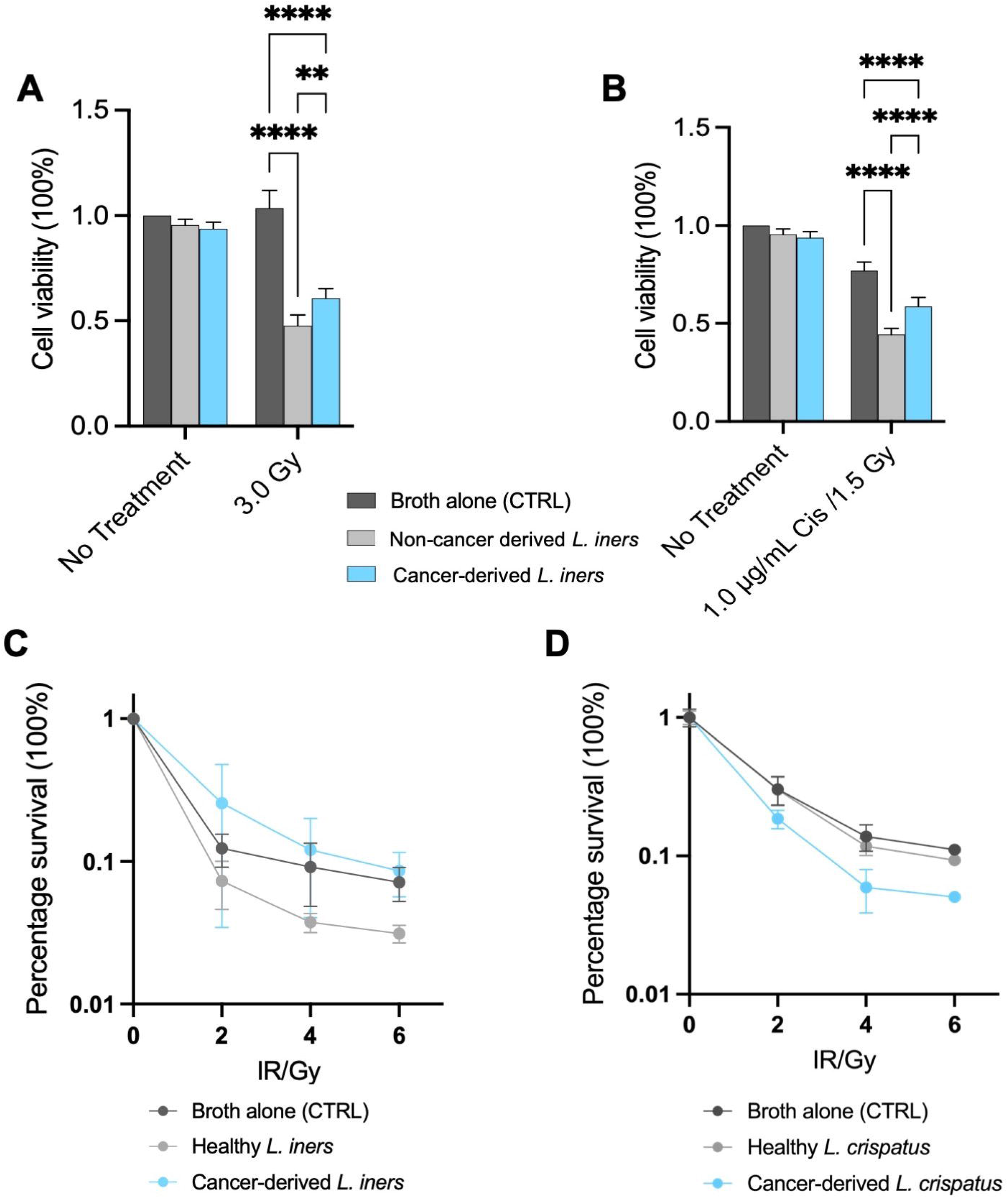
*L. iners* supernatant increases cancer cell survival and chemoradiation resistance. A) HeLa cells treated with 40% NYC broth or 40% *L. iners* supernatants for 6 hours prior to radiation treatment demonstrates increased cell survival for two cancer-derived *L. iners* strains (PD) B) HeLa cells. treated with 40% NYC broth or 40% *L. iners* supernatants for 6 hours prior to chemotherapy and radiation combined treatment also demonstrates increased cell survival for cancer-derived *L. iners* over healthy *L. iners*. C) Clonogenic assays at increasing radiation doses for cancer-derived *L. iners* and healthy *L. iners* show increased radiation resistance for cancer-derived *L. iners* treated HeLA cells. D) Clonogenic assays at increasing radiation doses for cancer-derived *L. crispatus*, healthy *L. crispatus* and negative controls show increased radiation sensitivity for HeLa cells treated with *L. crispatus*.

### *L. iners* is associated with dysfunctional CD8+ anti-tumor immune responses

We also hypothesized that *L. iners* might alter the immunologic tumor microenvironment. To evaluate this, we performed flow cytometry for activation, proliferation, and exhaustion T-cell markers at each time point before, during, and after chemoradiation as used previously with similar findings across timepoints (Fig. 4A) (Dorta-Estremera *et al*., 2018; Colbert *et al*., 2022). Dimension reduction using the Mclust (Scrucca *et al*., 2016) algorithm identified seven distinct clusters of CD8+ T cell subsets (Fig. 4B), including a functional cluster 3 with low exhaustion (low CTLA4+ low PD1+ high GranzymeB), a functional cluster 6 with exhaustion (high granzyme B and high CTLA4+ and high PD1+), and a low-functional cluster 7 (low CTLA4+ and low PD1+ but low Granzyme B). Patients without *L. iners* at baseline were more likely to have a functional CD8+ population with low levels of exhaustion in the tumor microenvironment at the end of treatment (Fig. 4C; p<0.001), while patients with high *L. iners* at baseline were more likely to have either a functional and exhausted phenotype or a non-functional phenotype at the end of treatment (both p<0.001).

**Fig 4.**
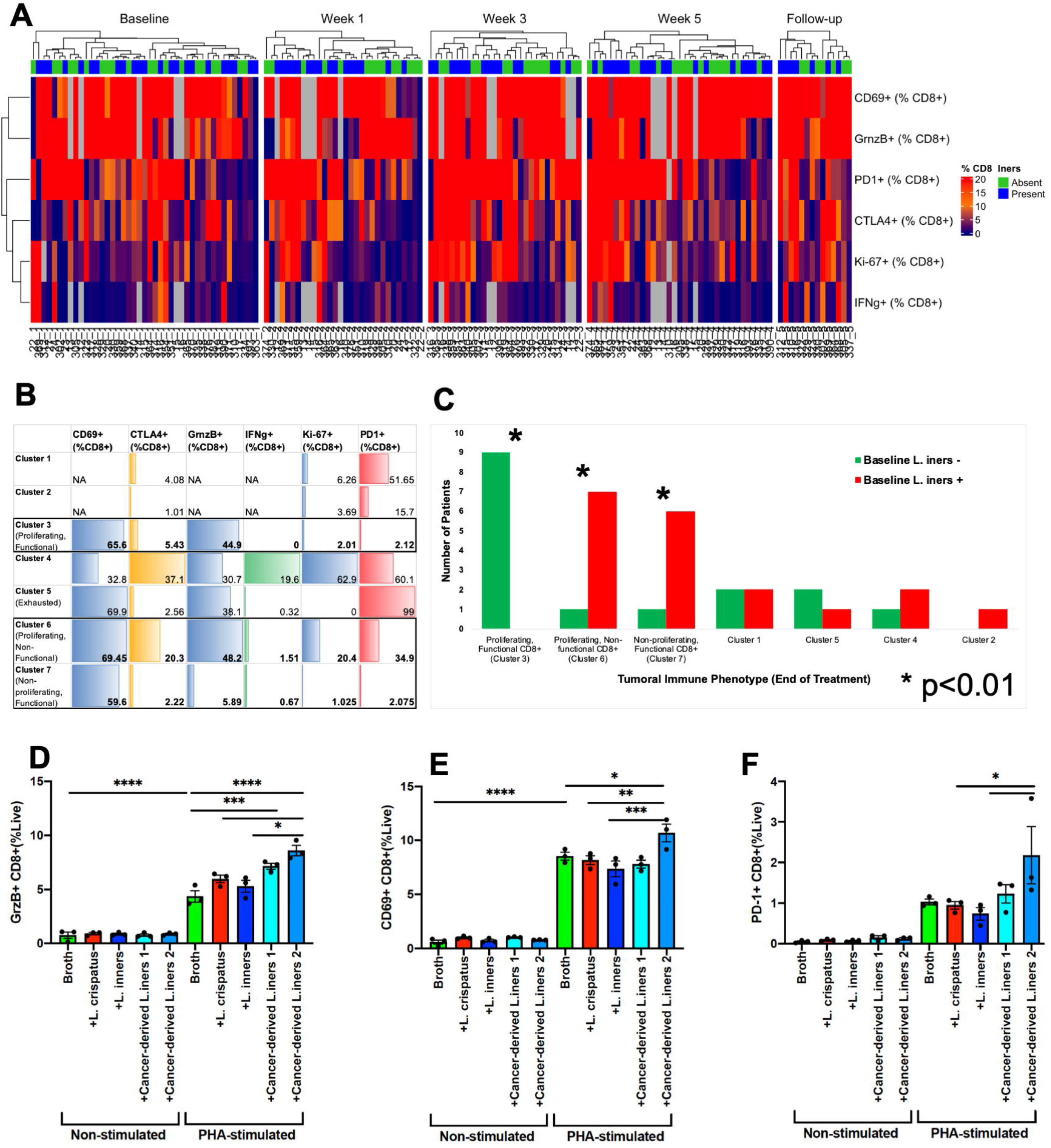
*L. iners* is associated with dysfunctional CD8+ anti-tumor immune responses. A) Clustering of CD8+ subsets by timepoint. B) Dimension reduction using Mclust algorithm identified 7 distinct clusters of CD8+ subsets, included a proliferating, functional cluster 3 (high CD69+ low CTLA4+ low PD1+ high GranzymeB), a proliferating, but non-functional cluster 6 (high granzyme B and high CD69+ but also high CTLA4+ and high PD1+), and a non-proliferating but functional cluster 7 (high CD69+, low CTLA4+ and low PD1+ but low Granzyme B). C) Patients without *L. iners* at baseline were more likely to have a proliferating, functional CD8+ population in the tumor microenvironment at the end of treatment (p<0.001), while patients with high *L. iners* at baseline were more likely to have either a non-proliferating, functional phenotype or a proliferating, non-functional phenotype at end of treatment (both p<0.001). In co-culture experiments with various Lactobacillus species and patient-derived PBMCs, although CD8+ GranzymeB (D) and CD69+ expression((E) is higher with PHA stimulation for cancer-derived *L. iners* strains over *L. crispatus*, non-cancer derived *L. iners* or media alone, (F) PD-1 expression is also significantly higher. IFNg and Ki67 expression is not different.

To evaluate the impact of *L. iners* on immune phenotype in-vitro, we performed co-culture experiments with various Lactobacillus species and peripheral blood mononuclear cells PBMCs (Fig. 4D-F). We found that CD8+ T cells with enhanced GranzymeB and PD-1 expression following incubation with cancer-derived L. *iners* strains compared to non-cancer-derived *L. iners, L. crispatus*, or media alone. IFNg and Ki67 expression were not significantly different (Supplemental Fig. 4). These in-vitro results suggest that *L. iners* from patients with cancer alter the tumor microenvironment and likely impact tumor-specific immune responses.

## DISCUSSION

This study has identified a novel pathotype of cervical cancer-associated *Lactobacillus iners* that results in chemoradiation resistance *in vitro* and is associated with poor patient survival. The metagenomic analysis identified additional common functions involved in immunity, pathogenicity and metabolism in the genomes of *L. iners* isolated from patients with cancer, which were not present in *L. iners* from patients without cancer. Many of these genes were also identified in patients with dysplasia, suggesting that the presence of these genes may favor malignant progression. We observed in-vitro effects of cancer-derived *L. iners* that were not found with *L. iners* from healthy patients, including detachment of epithelial cells, tumor cell resistance to radiation and chemotherapy, as well as altered immune cell response to PHA stimulation. We hypothesize that the phenotypic differences seen *in vitro* and in patients are driven by pathogenic, metabolic and immune functions that may metabolically benefit cancer cells and outcompete healthy Lactobacillus species. Metabolic functions may directly upregulate of the Warburg effect and also cause dysfunctional CD8+ T-cell responses.

In healthy women, beneficial *Lactobacillus* species secrete various metabolites, such as exopolysaccharides (EPSs), phosphorylated polysaccharides, and peptidoglycans that promote vaginal health. In the complicated microenvironment of cancer cells, however, these metabolites can also inhibit or accelerate the proliferation of malignant tumors (Zadravec, Štrukelj and Berlec, 2015; Homburg *et al*., 2017). In fact, one previous study showed that ATCC *L. iners* (used as our non-cancer control) actually inhibited the proliferation and migration of cervical cancer cells by activating the WNT pathway in epidermal cells (Fan *et al*., 2021). In HPV-related cervical cancer, the presence of *Lactobacillus* spp has been shown to widely vary with a typically reduced prevalence of *L. crispatus* and an *L*.*iners* dominant microenvironment among most women (Yang *et al*., 2018). Consistent with this observation, we observed a chemotherapy and radiation resistance from tumor cells incubated with cancer-derived *L. iners* supernatants that were not seen with healthy derived *L. iners*, providing additional evidence that our cancer-associated *L. iners* are genetically and functionally distinct from beneficial Lactobacilli or healthy *L. iners*.

One potential mechanism by which our cancer-derived strain of *L. iners* may drive poor response to CRT is through altered tumor metabolism. We hypothesize that this is via upregulation of various steps in the galactose metabolism pathway, specifically the conversion of lactose to Galactose-6-phosphate, D-fructose, and in turn to D-glyceraldehyde-3P. This results in increased G6PD and fructose in the tumor environment, leading to preferential use of aerobic respiration. This results in downregulation of mitochondrial respiration and an increase in aerobic respiration consistent with the Warburg effect, which results in radiation resistance (Vaupel, Schmidberger and Mayer, 2019).

Another potential pathway through which these cancer-associated *L. iners* may drive poor outcomes is by inducing a dysfunctional anti-cancer immune response in the setting of cancer-associated *L. iners*. Lactobacillus have well-described immunomodulatory features, although generally in the context of beneficial Lactobacilli such as *L. crispatus* and *L. reuteri*. These lactobacilli protect mucosal surfaces against opportunistic or pathogenic infections and are used as immune response boosting probiotics (Lebeer, Vanderleyden and De Keersmaecker, 2008; Artym and Zimecki, 2021). We hypothesize that it may not be the presence of cancer-associated *L. iners* that directly results in a dysfunctional immune response, but rather that its dominance outcompetes these more beneficial Lactobacillus species.

In clinical practice, modification of the vaginal microbiome is feasible and relatively low risk. For example, in the setting of bacterial vaginosis (BV), another disease defined by overgrowth of a particular vaginal microbe which competes with beneficial Lactobacilli, the application of topical metronidazole followed by a constitution of a beneficial strain of *L. crispatus*, known as LACTIN-V, resulted in the presence of *L. crispatus* in nearly 80% of patients 12 weeks after initial treatment with relatively few side effects and decreased the risk of BV recurrence by 1/3 (Cohen *et al*., 2020). Other potential focused interventions to eliminate *L. iners* such as topical application of specific bacteriocins, lytic phage use, or probiotics have successfully been used to treat recurrent *clostridium difficile* and methicillin-resistant Staphylococcus aureus (MRSA) infections (Nataraj and Mallappa, 2021; Ariyoshi *et al*., 2022).

Although this study generates new hypotheses, several significant limitations are worth noting. First, our *L. iners* collection is small, and it is difficult to ascertain whether additional strains isolated from other patients might yield different results. Additionally, co-cultures of cancer cells and bacteria or bacterial supernatants can cause additional effects on cells that are difficult to interpret, such as pH or nutrient alterations. We acknowledge a limited number of patient-derived strains and *L. iners* assemblies but posit these initial findings are worth further investigation.

In conclusion, our work identified a novel relationship between the tumor microbiome in cervical cancer and poor survival. *L. iners* dominates the microbiome in this subset of high-risk patients, which accounts for a significant percentage of relapses. Cancer-associated *L. iners* appears to have acquired multiple environment regulating genes that result in a tumor-promoting phenotype. Future studies will be focused on examining the critical role of these genes and pathways and investigating ways to target this population for microbiome intervention.

## Supporting information

Key Resources Table

## Data Availability

All data produced in the present study are available upon reasonable request to the authors and will be publicly available when the manuscript has been accepted after peer review,.

## Acknowledgments

This study was presented in part at the American Society for Therapeutic Radiation Oncology (ASTRO) Annual Meeting, September 24th-27th, 2017 and at the American Society of Clinical Oncology Clinical Immuno-Oncology Symposium (ASCO-SITC), January 25th-27th, 2018. This study was partially funded by The University of Texas MD Anderson Cancer Center HPV-related Cancers Moonshot and a Resident/Fellow grant from Radiological Sciences North America (RSNA). This shared resource is partially funded by NCI Cancer Center Support Grant P30CA16672.

## Author contributions

Conceptualization: A.H.K, L.E.C, N.A.

Methodology: A.H.K., L.E.C., T.K., M.B.E, X.W, T.S., P.J.E, A.M.F, J.K.S, S.D.E, P.C.O, N.A, J.Z.

Investigation: L.E.C, E.J.L, D.L, T.A.H, A.Y.DM., T.S., G.M., N.J.A

Writing – Original Draft: A.H.K., L.E.C., T.K., M.E.B, E.J.L

Writing – Review & Editing: All authors contributed

Funding Acquisition: A.H.K., L.E.C

Supervision: J.K.S., S.D.E., C.T., P.C.O., J.F.P., P.J.E., A.J., A.M.F

## Star Methods

### Resource Availability

#### Lead contact

Further information and requests for resources and reagents should be directed to and will be fulfilled by the Lead Contact, Lauren Colbert

#### Materials availability

Patient-derived Lactobacillus strains are grown at MD Anderson and available for sharing per request. The sequences of these strains have been uploaded to NCBI.

#### Data and code availability

1. Data: 16S and WGS raw data will be deposited in SRA and will be made publicly available upon acceptance of the manuscript. *L*.*iners* strain assemblies using WGS have been deposited in NCBI’s Genome and will be made publicly available upon acceptance of the manuscript (BioSample accession numbers: SAMN27176861, SAMN27176862, SAMN27176863, SAMN27176864)
2. Code: All original code is available in this paper’s supplemental information.
3. Any additional information required to reanalyze the data reported in this paper is available from the lead contact upon request

#### Experimental Models and Subject Details

##### Chemoradiation Cervical Cancer Patients

55 patients were enrolled from The University of Texas MD Anderson Cancer Center and 23 from Harris Health System, Lyndon B. Johnson General Hospital Oncology Clinic (Supplemental table 1). 82% of tumors were squamous carcinomas, 14% were adenocarcinoma, and 4% were adenosquamous carcinomas. The majority of tumors were FIGO stage IIB (37%). Patients received a minimum of 45Gy of external beam radiation therapy (EBRT) in 25 fractions over 5 weeks with weekly cisplatin, followed by two brachytherapy treatments at approximately week 5 and week 7 with EBRT between brachytherapy treatments for gross nodal disease or persistent disease in the parametrium. Stage IB1 patients were treated with CRT due to nodal disease.

##### Clinical Efficacy Analysis

Response to radiation was monitored using clinical exam through the course of treatment, week 5 MRI and surveillance PET/CT at 3 months. Poor responders to radiation were those patients with less than complete response during CRT and residual FDG-avidity at 3 month follow up exam. Short-term survivors were categorized as those that had a recurrence or cancer-related death within 24 months of diagnosis and long-term survivors were those who had reached 36 months follow-up with no recurrence or cancer-related death. Patients with incomplete follow up data were excluded from this analysis. For survival analysis, any biopsy-proven recurrence on physical exam or follow up imaging was coded as a recurrence event. Last known follow up was last clinic visit with exam and/or imaging. For overall survival, any contact with patient was used as last follow up with alive/ dead status. Clinical response was also categorized as poor, standard, and excellent according to the tumor response on week 5 MRI (prior to intracavitary radiation treatment) and surveillance PET/CT at 3 months. The clinical response was considered poor if residual tumor was found on week 5 MRI. Standard response corresponded to the absence of tumor on week 5 MRI with residual tumor on surveillance PET/CT. The response was considered excellent when no tumor was found on week 5 MRI and PET/CT at 3 months.

##### Cell Lines

C33-A cells, an epithelial cell line from the cervical tumor of a female patient, were obtained from the ATCC. Cells were cultured in 1X MEM with 10% FBS at 37°C and 5% CO_2_. These cells have not been authenticated.

HeLa cells, an epithelial cell line from the cervical tumor of a female patient, were a generous gift of the Sam Mok lab. Cells were cultured in 1X MEM with 10% FBS and 1% Penicillin/Streptomycin at 37°C and 5% CO_2_. These cells have not been authenticated.

##### Bacterial Strains

*Lactobacillus crispatus* strains (ATCC and Pt-3) were cultured in MRS broth at 37°C in anaerobic conditions (10% CO_2_, 5% H_2_, nitrogen balance).

*Lactobacillus iners* strains (ATCC, Pt-1, Pt-2) were cultured in NYC III broth at 37°C in anaerobic conditions (10% CO_2_, 5% H_2_, nitrogen balance).

#### Method Details

##### Sample Collection

Physicians collected tumor swabs of the cervix at five time points: baseline, week 1 (after 5 radiotherapy fractions), week 3, week 5, and first follow-up (12 weeks post-treatment) for 16S rRNA sequencing. For flow cytometry analysis, tumor cytobrush and blood samples (10-20 mL collected in EDTA-containing vacutainers) were also collected at the same timepoints.

Tumor swabs for microbiome sequencing were collected with an Isohelix Buccal Swab (Isohelix, DSK-50) by swabbing the tumor and immediate region. Swabs were placed into individual collection tubes and transported at room temperature to the lab within 4 hrs. In the lab, 400 μLs of stabilization buffer (Isohelix, BFX-25) were added to each tube, which were vortexed for 15 secs and stored at -80°C until DNA extraction.

Two BBL CultureSwabs (BD Biosciences, Cat# 220135) were swabbed in the tumor region by a physician and transported to the lab within 30 minutes for downstream culturing to isolate patient *Lactobacillus* strains.

Two Cytobrush Plus Endocervical Samplers (Cooper Surgical, C0012) were rotated against the tumor and immediate region. Brushes were placed into individual conical tubes and immediately transported at room temperature to the lab. In the lab, 10 mLs of sterile complete RPMI-1640 media, containing 1% penicillin-streptomycin and gentamicin antibiotics (Fisher Scientific, SH30027FS, SV30010, and BW17-518Z, respectively) and 10% fetal bovine serum (Mediatech, MT35010CV), were added to each tube, which were then vortexed for 1 min to dislodge and suspend cells. When large amounts of mucus were present, 5 mLs of dithiothreitol solution (1X Hank’s balanced salt solution, 4% bovine serum albumin, 2 mM dithiothreitol; Invitrogen, P2325) were added to the cell suspensions and passed through a 70-μm cell strainer into new conical tubes. Cells were pelleted by centrifugation and resuspended in sterile complete RPMI-1640 media to be immediately used for flow cytometry.

For blood samples, a total of 10-20 mLs of blood were collected into 10-mL EDTA-containing vacutainers (BD Biosciences, 366643) and were transported at room temperature to the lab within 4 hrs. In the lab, sterile PBS was added to the blood in the vacutainers at a 1:1 ratio. The diluted blood was then layered in new conical tubes at a 2:1 ratio onto Lymphoprep (Stemcell Technologies, 07851; or Cosmo Bio Usa, AXS1114545) and centrifuged at 400 x g, with the brakes off, for 40 mins at room temperature. The peripheral blood mononuclear cells (PBMC) were rinsed once in sterile PBS and twice in sterile complete RPMI-1640 media. were immediately used for flow cytometry.

##### Isolation of L. Iners strains from Patient Tumor Swab Collections

Within 30 minutes of collection, tumor swabs were plated onto a TSA plate (BD Biosciences, Cat# 221239) and a MRS plate (Moltox, Cat# 51-40S020.140) and incubated at 37 degrees in anaerobic conditions for 3-5 days. Bacterial growth from plates was sub-cultured until single colonies could be isolated. MALDI-TOF mass spectroscopy was used to identify the bacterial isolate species.

##### 16S rRNA Sequencing

16S rRNA gene sequencing was performed by personnel of the Alkek Center for Metagenomics and Microbiome Research at Baylor College of Medicine. 16S rRNA was sequenced using methods adapted from the methods used for the Human Microbiome Project11,12. Briefly, bacterial genomic DNA was extracted using MO BIO PowerSoil DNA Isolation Kit (MO BIO Laboratories). The 16S rDNA V4 region was amplified by PCR and sequenced on the MiSeq platform (Illumina) using the 2×250 bp paired-end protocol, yielding pair-end reads that overlapped almost completely. The primers used for amplification contain adapters for MiSeq sequencing and single-end barcodes allowing pooling and direct sequencing of PCR products12

The 16S rRNA gene data incorporate phylogenetic-based and alignment-based approaches to maximize data resolution. Sequence read pairs were demultiplexed using unique molecular barcodes, and reads were merged using USEARCH version 7.0.1090. 16S analysis was performed using custom analytic packages and pipelines developed at the Alkek Center for Metagenomics and Microbiome Research at Baylor University to create summary statistics and quality control measurements for each sequencing run, as well as multi-run reports and data-merging capabilities for validating built-in controls and characterizing microbial communities across large numbers of samples or sample groups.

We processed the 16S rRNA sequence reads using the QIIME2 microbiome bioinformatics platform (version 2020.11) (Bolyen *et al*., 2019). We did not set a sampling depth and so the data was not rarefied for the analysis presented in this study. We demultiplexed the sequence data using the Earth Microbiome Project (EMP) protocol (Hamady *et al*., 2008; Hamady and Knight, 2009). We then constructed the Amplicon Sequence Variant (ASV) feature table using DADA2 denoising (Nearing *et al*., 2018). Phylogenetic reference construction was performed using a pre-trained Naïve Bayes classifier and the q2-feature-classifier plugin. The taxonomic classifier used was trained on the SILVA 138 515F/806R region of sequences which have been trimmed to include 250 bases from the 16S V4 region (Pruesse *et al*., 2007). Alpha diversity and evenness metrics were calculated through QIIME2. The indices used in this study are Shannon Diversity Index (SDI), Simpson Diversity, Faith’s Phylogenetic Diversity (PD), Fisher’s index, Pielou’s evenness, Simpson’s evenness, and Observed ASV’s (Observed Features).

##### Shotgun Metagenomic Sequencing

Shotgun metagenomic sequencing was performed by personnel of the Alkek Center for Metagenomics and Microbiome Research at Baylor College of Medicine.

DNA was isolated from Lactobacillus strains with Gentra Puregene Yeast/Bact. Kit (Qiagen, Cat.# 158567) following manufacturer’s instructions. After 16Sv4 sequencing, DNA isolates from tumor swabs were sequenced with Illumina sequencers.

##### Cell Adherence Assay

C-33A cells were seeded into chamber slides with 5×10^5^ cells per well. The following day each bacteria strain was added to the cultures to infect the cells with a 1:1 MOI and incubated for 3 hours. Slides were washed 5 times in PBS, fixed with 100% methanol, and stained with Wright-Giesma.

##### *Lactobacillus* Supernatant Treatments

Bacterial strains were cultured under anaerobic conditions, temperature at 37°C, shaker 190 rpm for 3-4 days until cells reached a density of approximately 1 × 10^9^ cells/mL. Cultures were centrifuged at 4000 rpm for 40 minutes at 20°C. Supernatants were sterile filtered [Corning Incorporated, NY 14831]. HeLa cells were seeded into 96-well plates at 100 cells per well and left to adhere overnight. Supernatants from the bacterial culture or the corresponding bacterial culture broth were applied to cells at 40% v/v bacteria broth to complete MEM culture medium and incubated at 37°C and 5% CO_2_ for 6 hours. after the incubation of supernatants, the medium was replaced, and cells were then treated with or without cisplatin, ionizing radiation or both as indicated in the figure and figure legends. After all treatments, cells were allowed to grow for 4-6 days until the negative control wells reached roughly 75% confluency before cell viability was assessed. The cell viability assay was performed with Cell Titer Glo per manufacturer’s instructions. Cell culture plates were allowed to come to room temperature, then the Cell Titer Glo Reagent was applied to culture medium at a 1:1 ratio and incubated for 2 minutes. Lysed cells and well contents were transferred to a clear-bottom opaque 96-well plate and incubated at room temperature for 10 minutes. Luminescence was measured with a Perkin Elmer Victor X3 plate reader for 1 second per well. Percent cell viability for each treatment group was calculated by normalizing luminescence readings to that of the broth negative control group.

##### Flow Cytometry

Immunostaining of lymphocytes was performed according to standard protocols. Cells were fixed using the Foxp3/Transcription Factor Staining Buffer Set (eBioscience, Cat# 00-5523-00) and stained with a 16 color panel with antibodies from Biolegend, BD Bioscience, eBioscience, and Life Technologies. Analysis was performed on a 5-laser, 18 color LSRFortessa X-20 Flow Cytometer (BD Biosciences). Analysis was performed on Flowjo Software (INFO).

##### Co-Culture of *Lactobacillus* with patient-derived PBMCs

1 × 10^6^ human patient-derived PBMCs were co-cultured with either 5 mg/mL phytohemagglutinin (PHA), live bacteria at a 1:1 ratio of bacteria cells to PBMCs, both PHA and bacteria, or in media as a negative control. Plates were incubated at 37°C and 5% CO_2_ for 16 hours total. After 12 hours, Golgiplug (containing Brefeldin A, BD Biosciences, Cat.# 555029) was added to each of the wells and then returned to the incubator for an additional 4 hours. PBMCs were stained with surface antibodies, fixed, and then stained with intracellular antibodies. Cells were stored at 4°C until being acquired by flow cytometry.

#### Quantification and Statistical Analysis

##### Microbiome Sequence Processing

Postprocessing of shotgun metagenome sequencing data was implemented using a set of software tools summarized in the key resources table. The paired-end raw sequence reads in fastq format were filtered and trimmed using BBMap [https://jgi.doe.gov/data-and-tools/bbtools]. To remove contamination by human DNA, which was abundant in the cervical swabs, the Bowtie 2(Langmead and Salzberg, 2012) was used to align filtered and trimmed sequencing reads to a hg38 reference genome (GCA_000001405.28). After removing the contaminated human reads, the remaining reads were assembled into contigs by both MEGAHIT [ doi: 10.1093/bioinformatics/btv033] and metaSPAdes(Nurk *et al*., 2017). Only contigs that were larger than 1000 bp were used for binning by MetaBAT2(Kang *et al*., 2019). Genes were predicted in each assembled contigs by Prodigal (Hyatt *et al*., 2010) and then annotated by KofamKOALA (Aramaki *et al*., 2020), which assign KOs ID, gene name, function, and EC number. Taxonomic classification of each contig was implemented by CAT and BAT (von Meijenfeldt *et al*., 2019). All the software tools were running with default parameters. Versions and sources of the software tools or packages used in the pipeline are listed Table S1. Output of the assembly was a set of assembled contigs for each sample, their taxonomic annotation, predicted genes, their location in the contig, strand, size, KO IDs, genes names, gene functions (description), and EC numbers. Some genes were annotated by 2 or more KOs or/and EC numbers.

##### Statistical Analysis

All analyses presented were performed on the initial cohort, the validation cohort, and the full cohort independently. We performed strict evaluation for batch effect, including sensitivity analyses for endpoints of interest by batch for each timepoint, institution and by patient demographics. We also analyzed batches separately by institution with no differences in outcome. We also noted that in, particular, batch 4 had higher number of features compared to the other batches (Supplemental Fig 5) and so we performed sensitivity analyses with and without it to ensure it did not affect our results.

We studied the changes in alpha diversity metrics, and relative counts of Lactobacillus and *L. Iners* over time using a paired T-test. Linear discriminant analysis effect size (LEfSe) was used to identify taxa that were enriched in baseline samples with the clinical response set as “class”(Segata *et al*., 2011). A linear discriminant analysis (LDA) score of 4.0 was used with a 0.05 alpha value for the Kruskal-Wallis all-against-all test.

Univariate Cox proportional hazards models were built for overall survival and recurrence free survival. The models included the alpha diversity metrics as well as the relative counts of *L. iners* and Lactobacillus at each of the study timepoints. We also included clinical and demographic characteristics. Co-variates that were found to be significant at the univariate level (p<0.2) were then all fitted in a multivariate Cox proportional hazard model. Unsupervised hierarchical clustering of the top 25 species and genera in baseline samples was conducted using the mclust machine learning algorithm in the full cohort (Scrucca *et al*., 2016). We then ran survival analysis on the cluster groups by fitting Cox proportional hazards models for overall and recurrence free survival. We also tested associations between the cluster groups and clinical response using a Pearson’s chi squared test.

Postprocessing of the obtained fastq files was implemented by the same computational workflow used for postprocessing of shotgun metagenome sequencing data. The workflow, which is described above, generated assembled contigs for each strain that were further curated to avoid duplications. The assembled contigs for each strain were annotated using a prokaryotic genome annotation pipeline DFAST (Eriksson *et al*., 2021). The software implements gene predictions for protein coding sequences, rRNA, tRNA, and CRISPR, and infers protein functions. Completeness check was calculated by DFAST at the genus level (Lactobacillus, 14 genome, 238 marker sets) and revealed completeness values 98.7% for ATCC 55195 strain and 99.6% and 90.5% for Pt1 and Pt2 respectively. The level of contamination calculated by DFAST was 0.6-0.67%.

Identity check evaluated taxonomy identity by calculating average nucleotide identity and by comparison the value with 13000 reference genomes using FastANI software (Jain *et al*., 2018), HMM scan against TIGRFAM and RPSBLAST against COG were enabled as advanced options of the annotation. Gene predictions were done using Prodigal (Hyatt *et al*., 2010) as an option provided by DFAST. Characteristics of the assemblies are provided in Supplemental table 4. The PathoLogic component (Karp, Latendresse and Caspi, 2011) of the Pathway Tools software 25.5 and MetaCyc v.25.5 (*MetaCyc database of metabolic pathways and enzymes* | *Nucleic Acids Research* | *Oxford Academic*) were used to generate PGDBs for draft genomes of 2 L. iners strains (pt1 and pt2) isolated from cervical swabs of 2 CC patients, ATCC 55195 strain isolated form a patient with bacterial vaginosis, I012T4 strain (draft genome assembled from WGS data of CC patient 12) and for the complete genome of L.iners isolated from a healthy individual. The default parameters were used if not specified to run the tools. The Pathway tools were also used to predict transcription units, generate contigs overviews, pathway diagrams, and to compare the generated PGDBs. Characteristics of the generated PGDBs are provided in Supplemental table 5.

We then studied how the clinical and demographic characteristics might affect the diversity and evenness indices as well as the relative counts of Lactobacillus and *L. Iners* in the cervical microenvironment by fitting univariate linear regression models. We also used the model-based hierarchical clustering algorithm mclust on different subsets of flow cytometry data from blood and tumor samples. The subsets used were CD4 (% live lymphocytes), CD8 (% live lymphocytes), CD4 subsets, and CD8 subsets. We set the classifier as the baseline presence or absence of *L. Iners* in each patient. We then tested for differences in the median relative counts of *L. Iners* at baseline between the clusters at each time point in both sample types by a Kruskal-Wallis test. We tested for associations between the clusters and patients’ baseline *L. Iners* status using an exact Pearson’s chi squared test. Statistical significance was set at an a of 5% for a two-sided P value. All available samples were used for analyses. Analyses were conducted using RStudio 2021.09.1Ghost Orchid (2020).

## Supplemental Information

**Supplemental Table 1. Patient Characteristics**. Patients’ clinical and demographic characteristics (N=78)

**Supplemental Table 2. Availability of samples**. List of all available samples at each timepoint for 16S and flow in tumor and blood samples.

**Supplemental Table 3. Association of L. iners with patient characteristics**. Univariate linear regression models of *L. iners* with patients’ clinical and demographic characteristics

**Supplemental Table 4. Characteristics of draft genomes of the strains used for further annotation and analysis**. L.iners ATCC 55195 is a draft genome of L.iners ATCC 55195 strain isolated form a patient with bacterial vaginosis. L.iners I012T4 is the high quality draft genome of L.iners assembled from WGS sequencing data of a cervical swab of CC patient.

L.iners KY is the complete genome of L.iners isolated from a healthy individual. L.iners pt1 and L.iners pt2 are draft genomes of L.iners isolated from cervical swabs of 2 CC patients.

The table was generated by Pathway Tools software.

**Supplemental table 5. Comparative Analysis Summary Results**

Supplemental table 5A. Database Summary Statistics

Supplemental table 5B. Breakdown of Reactions by Type

Supplemental table 5C. Reactions of Small Molecule Metabolism

Supplemental table 5D. Breakdown of SMM Reactions by Top-Level EC Category

Supplemental table 5E. Shared Reactions

Supplemental table 5F. Unique Reactions

Supplemental table 5G. Breakdown of Pathways by Pathway Class

Supplemental table 5 H. Shared Pathways

Supplemental table 5 I. Unique Pathways

Supplemental table 5 J. Pathway Holes

Supplemental table 5 K. Pathways involved in degradation of carbohydrate in studied *L. iners* strains

Supplemental table 5L. Cross-Species Comparison of lactose and galactose degradation I

**Supplemental Figure 1. Alpha diversity over time. Genus level changes over time**. A-B) Relative counts of Lactobacillus genus and L. iners before, during, and after chemoradiotherapy. C-I) Changes in the different alpha diversity and evenness indices before, during, and after chemoradiotherapy. Significant changes between each time point and baseline were determined using the Paired T-test.

**Supplemental Figure 2. Genome of L. iners isolated from 2 cervical cancer patients encode additional functions related to defense and pathogenesis**. A. Overlapping functions (KEGG KOs) of L. iners isolated from 2 cervical cancer patients (Pt1 and Pt2) and L. iners ATCC 55195 B. Molecular functions specific for L. iners isolates from cervical cancer. The functions resemble those identified in L. iners by WGS by comparison with the complete genome of L iners isolated from a healthy individual (see Fig. 2D).

**Supplemental Figure 3. Cellular overview of L. iners isolated from cervical swabs of patient pt1**. The diagram of lactose and galactose degradation I is colored in red. The lines show connections of compounds produced in the pathway to the same compounds in other reactions and pathways where the compound is produced (green line), consumed (purple line), consumed and/or produced (orange line). The overview demonstrates that the organism has limited metabolic capabilities consistent with the pathogenic phenotype and metabolites of lactose and galactose degradation I pathways play an important role in metabolism of the organism.

**Supplemental Figure 4. IFNg+ CD8+ cells and Ki67+ CD8 cells in co-culture experiments with Lactobacillus species**. In co-culture experiments with various Lactobacillus species and patient-derived PBMCs IFNg (A) and Ki67 (B) expression is not different.

**Supplemental Figure 5. Rarefaction curves and association network by batch for each timepoint/ response endpoint**. Alpha rarefaction curves for baseline (A) and week 5 (B) samples colored by batch. C-F) Association networks of all samples included in the study colored by batch (C), institution (D), clinical response (E), and individual patient (F). G-H) Association networks of baseline samples colored by batch (G) and clinical response (E)

**Supplemental information**. Original code used for analysis included in this manuscript

## REFERENCES

Aramaki, T. et al. (2020) ‘KofamKOALA: KEGG Ortholog assignment based on profile HMM and adaptive score threshold’, Bioinformatics (Oxford, England), 36(7), pp. 2251–2252. doi:10.1093/bioinformatics/btz859.

Arbyn, M. et al. (2020) ‘Estimates of incidence and mortality of cervical cancer in 2018: a worldwide analysis’, The Lancet. Global Health, 8(2), pp. e191–e203. doi:10.1016/S2214-109X(19)30482-6.

Ariyoshi, T. et al. (2022) ‘Effect of Clostridium butyricum on Gastrointestinal Infections’, Biomedicines, 10(2), p. 483. doi:10.3390/biomedicines10020483.

Artym, J. and Zimecki, M. (2021) ‘Antimicrobial and Prebiotic Activity of Lactoferrin in the Female Reproductive Tract: A Comprehensive Review’, Biomedicines, 9(12), p. 1940. doi:10.3390/biomedicines9121940.

Bissett, D.L. and Anderson, R.L. (1973) ‘Lactose and D0galactose metabolism in Staphylococcus aureus: pathway of D-galactose 6-phosphate degradation’, Biochemical and Biophysical Research Communications, 52(2), pp. 641–647. doi:10.1016/0006-291x(73)90761-4.

Bissett, D.L. and Anderson, R.L. (1980) ‘Lactose and D-galactose metabolism in Staphylococcus aureus. III. Purification and properties of D-tagatose-6-phosphate kinase’, The Journal of Biological Chemistry, 255(18), pp. 8745–8749.

Bolyen, E. et al. (2019) ‘Reproducible, interactive, scalable and extensible microbiome data science using QIIME 2’, Nature Biotechnology, 37(8), pp. 852–857. doi:10.1038/s41587-019-0209-9.

Bray, F. et al. (2018) ‘Global cancer statistics 2018: GLOBOCAN estimates of incidence and mortality worldwide for 36 cancers in 185 countries’, CA: a cancer journal for clinicians, 68(6), pp. 394–424. doi:10.3322/caac.21492.

Button, D.K. et al. (1973) ‘Carbohydrate transport in staphylococcus aureus IV. Maltose accumulation and metabolism’, Biochemical and Biophysical Research Communications, 52(3), pp. 850–855. doi:10.1016/0006-291X(73)91015-2.

Chase, D. et al. (2015) ‘The vaginal and gastrointestinal microbiomes in gynecologic cancers: a review of applications in etiology, symptoms and treatment’, Gynecologic Oncology, 138(1), pp. 190–200. doi:10.1016/j.ygyno.2015.04.036.

Choi, H.S. et al. (2006) ‘Hydrogen peroxide producing lactobacilli in women with cervical neoplasia’, Cancer Research and Treatment, 38(2), pp. 108–111. doi:10.4143/crt.2006.38.2.108.

Cohen, C.R. et al. (2020) ‘Randomized Trial of Lactin-V to Prevent Recurrence of Bacterial Vaginosis’, The New England Journal of Medicine, 382(20), pp. 1906–1915. doi:10.1056/NEJMoa1915254.

Colbert, L.E. et al. (2022) ‘Expansion of Candidate HPV-Specific T Cells in the Tumor Microenvironment during Chemoradiotherapy Is Prognostic in HPV16+ Cancers’, Cancer Immunology Research, 10(2), pp. 259–271. doi:10.1158/2326-6066.CIR-21-0119.

Dorta-Estremera, S. et al. (2018) ‘Kinetics of Intratumoral Immune Cell Activation During Chemoradiation for Cervical Cancer’, International Journal of Radiation Oncology, Biology, Physics, 102(3), pp. 593–600. doi:10.1016/j.ijrobp.2018.06.404.

Eriksson, P. et al. (2021) ‘A comparison of rule-based and centroid single-sample multiclass predictors for transcriptomic classification’, Bioinformatics (Oxford, England), p. btab763. doi:10.1093/bioinformatics/btab763.

Eschenbach, D.A. et al. (1989) ‘Prevalence of hydrogen peroxide-producing Lactobacillus species in normal women and women with bacterial vaginosis’, Journal of Clinical Microbiology, 27(2), pp. 251–256. doi:10.1128/jcm.27.2.251-256.1989.

Fan, Q. et al. (2021) ‘Lactobacillus spp. create a protective micro-ecological environment through regulating the core fucosylation of vaginal epithelial cells against cervical cancer’, Cell Death & Disease, 12(12), pp. 1–13. doi:10.1038/s41419-021-04388-y.

Hamady, M. et al. (2008) ‘Error-correcting barcoded primers for pyrosequencing hundreds of samples in multiplex’, Nature Methods, 5(3), pp. 235–237. doi:10.1038/nmeth.1184.

Hamady, M. and Knight, R. (2009) ‘Microbial community profiling for human microbiome projects: Tools, techniques, and challenges’, Genome Research, 19(7), pp. 1141–1152. doi:10.1101/gr.085464.108.

Homburg, C. et al. (2017) ‘Inducer exclusion in Firmicutes: insights into the regulation of a carbohydrate ATP binding cassette transporter from Lactobacillus casei BL23 by the signal transducing protein P-Ser46-HPr’, Molecular Microbiology, 105(1), pp. 25–45. doi:10.1111/mmi.13680.

Hyatt, D. et al. (2010) ‘Prodigal: prokaryotic gene recognition and translation initiation site identification’, BMC bioinformatics, 11, p. 119. doi:10.1186/1471-2105-11-119.

Jain, C. et al. (2018) ‘High throughput ANI analysis of 90K prokaryotic genomes reveals clear species boundaries’, Nature Communications, 9(1), p. 5114. doi:10.1038/s41467-018-07641-9.

Kang, D.D. et al. (2019) ‘MetaBAT 2: an adaptive binning algorithm for robust and efficient genome reconstruction from metagenome assemblies’, PeerJ, 7, p. e7359. doi:10.7717/peerj.7359.

Karp, P.D. et al. (2020) ‘Pathway Tools version 24.0: Integrated Software for Pathway/Genome Informatics and Systems Biology’, 1510.03964 [q-bio] [Preprint]. Available at: http://arxiv.org/abs/1510.03964 (Accessed: 31 March 2022).

Karp, P.D., Latendresse, M. and Caspi, R. (2011) ‘The pathway tools pathway prediction algorithm’, Standards in Genomic Sciences, 5(3), pp. 424–429. doi:10.4056/sigs.1794338.

Kwak, W. et al. (2020) ‘Complete Genome of Lactobacillus iners KY Using Flongle Provides Insight Into the Genetic Background of Optimal Adaption to Vaginal Econiche’, Frontiers in Microbiology, 11, p. 1048. doi:10.3389/fmicb.2020.01048.

Langmead, B. and Salzberg, S.L. (2012) ‘Fast gapped-read alignment with Bowtie 2’, Nature Methods, 9(4), pp. 357–359. doi:10.1038/nmeth.1923.

Lebeer, S., Vanderleyden, J. and De Keersmaecker, S.C.J. (2008) ‘Genes and Molecules of Lactobacilli Supporting Probiotic Action’, Microbiology and Molecular Biology Reviews, 72(4), pp. 728–764. doi:10.1128/MMBR.00017-08.

Macklaim, J.M. et al. (2011) ‘At the crossroads of vaginal health and disease, the genome sequence of Lactobacillus iners AB-1’, Proceedings of the National Academy of Sciences of the United States of America, 108 Suppl 1, pp. 4688–4695. doi:10.1073/pnas.1000086107.

McMillan, A. et al. (2013) ‘Adhesion of Lactobacillus iners AB-1 to human fibronectin: a key mediator for persistence in the vagina?’, Reproductive Sciences (Thousand Oaks, Calif.), 20(7), pp. 791–796. doi:10.1177/1933719112466306.

von Meijenfeldt, F.A.B. et al. (2019) ‘Robust taxonomic classification of uncharted microbial sequences and bins with CAT and BAT’, Genome Biology, 20(1), p. 217. doi:10.1186/s13059-019-1817-x.

MetaCyc database of metabolic pathways and enzymes | Nucleic Acids Research | Oxford Academic (no date). Available at: https://academic.oup.com/nar/article/46/D1/D633/4559117 (Accessed: 1 April 2022).

Nataraj, B.H. and Mallappa, R.H. (2021) ‘Antibiotic Resistance Crisis: An Update on Antagonistic Interactions between Probiotics and Methicillin-Resistant Staphylococcus aureus (MRSA)’, Current Microbiology, 78(6), pp. 2194–2211. doi:10.1007/s00284-021-02442-8.

Nearing, J.T. et al. (2018) ‘Denoising the Denoisers: an independent evaluation of microbiome sequence error-correction approaches’, PeerJ, 6, p. e5364. doi:10.7717/peerj.5364.

Nurk, S. et al. (2017) ‘metaSPAdes: a new versatile metagenomic assembler’, Genome Research, 27(5), pp. 824–834. doi:10.1101/gr.213959.116.

Petrova, M.I. et al. (2017) ‘Lactobacillus iners : Friend or Foe?’, Trends in Microbiology, 25(3), pp. 182–191. doi:10.1016/j.tim.2016.11.007.

Pruesse, E. et al. (2007) ‘SILVA: a comprehensive online resource for quality checked and aligned ribosomal RNA sequence data compatible with ARB’, Nucleic Acids Research, 35(21), pp. 7188–7196. doi:10.1093/nar/gkm864.

Rampersaud, R. et al. (2011) ‘Inerolysin, a cholesterol-dependent cytolysin produced by Lactobacillus iners’, Journal of Bacteriology, 193(5), pp. 1034–1041. doi:10.1128/JB.00694-10.

RStudio Team (2020) RStudio: Integrated Development for R. Boston, MA: RStudio, Inc. Available at: http://www.rstudio.com/.

Scrucca, L. et al. (2016) ‘mclust 5: Clustering, Classification and Density Estimation Using Gaussian Finite Mixture Models’, The R Journal, 8(1), pp. 289–317.

Segata, N. et al. (2011) ‘Metagenomic biomarker discovery and explanation’, Genome Biology, 12(6), p. R60. doi:10.1186/gb-2011-12-6-r60.

Vaupel, P., Schmidberger, H. and Mayer, A. (2019) ‘The Warburg effect: essential part of metabolic reprogramming and central contributor to cancer progression’, International Journal of Radiation Biology, 95(7), pp. 912–919. doi:10.1080/09553002.2019.1589653.

Verhelst, R. et al. (2005) ‘Comparison between Gram stain and culture for the characterization of vaginal microflora: Definition of a distinct grade that resembles grade I microflora and revised categorization of grade I microflora’, BMC Microbiology, 5(1), p. 61. doi:10.1186/1471-2180-5-61.

Witkin, S.S. et al. (2013) ‘Influence of Vaginal Bacteria and D - and L -Lactic Acid Isomers on Vaginal Extracellular Matrix Metalloproteinase Inducer: Implications for Protection against Upper Genital Tract Infections’, mBio. Edited by M.J. Blaser, 4(4). doi:10.1128/mBio.00460-13.

Yang, X. et al. (2018) ‘Role of Lactobacillus in cervical cancer’, Cancer Management and Research, 10, pp. 1219–1229. doi:10.2147/CMAR.S165228.

Yuh, W.T.C. et al. (2009) ‘Predicting control of primary tumor and survival by DCE MRI during early therapy in cervical cancer’, Investigative Radiology, 44(6), pp. 343–350. doi:10.1097/RLI.0b013e3181a64ce9.

Zadravec, P., Štrukelj, B. and Berlec, A. (2015) ‘Improvement of LysM-mediated surface display of designed ankyrin repeat proteins (DARPins) in recombinant and nonrecombinant strains of Lactococcus lactis and Lactobacillus Species’, Applied and Environmental Microbiology, 81(6), pp. 2098–2106. doi:10.1128/AEM.03694-14.

