## Supplementary material for "Cancer-associated *Lactobacillus iners* are genetically distinct and associated with chemoradiation resistance in cervical cancer": Key Resources Table

| REAGENT or RESOURCE | SOURCE | IDENTIFIER |
| --- | --- | --- |
| Antibodies | | |
| CD33 antibody | BD Biosciences | Cat# 740293; RRID:AB_2740032 |
| CD86 (B7-2) antibody | BD Biosciences | Cat# 564428; RRID:AB_2738804) |
| CD141 antibody | BD Biosciences | Cat# 565321; RRID:AB_2739180) |
| Brilliant Violet 605(TM) anti-human CD11b antibody | BioLegend | Cat# 301332; RRID:AB_2562021) |
| Brilliant Violet 711(TM) anti-human Ki-67 antibody | BioLegend | Cat# 350516; RRID:AB_2563861) |
| CD152 (CTLA-4) antibody | BD Biosciences | Cat# 563931; RRID:AB_2738491) |
| CD3 antibody | BD Biosciences | Cat# 560835; RRID:AB_2033956) |
| PE anti-human CD45 antibody | BioLegend | Cat# 368510; RRID:AB_2566370) |
| Granzyme B antibody | BD Biosciences | Cat# 562462; RRID:AB_2737618) |
| CD11c antibody | BD Biosciences | Cat# 551077; RRID:AB_394034) |
| CD4 Monoclonal Antibody (S3.5),  PE-Alexa Fluor 700 | Thermo Fisher Scientific | Cat# MHCD0424; RRID:AB_10372802) |
| PKR Polyclonal Antibody | Thermo Fisher Scientific | Cat# PA1-990; RRID:AB_560651) |
| Mouse Anti-CD69 Monoclonal Antibody | BD Biosciences | Cat# 555533; RRID:AB_398602) |
| Alexa Fluor(R) 700 anti-human CD8a antibody | BioLegend | Cat# 300920; RRID:AB_528885) |
| APC/Cyanine7 anti-human CD279 (PD-1) antibody | BioLegend | Cat# 329922; RRID:AB_10933429) |
| Biological Samples |  |  |
| Human Tumor Sampling - Cytobrush | The University of Texas MD Anderson Cancer Center, Houston, TX | Protocol 2014-0543? |
| Chemicals, Peptides, and Recombinant Proteins |  |  |
| MRS Agar Plates | Moltox | Cat# 51-40S020.140 |
| TSA Agar Plates (BD BBL™ Trypticase™ Soy Agar with 5% Sheep Blood (BD Trypticase™ Soy Agar II™)) | BD Biosciences | Cat# 221239 |
| Lactobacillus MRS Dehydrated Culture | BD Difco | Cat# 288210 |
| HEPES | Fisher Bioreagents | Cat# BP310100 |
| Proteose Peptone No. 3 | Bacto | Cat# C838M71 |
| NaCl | Fisher Chemical | Cat# S271500 |
| D-Glucose | Avantor Macron | Cat# 491212 |
| Fresh yeast extract | Gibco | Cat# 18180059 |
| Heat Inactivated Horse Serum | Remel | Cat# R55075 |
| Corning™ Minimum Essential Medium Eagle (Mod.) 1X (MEM) with Glutamine | Corning | Cat# 10010CV |
| Corning® Fetal Bovine Serum (FBS), 500 mL, Regular, USDA Approved Origin | Corning | Cat# 35010CV |
| Corning® 100 mL Penicillin-Streptomycin Solution, 100x | Corning | Cat# 30002CI |
| Gentra Puregene Yeast/Bact. Kit | Qiagen | Cat# 158567 |
| Cisplatin (cis-Diammineplatinum(II) dichloride) | Sigma-Aldrich | Cat# [P4394](https://www.sigmaaldrich.com/US/en/product/sigma/p4394) |
| CellTiter-Glo Luminescent Cell Viability Assay | Promega | Cat# G7572 |
| LIVE/DEAD™ Fixable Aqua Dead Cell Stain Kit, for 405 nm excitation | Thermo Fisher | Cat# L34957 |
| Phytohemagglutinin (PHA) |  |  |
| Golgi Plug | BD Biosciences | 555029 |
| Deposited Data |  |  |
| 16S | SRA |  |
| WGS | SRA |  |
| *L. iners* strain assemblies | Genome | BioSample accession numbers: SAMN27176861, SAMN27176862, SAMN27176863, SAMN27176864 |
| Experimental Models: Cell Lines | | |
| C-33 A | ATCC | ATCC Cat# HTB-31, RRID:CVCL_1094 |
| HeLa | Sam Mok lab, MD Anderson Cancer Center | RRID:CVCL_0030 |
| Experimental Models: Organisms/Strains |  |  |
| ATCC *Lactobacillus iners* | ATCC | ATCC 55195; NCBITaxon:888801 |
| ATCC *Lactobacillus crispatus* SJ-3C-US | ATCC | ATCC SJ-3C (PTA-10138);  NCBITaxon:575598 |
| Software and Algorithms |  |  |
| ATIMA (Agile Toolkit for Incisive Microbial Analyses) | R Studio | http://atima.jplab.net/ |
| Galaxy: LEfSe | Segata et. al 2010 | https://huttenhower.sph.harvard.edu/galaxy/ |
| Other (Resources) |  |  |
| FlowJo | FlowJo Software | www.flowjo.com |
| GraphPad Prism | GraphPad Software | www.graphpad.com |
| JMP | SAS | www.jmp.com |
| Living Image Software | IVIS Spectrum – Perkin Elmer | Part #128113 |
| FastQC (version 0.11.8) |  | https://www.bioinformatics.babraham.ac.uk/projects/fastqc |
| FastQ_Screen (version 0.14.0) | (Wingett and Andrews 2018) | https://www.bioinformatics.babraham.ac.uk/projects/fastq_screen |
| BBDuk/BBTools (version 20190109) |  | https://jgi.doe.gov/data-and-tools/bbtools |
| BBMap/BBTools (version 20190109) |  | https://jgi.doe.gov/data-and-tools/bbtools |
| Bowtie2 (version 2.3.5) | (Langmead and Salzberg 2012) | https://github.com/BenLangmead/bowtie2 |
| Samtools (version 1.9) | (H. Li et al. 2009) | https://github.com/samtools |
| IGGsearch (version 20190921) | (Nayfach et al. 2019) | https://github.com/snayfach/IGGsearch |
| MetaPhlAn2 (version 2.0) (mpa_v295_CHOCOPhlAn_201901) | (Truong et al. 2015) | https://bitbucket.org/biobakery/metaphlan2 |
| MEGAHIT (version 1.2.8) | (D. Li et al. 2015) | https://github.com/voutcn/megahit |
| metaSPAdes (version 3.13.1) | (Nurk et al. 2017) | https://github.com/ablab/spades/releases |
| MetaBAT2 (version 20191004) | (Kang et al. 2019) | https://bitbucket.org/berkeleylab/metabat |
| Prodigal (version 2.6.3) | (Hyatt et al. 2010) | https://github.com/hyattpd/Prodigal |
| Diamond (version 0.9.24) | (Buchfink, Xie, and Huson 2015) | https://github.com/bbuchfink/diamond |
| KofamScan (version 1.1.0) | (Aramaki et al. 2020) | https://github.com/takaram/kofam_scan |
| Subread (version 1.6.3) | (Liao, Smyth, and Shi 2014) | http://subread.sourceforge.net |
| DFAST (version **2021.7.12)** | <https://doi.org/10.1093/bioinformatics/btx713> | https://dfast.ddbj.nig.ac.jp |
| MetaCyc (version 25.5) | <https://doi.org/10.1093/nar/gkx935> | https://biocyc.org/download-bundle.shtml |
| Pathway Tools (version 25.5) | <https://doi.org/10.1093/bib/bbv079> | https://biocyc.org/download-bundle.shtml |
| FastANI | DOI: [10.1038/s41467-018-07641-9](https://doi-org.elibrary.mdanderson.org/10.1038/s41467-018-07641-9) | https://github.com/ParBLiSS/FastANI |
| QIIME2 (version 2020.11) |  | https://www.nature.com/articles/s41587-019-0209-9 |
| DADA2 |  | doi:10.7717/peerj.5364 |
