## Supplementary figures and images for "Cancer-associated *Lactobacillus iners* are genetically distinct and associated with chemoradiation resistance in cervical cancer"

### Supplemental Figure 1

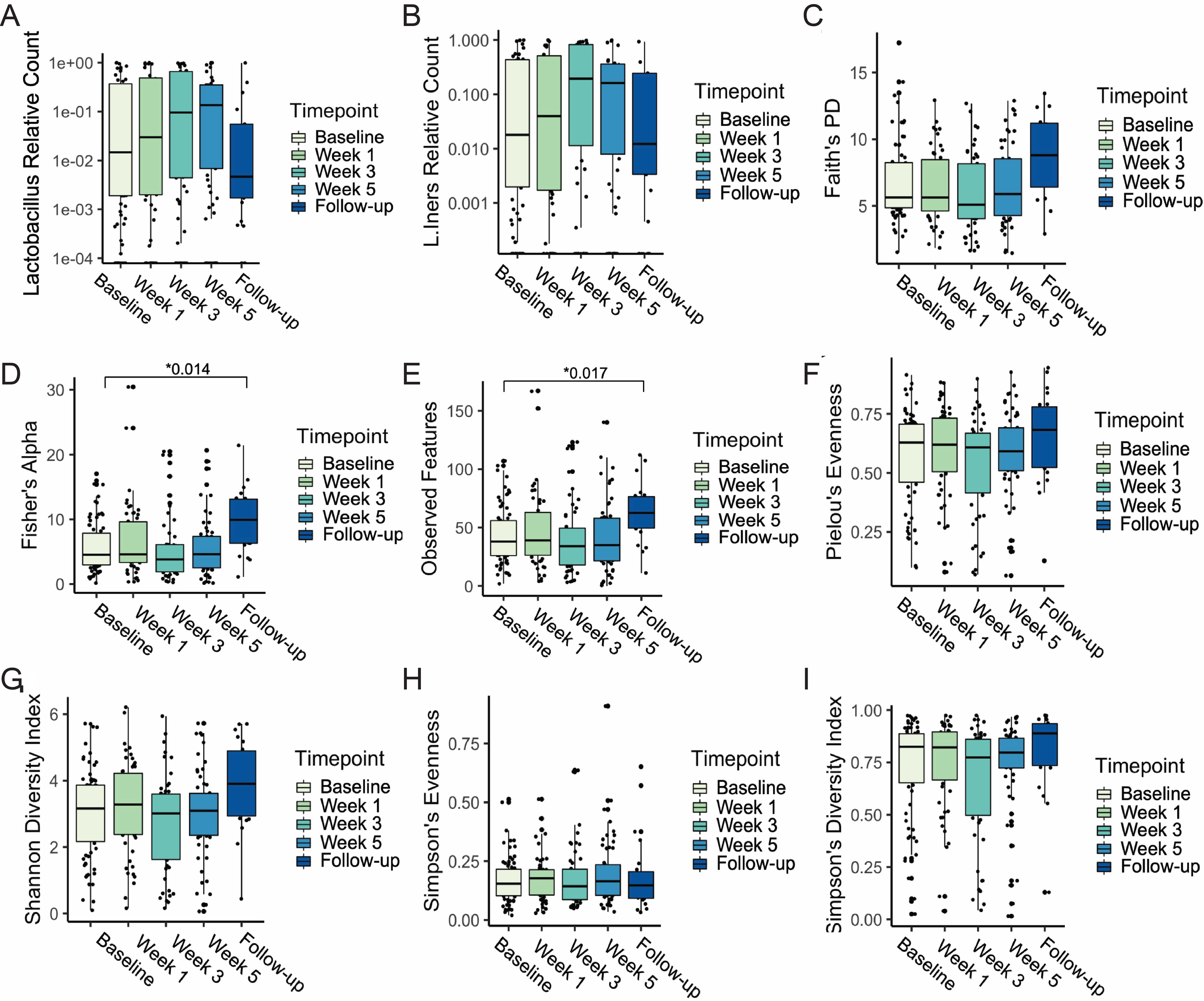

### Supplemental Figure 2

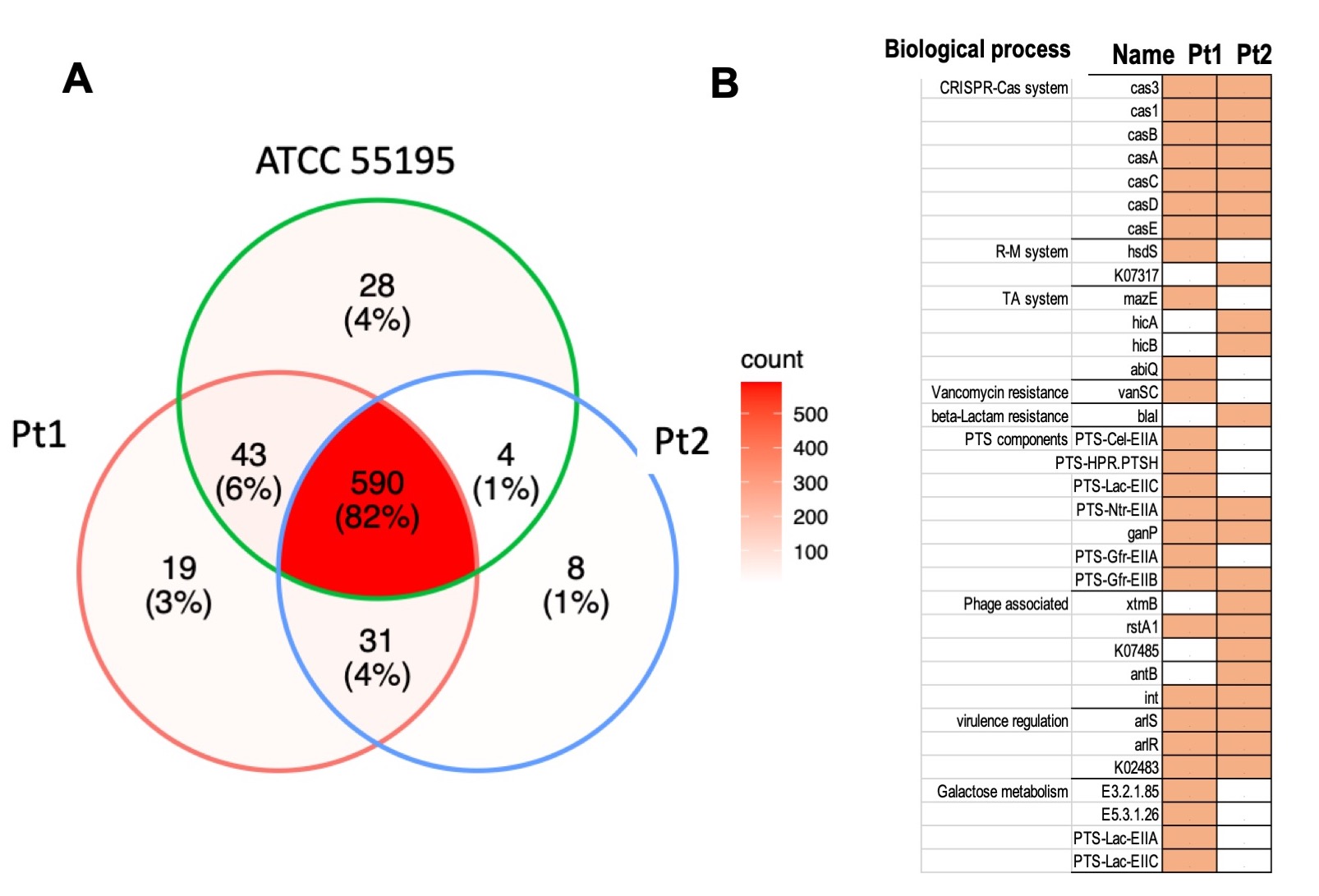

### Supplemental Figure 3

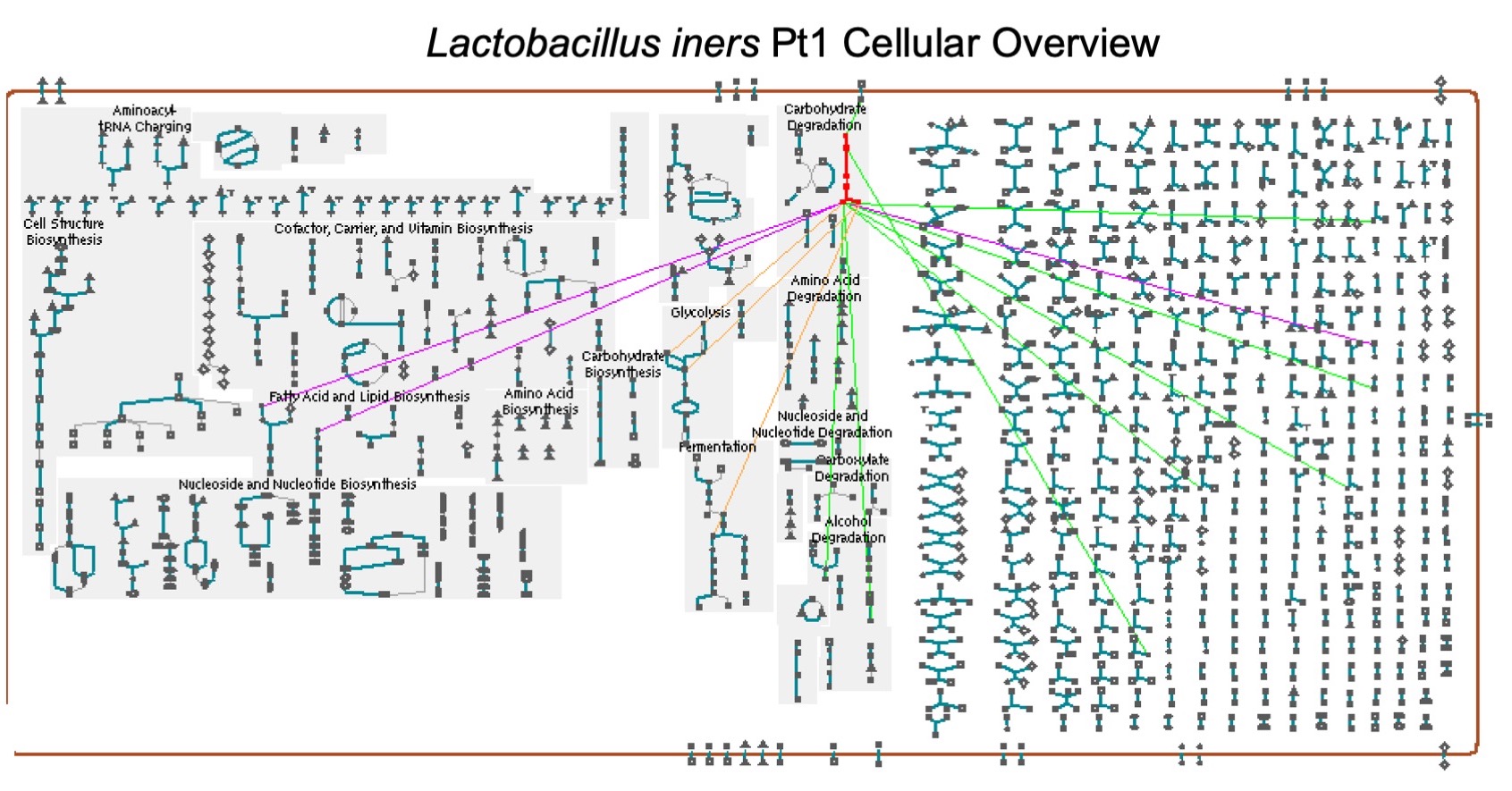

### Supplemental Figure 4

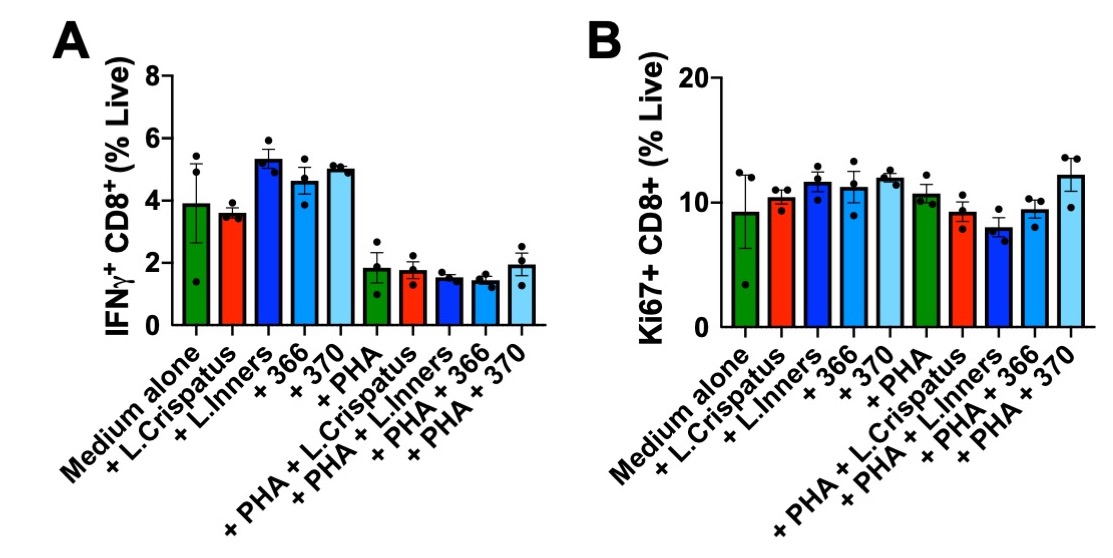

### Supplemental Figure 5

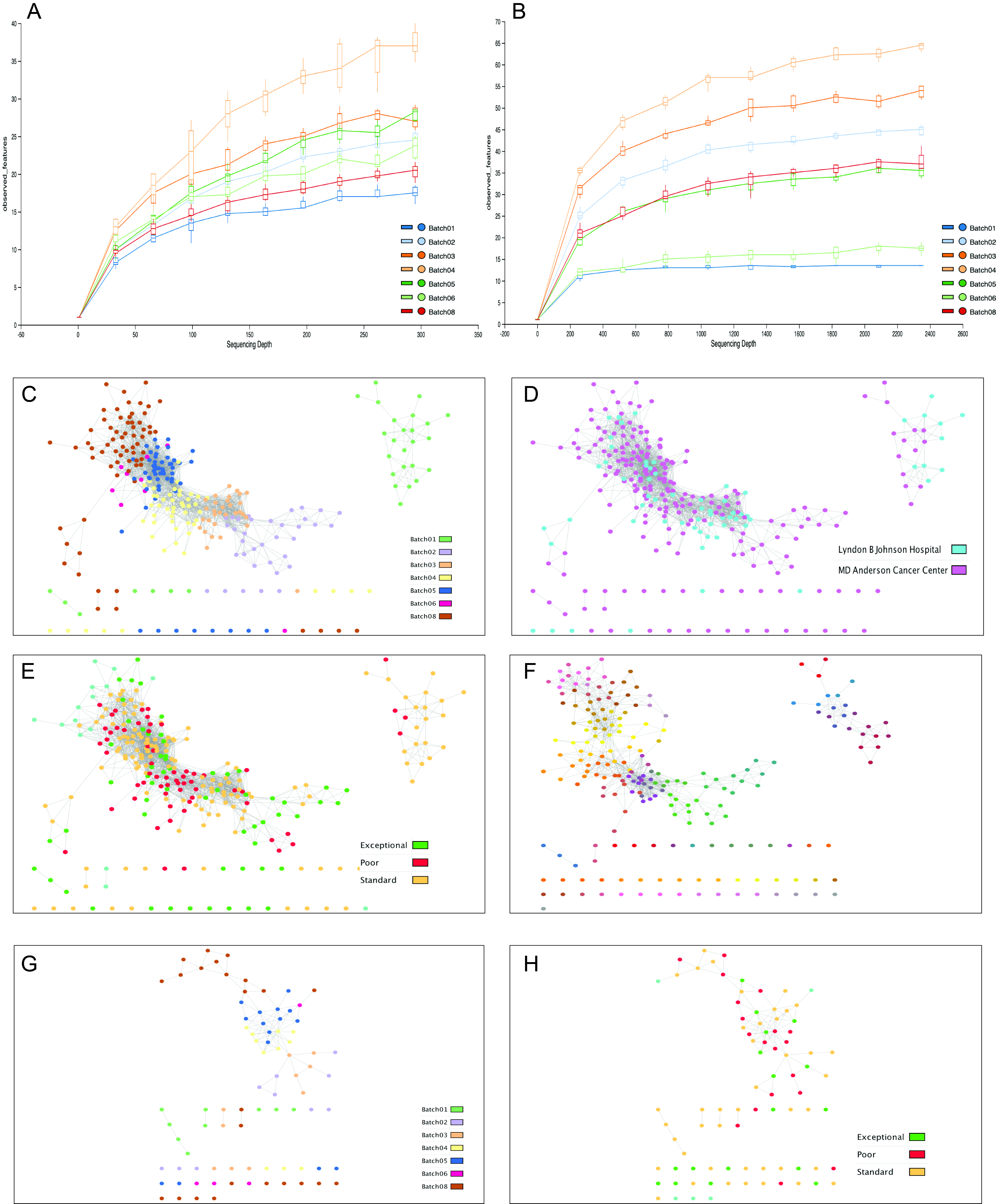
