## Supplemental Table 1 for "Cancer-associated *Lactobacillus iners* are genetically distinct and associated with chemoradiation resistance in cervical cancer"

Supplemental Table 1. Patient Characteristics (N=78)

|  | **N** | **%** | |
| --- | --- | --- | --- |
| **Age**, Mean (Range), Yrs | 47.65 (25.6 – 79.4) |  | |
| **BMI,** Mean (Range), Kg/m^2^ | 29.43 (18.50 – 54) |  | |
| **Institution** | | | |
| Lyndon B. Johnson Hospital | 23 | | 29.49 |
| MD Anderson Cancer Center | 55 | | 70.51 |
| **Race** | | | |
| Hispanic | 37 | 47.44 | |
| White | 32 | 41.03 | |
| Black | 5 | 6.41 | |
| Asian | 2 | 2.56 | |
| Other | 2 | 2.56 | |
| **Smoking** | | | |
| Current | 7 | 9.09 | |
| Former | 26 | 33.77 | |
| Never | 44 | 57.14 | |
| **Histology** | | | |
| Squamous Carcinoma | 64 | 82.05 | |
| Adenocarcinoma | 11 | 14.10 | |
| Adenosquamous Carcinoma | 3 | 3.85 | |
| **Lymphovascular space invasion (LVSI)** | | | |
| Yes | 6 | 7.69 | |
| No | 18 | 23.08 | |
| Unknown | 54 | 69.23 | |
| **Tumor Size**, Mean (Range), Cm | 5.27 (1.2 – 10.0) |  | |
| **FIGO Stage** | | | |
| IA1 | 1 | 1.28 | |
| IB1 | 5 | 6.41 | |
| IB2 | 9 | 11.54 | |
| IB3 | 5 | 6.41 | |
| IIA | 3 | 3.85 | |
| IIB | 29 | 37.18 | |
| IIIB | 13 | 16.67 | |
| IIIC1 | 7 | 8.97 | |
| IIIC2 | 1 | 1.28 | |
| IVA | 4 | 5.13 | |
| IVB | 1 | 1.28 | |
| **Grade** | | | |
| 1 | 3 | 4.00 | |
| 2 | 29 | 38.67 | |
| 3 | 27 | 36.00 | |
| Other | 3 | 4.00 | |
| Unknown | 13 | 17.33 | |
| **Nodes** | | | |
| Common Iliac | 14 | 18.67 | |
| External Iliac | 24 | 32.00 | |
| Internal Iliac | 4 | 5.33 | |
| Para-Aortic | 6 | 8.00 | |
| None | 25 | 33.33 | |
| Unknown | 2 | 2.67 | |
| **HPV** | | | |
| HPV18 | 8 | 10.81 | |
| HPV16 | 36 | 48.65 | |
| Negative | 13 | 17.57 | |
| Other | 17 | 22.97 | |
| **Cisplatin Cycles** | | | |
| 0 | 2 | 2.56 | |
| 1 | 0 | 0.00 | |
| 2 | 2 | 2.56 | |
| 3 | 9 | 11.54 | |
| 4 | 6 | 7.69 | |
| 5 | 28 | 35.90 | |
| 6 | 30 | 38.46 | |
| 7 | 1 | 1.28 | |
| **Radiation Dose**, Mean (Range), Gy | 8,288 ( 4,500 – 11,531) |  | |
| **Response** | | | |
| Exception | 17 | 21.79 | |
| Standard | 40 | 51.28 | |
| Poor | 21 | 26.92 | |
| **First Recurrence** | | | |
| In Field | 14 | 51.85 | |
| Out of Field | 13 | 48.15 | |
| **Survival** | | | |
| Alive | 68 | 87.17 | |
| Dead | 10 | 12.82 | |
| **Antibiotic Use During Study Period** | | | |
| Yes | 35 | 45.45 | |
| No | 42 | 54.54 | |
