## Supplemental Table 3 for "Cancer-associated *Lactobacillus iners* are genetically distinct and associated with chemoradiation resistance in cervical cancer"

Supplemental Table 3. Association of L. iners with patient characteristics

|  | ***Lactobacillus Iners ....*** | |
| --- | --- | --- |
| **Demographics** | **Estimate (95% CI)** | **P** |
| **Institution** |  |  |
| Lyndon B. Johnson Hospital | – | – |
| MD Anderson Cancer Center | 0.08 (-0.05-0.21) | 0.22 |
| **Age** | 0 (-0.01-0) | 0.15 |
| **Race** |  |  |
| Asian | – | – |
| Black | -0.52 (-0.95- -0.09) | **0.02** |
| Hispanic | -0.39 (-0.75- -0.03) | **0.03** |
| Other | -0.52 (-1.02- -0.03) | **0.04** |
| White | -0.44 (-0.8- -0.08) | **0.02** |
| **BMI** | -0.01 (-0.02-0) | 0.14 |
| **Smoking Status** |  |  |
| Current | – | – |
| Former | -0.06 (-0.29-0.18) | 0.62 |
| Never | -0.05 (-0.27-0.18) | 0.67 |
| **Histology** |  |  |
| Adenocarcinoma | – | – |
| Adenosquamous Carcinoma | 0.31 (-0.01-0.63) | 0.06 |
| Squamous Cell Carcinoma | -0.08 (-0.24-0.08) | 0.35 |
| **LVSI** |  |  |
| No | – | – |
| Yes | 0.26 (0.02-0.49) | **0.03** |
| Unknown | 0.01 (-0.13-0.15) | 0.87 |
| **Stage** |  |  |
| Stage 1-2 | – | – |
| Stage 3-4 | -0.07 (-0.2-0.05) | 0.25 |
| **Grade** |  |  |
| Grade 1 | – | – |
| Grade 2 | 0.07 (-0.21-0.36) | 0.61 |
| Grade 3 | 0.08 (-0.2-0.37) | 0.57 |
| Other | 0.33 (-0.05-0.71) | 0.09 |
| Unknown | 0.12 (-0.18-0.42) | 0.42 |
| **HPV** |  |  |
| HPV 16 | – | – |
| HPV 18 | 0.28 (0.06-0.5) | **0.01** |
| Negative | 0.02 (-0.14-0.18) | 0.83 |
| Other | 0.02 (-0.13-0.17) | 0.78 |
| **Cisplatin Cycles** | 0.01 (-0.04-0.05) | 0.75 |
| **Radiation Dose** | 0 (0-0) | 0.89 |
| **Antibiotic Use** |  |  |
| No | – | – |
| Yes | 0.04 (-0.08-0.16) | 0.52 |
| **Node** |  |  |
| Pelvic Node | – | – |
| None | 0.02 (-0.1-0.15) | 0.71 |
| Para- Aortic Node | -0.02 (-0.23-0.2) | 0.88 |
| **Tumor Dimension** | -0.02 (-0.06-0.01) | 0.11 |
