## Supplemental Table 4 for "Cancer-associated *Lactobacillus iners* are genetically distinct and associated with chemoradiation resistance in cervical cancer"

| **Strain** | ATCC 55195 | Pt2 | Pt1 | I012T4 |
| --- | --- | --- | --- | --- |
| **PGDB name** | IN55195 | IN370 | IN366 | I012T4 |
| **Total Sequence Length (bp):** | 1249908 | 1228104 | 1339170 | 1211893 |
| **Number of Sequences:** | 114 | 111 | 149 | 14 |
| **Longest Sequences (bp):** | 286503 | 226335 | 413171 | 242589 |
| ***N50 (bp):** | 144291 | 138957 | 148365 | 146463 |
| **GCcontent (%):** | 32.8 | 33.1 | 32.8 | 32.4 |
| **Number of CDSs:** | 1119 | 1115 | 1209 | **1150** |
| **Average Protein Length:** | 319.3 | 311.1 | 314 | 318 |
| **Coding Ratio (%):** | 85.8 | 84.7 | 85 | 91 |
| **Number of rRNAs:** | 6 | 9 | 6 | 2 |
| **Number of tRNAs:** | 63 | 55 | 60 | 30 |
| **Number of CRISPRs:** | 1 | 11 | 8 | 0 |

*N50 (bp) is a weighted median statistic such that 50% of the entire assembly is contained in contigs or scaffolds equal to or larger than 50 bp.
