## Supplemental Table 5 for "Cancer-associated *Lactobacillus iners* are genetically distinct and associated with chemoradiation resistance in cervical cancer"

**Supplemental Table 5. Comparative Analysis Summary Results**

Note: In addition to reflecting differences in biology of different organisms, these statistics will reflect differences in the levels of curation, data availability, and completeness of the PGDBs for these organisms.

Comparative analysis and statistics were computed for the following organism databases:

- *Lactobacillus iners ATCC 55195*
- *Lactobacillus iners I012T4*
- *Lactobacillus iners KY*
- *Lactobacillus iners pt1*
- *Lactobacillus iners pt2*

Metadata of the organisms:

- *L. iners* ATCC 55195 is a draft genome of *L.iners* ATCC 55195 strain isolated form a patient with bacterial vaginosis
- *L. iners* I012T4 is the high quality draft genome of *L.iners* assembled from WGS sequencing data of a cervical swab of CC patient.
- *L. iners* KY is the complete genome of *L.iners* isolated from a healthy individual
- *L. iners* pt1 and *L.iners* pt2 are draft genomes of *L.iners*  isolated from cervical swabs of 2 CC patients

**Table of Contents**

- **Organism**
  - Supplemental Table 5A: Database Summary Statistics
- **Reactions**
  - Supplemental Table 5B: Breakdown of Reactions by Type
  - Supplemental Table 5C: Reactions of Small Molecule Metabolism
  - Supplemental Table 5D: Breakdown of SMM Reactions by Top-Level EC Category
  - Supplemental Table 5E: Shared Reactions
  - Supplemental Table 5F: Unique Reactions
- **Pathways**
  - Supplemental Table 5G: Breakdown of Pathways by Pathway Class
  - Supplemental Table 5H: Shared Pathways
  - Supplemental Table 5I: Unique Pathways
  - Supplemental Table 5J: Pathway Holes
  - Supplemental Table 5K. Pathways involved in degradation of carbohydrate in studied *L. iners* strains
  - Supplemental Table 5L: Cross-Species Comparison of lactose and galactose degradation I

**Organism**

**Supplemental Table 5A: Database Summary Statistics**

| Database | [*L.iners* ATCC 55195](http://localhost:1555/IN55195/organism-summary) | [*L.iners* I012T4](http://localhost:1555/I012T4/organism-summary) | [*L.iners* KY](http://localhost:1555/INKY1/organism-summary) | [*L.iners* pt1](http://localhost:1555/IN366/organism-summary) | [*L.iners* pt2](http://localhost:1555/IN370/organism-summary) |
| --- | --- | --- | --- | --- | --- |
| [Genome Size (bp)](http://localhost:1555/comp-genomics?type=(ORGANISM-STATS+GENOME-SIZE)&orgids=(IN55195+I012T4+INKY1+IN366+IN370)) | 1,217,911 | 1,211,893 | 1,339,101 | 1,292,390 | 1,191,294 |
| [Chromosomes](http://localhost:1555/comp-genomics?type=(ORGANISM-STATS+CHROMOSOMES)&orgids=(IN55195+I012T4+INKY1+IN366+IN370)) | [0](http://localhost:1555/comp-genomics?type=(ORGANISM-STATS+CHROMOSOMES)&orgids=IN55195) | [0](http://localhost:1555/comp-genomics?type=(ORGANISM-STATS+CHROMOSOMES)&orgids=I012T4) | [1](http://localhost:1555/comp-genomics?type=(ORGANISM-STATS+CHROMOSOMES)&orgids=INKY1) | [0](http://localhost:1555/comp-genomics?type=(ORGANISM-STATS+CHROMOSOMES)&orgids=IN366) | [0](http://localhost:1555/comp-genomics?type=(ORGANISM-STATS+CHROMOSOMES)&orgids=IN370) |
| [Organelle Chromosomes](http://localhost:1555/comp-genomics?type=(ORGANISM-STATS+ORGANELLE-CHROMOSOMES)&orgids=(IN55195+I012T4+INKY1+IN366+IN370)) | [0](http://localhost:1555/comp-genomics?type=(ORGANISM-STATS+ORGANELLE-CHROMOSOMES)&orgids=IN55195) | [0](http://localhost:1555/comp-genomics?type=(ORGANISM-STATS+ORGANELLE-CHROMOSOMES)&orgids=I012T4) | [0](http://localhost:1555/comp-genomics?type=(ORGANISM-STATS+ORGANELLE-CHROMOSOMES)&orgids=INKY1) | [0](http://localhost:1555/comp-genomics?type=(ORGANISM-STATS+ORGANELLE-CHROMOSOMES)&orgids=IN366) | [0](http://localhost:1555/comp-genomics?type=(ORGANISM-STATS+ORGANELLE-CHROMOSOMES)&orgids=IN370) |
| [Plasmids](http://localhost:1555/comp-genomics?type=(ORGANISM-STATS+PLASMIDS)&orgids=(IN55195+I012T4+INKY1+IN366+IN370)) | [0](http://localhost:1555/comp-genomics?type=(ORGANISM-STATS+PLASMIDS)&orgids=IN55195) | [0](http://localhost:1555/comp-genomics?type=(ORGANISM-STATS+PLASMIDS)&orgids=I012T4) | [0](http://localhost:1555/comp-genomics?type=(ORGANISM-STATS+PLASMIDS)&orgids=INKY1) | [0](http://localhost:1555/comp-genomics?type=(ORGANISM-STATS+PLASMIDS)&orgids=IN366) | [0](http://localhost:1555/comp-genomics?type=(ORGANISM-STATS+PLASMIDS)&orgids=IN370) |
| [Contigs](http://localhost:1555/comp-genomics?type=(ORGANISM-STATS+CONTIGS)&orgids=(IN55195+I012T4+INKY1+IN366+IN370)) | [27](http://localhost:1555/comp-genomics?type=(ORGANISM-STATS+CONTIGS)&orgids=IN55195) | [14](http://localhost:1555/comp-genomics?type=(ORGANISM-STATS+CONTIGS)&orgids=I012T4) | [0](http://localhost:1555/comp-genomics?type=(ORGANISM-STATS+CONTIGS)&orgids=INKY1) | [28](http://localhost:1555/comp-genomics?type=(ORGANISM-STATS+CONTIGS)&orgids=IN366) | [29](http://localhost:1555/comp-genomics?type=(ORGANISM-STATS+CONTIGS)&orgids=IN370) |
| [Genes](http://localhost:1555/comp-genomics?type=(ORGANISM-STATS+GENES)&orgids=(IN55195+I012T4+INKY1+IN366+IN370)) | [1,189](http://localhost:1555/comp-genomics?type=(ORGANISM-STATS+GENES)&orgids=IN55195) | [1,183](http://localhost:1555/comp-genomics?type=(ORGANISM-STATS+GENES)&orgids=I012T4) | [1,439](http://localhost:1555/comp-genomics?type=(ORGANISM-STATS+GENES)&orgids=INKY1) | [1,275](http://localhost:1555/comp-genomics?type=(ORGANISM-STATS+GENES)&orgids=IN366) | [1,180](http://localhost:1555/comp-genomics?type=(ORGANISM-STATS+GENES)&orgids=IN370) |
| [Genes of known or predicted molecular function](http://localhost:1555/comp-genomics?type=(ORGANISM-STATS+CURATED-GENES)&orgids=(IN55195+I012T4+INKY1+IN366+IN370)" \t "_blank) | [420](http://localhost:1555/comp-genomics?type=(ORGANISM-STATS+CURATED-GENES)&orgids=IN55195) | [510](http://localhost:1555/comp-genomics?type=(ORGANISM-STATS+CURATED-GENES)&orgids=I012T4) | [400](http://localhost:1555/comp-genomics?type=(ORGANISM-STATS+CURATED-GENES)&orgids=INKY1) | [432](http://localhost:1555/comp-genomics?type=(ORGANISM-STATS+CURATED-GENES)&orgids=IN366) | [383](http://localhost:1555/comp-genomics?type=(ORGANISM-STATS+CURATED-GENES)&orgids=IN370) |
| [Genes with experimental evidence](http://localhost:1555/comp-genomics?type=(ORGANISM-STATS+GENES-WITH-EXP-EVIDENCE)&orgids=(IN55195+I012T4+INKY1+IN366+IN370)" \t "_blank) | [0](http://localhost:1555/comp-genomics?type=(ORGANISM-STATS+GENES-WITH-EXP-EVIDENCE)&orgids=IN55195) | [0](http://localhost:1555/comp-genomics?type=(ORGANISM-STATS+GENES-WITH-EXP-EVIDENCE)&orgids=I012T4) | [0](http://localhost:1555/comp-genomics?type=(ORGANISM-STATS+GENES-WITH-EXP-EVIDENCE)&orgids=INKY1) | [0](http://localhost:1555/comp-genomics?type=(ORGANISM-STATS+GENES-WITH-EXP-EVIDENCE)&orgids=IN366) | [0](http://localhost:1555/comp-genomics?type=(ORGANISM-STATS+GENES-WITH-EXP-EVIDENCE)&orgids=IN370) |
| [Pseudogenes](http://localhost:1555/comp-genomics?type=(ORGANISM-STATS+PSEUDO-GENES)&orgids=(IN55195+I012T4+INKY1+IN366+IN370)" \t "_blank) | [0](http://localhost:1555/comp-genomics?type=(ORGANISM-STATS+PSEUDO-GENES)&orgids=IN55195) | [0](http://localhost:1555/comp-genomics?type=(ORGANISM-STATS+PSEUDO-GENES)&orgids=I012T4) | [0](http://localhost:1555/comp-genomics?type=(ORGANISM-STATS+PSEUDO-GENES)&orgids=INKY1) | [0](http://localhost:1555/comp-genomics?type=(ORGANISM-STATS+PSEUDO-GENES)&orgids=IN366) | [0](http://localhost:1555/comp-genomics?type=(ORGANISM-STATS+PSEUDO-GENES)&orgids=IN370) |
| [Essential Genes](http://localhost:1555/comp-genomics?type=(ORGANISM-STATS+ESSENTIAL-GENES)&orgids=(IN55195+I012T4+INKY1+IN366+IN370)" \t "_blank) | [0](http://localhost:1555/comp-genomics?type=(ORGANISM-STATS+ESSENTIAL-GENES)&orgids=IN55195) | [0](http://localhost:1555/comp-genomics?type=(ORGANISM-STATS+ESSENTIAL-GENES)&orgids=I012T4) | [0](http://localhost:1555/comp-genomics?type=(ORGANISM-STATS+ESSENTIAL-GENES)&orgids=INKY1) | [0](http://localhost:1555/comp-genomics?type=(ORGANISM-STATS+ESSENTIAL-GENES)&orgids=IN366) | [0](http://localhost:1555/comp-genomics?type=(ORGANISM-STATS+ESSENTIAL-GENES)&orgids=IN370) |
| [%GC Content](http://localhost:1555/comp-genomics?type=(ORGANISM-STATS+GC-CONTENT)&orgids=(IN55195+I012T4+INKY1+IN366+IN370)) | 32.55 | 32.38 | 33.35 | 32.58 | 32.82 |
| [Protein Features](http://localhost:1555/comp-genomics?type=(ORGANISM-STATS+PROTEIN-FEATURES)&orgids=(IN55195+I012T4+INKY1+IN366+IN370)) | [0](http://localhost:1555/comp-genomics?type=(ORGANISM-STATS+PROTEIN-FEATURES)&orgids=IN55195) | [0](http://localhost:1555/comp-genomics?type=(ORGANISM-STATS+PROTEIN-FEATURES)&orgids=I012T4) | [0](http://localhost:1555/comp-genomics?type=(ORGANISM-STATS+PROTEIN-FEATURES)&orgids=INKY1) | [0](http://localhost:1555/comp-genomics?type=(ORGANISM-STATS+PROTEIN-FEATURES)&orgids=IN366) | [0](http://localhost:1555/comp-genomics?type=(ORGANISM-STATS+PROTEIN-FEATURES)&orgids=IN370) |
| [Protein Complexes](http://localhost:1555/comp-genomics?type=(ORGANISM-STATS+COMPLEXES)&orgids=(IN55195+I012T4+INKY1+IN366+IN370)) | [0](http://localhost:1555/comp-genomics?type=(ORGANISM-STATS+COMPLEXES)&orgids=IN55195) | [35](http://localhost:1555/comp-genomics?type=(ORGANISM-STATS+COMPLEXES)&orgids=I012T4) | [24](http://localhost:1555/comp-genomics?type=(ORGANISM-STATS+COMPLEXES)&orgids=INKY1) | [0](http://localhost:1555/comp-genomics?type=(ORGANISM-STATS+COMPLEXES)&orgids=IN366) | [0](http://localhost:1555/comp-genomics?type=(ORGANISM-STATS+COMPLEXES)&orgids=IN370) |
| [Pathways](http://localhost:1555/comp-genomics?type=(ORGANISM-STATS+PATHWAYS)&orgids=(IN55195+I012T4+INKY1+IN366+IN370)) | [114](http://localhost:1555/comp-genomics?type=(ORGANISM-STATS+PATHWAYS)&orgids=IN55195) | [110](http://localhost:1555/comp-genomics?type=(ORGANISM-STATS+PATHWAYS)&orgids=I012T4) | [108](http://localhost:1555/comp-genomics?type=(ORGANISM-STATS+PATHWAYS)&orgids=INKY1) | [118](http://localhost:1555/comp-genomics?type=(ORGANISM-STATS+PATHWAYS)&orgids=IN366) | [105](http://localhost:1555/comp-genomics?type=(ORGANISM-STATS+PATHWAYS)&orgids=IN370) |
| [Pathways with experimental evidence](http://localhost:1555/comp-genomics?type=(ORGANISM-STATS+PATHWAYS-WITH-EXP-EVIDENCE)&orgids=(IN55195+I012T4+INKY1+IN366+IN370)" \t "_blank) | [0](http://localhost:1555/comp-genomics?type=(ORGANISM-STATS+PATHWAYS-WITH-EXP-EVIDENCE)&orgids=IN55195) | [0](http://localhost:1555/comp-genomics?type=(ORGANISM-STATS+PATHWAYS-WITH-EXP-EVIDENCE)&orgids=I012T4) | [0](http://localhost:1555/comp-genomics?type=(ORGANISM-STATS+PATHWAYS-WITH-EXP-EVIDENCE)&orgids=INKY1) | [0](http://localhost:1555/comp-genomics?type=(ORGANISM-STATS+PATHWAYS-WITH-EXP-EVIDENCE)&orgids=IN366) | [0](http://localhost:1555/comp-genomics?type=(ORGANISM-STATS+PATHWAYS-WITH-EXP-EVIDENCE)&orgids=IN370) |
| [Metabolic Reactions](http://localhost:1555/comp-genomics?type=(ORGANISM-STATS+METABOLIC-RXNS)&orgids=(IN55195+I012T4+INKY1+IN366+IN370)) | [616](http://localhost:1555/comp-genomics?type=(ORGANISM-STATS+METABOLIC-RXNS)&orgids=IN55195) | [599](http://localhost:1555/comp-genomics?type=(ORGANISM-STATS+METABOLIC-RXNS)&orgids=I012T4) | [560](http://localhost:1555/comp-genomics?type=(ORGANISM-STATS+METABOLIC-RXNS)&orgids=INKY1) | [634](http://localhost:1555/comp-genomics?type=(ORGANISM-STATS+METABOLIC-RXNS)&orgids=IN366) | [572](http://localhost:1555/comp-genomics?type=(ORGANISM-STATS+METABOLIC-RXNS)&orgids=IN370) |
| [Metabolic Reactions with experimental evidence](http://localhost:1555/comp-genomics?type=(ORGANISM-STATS+REACTIONS-WITH-EXP-EVIDENCE)&orgids=(IN55195+I012T4+INKY1+IN366+IN370)" \t "_blank) | [0](http://localhost:1555/comp-genomics?type=(ORGANISM-STATS+REACTIONS-WITH-EXP-EVIDENCE)&orgids=IN55195) | [0](http://localhost:1555/comp-genomics?type=(ORGANISM-STATS+REACTIONS-WITH-EXP-EVIDENCE)&orgids=I012T4) | [0](http://localhost:1555/comp-genomics?type=(ORGANISM-STATS+REACTIONS-WITH-EXP-EVIDENCE)&orgids=INKY1) | [0](http://localhost:1555/comp-genomics?type=(ORGANISM-STATS+REACTIONS-WITH-EXP-EVIDENCE)&orgids=IN366) | [0](http://localhost:1555/comp-genomics?type=(ORGANISM-STATS+REACTIONS-WITH-EXP-EVIDENCE)&orgids=IN370) |
| [Transport Reactions](http://localhost:1555/comp-genomics?type=(ORGANISM-STATS+TRANSPORT-RXNS)&orgids=(IN55195+I012T4+INKY1+IN366+IN370)) | [38](http://localhost:1555/comp-genomics?type=(ORGANISM-STATS+TRANSPORT-RXNS)&orgids=IN55195) | [62](http://localhost:1555/comp-genomics?type=(ORGANISM-STATS+TRANSPORT-RXNS)&orgids=I012T4) | [30](http://localhost:1555/comp-genomics?type=(ORGANISM-STATS+TRANSPORT-RXNS)&orgids=INKY1) | [37](http://localhost:1555/comp-genomics?type=(ORGANISM-STATS+TRANSPORT-RXNS)&orgids=IN366) | [32](http://localhost:1555/comp-genomics?type=(ORGANISM-STATS+TRANSPORT-RXNS)&orgids=IN370) |
| [Transport Reactions with experimental evidence](http://localhost:1555/comp-genomics?type=(ORGANISM-STATS+TRANSPORT-REACTIONS-WITH-EXP-EVIDENCE)&orgids=(IN55195+I012T4+INKY1+IN366+IN370)" \t "_blank) | [0](http://localhost:1555/comp-genomics?type=(ORGANISM-STATS+TRANSPORT-REACTIONS-WITH-EXP-EVIDENCE)&orgids=IN55195) | [0](http://localhost:1555/comp-genomics?type=(ORGANISM-STATS+TRANSPORT-REACTIONS-WITH-EXP-EVIDENCE)&orgids=I012T4) | [0](http://localhost:1555/comp-genomics?type=(ORGANISM-STATS+TRANSPORT-REACTIONS-WITH-EXP-EVIDENCE)&orgids=INKY1) | [0](http://localhost:1555/comp-genomics?type=(ORGANISM-STATS+TRANSPORT-REACTIONS-WITH-EXP-EVIDENCE)&orgids=IN366) | [0](http://localhost:1555/comp-genomics?type=(ORGANISM-STATS+TRANSPORT-REACTIONS-WITH-EXP-EVIDENCE)&orgids=IN370) |
| [Compounds](http://localhost:1555/comp-genomics?type=(ORGANISM-STATS+COMPOUNDS)&orgids=(IN55195+I012T4+INKY1+IN366+IN370)) | [672](http://localhost:1555/comp-genomics?type=(ORGANISM-STATS+COMPOUNDS)&orgids=IN55195) | [675](http://localhost:1555/comp-genomics?type=(ORGANISM-STATS+COMPOUNDS)&orgids=I012T4) | [636](http://localhost:1555/comp-genomics?type=(ORGANISM-STATS+COMPOUNDS)&orgids=INKY1) | [685](http://localhost:1555/comp-genomics?type=(ORGANISM-STATS+COMPOUNDS)&orgids=IN366) | [645](http://localhost:1555/comp-genomics?type=(ORGANISM-STATS+COMPOUNDS)&orgids=IN370) |
| [Regulatory Interactions](http://localhost:1555/comp-genomics?type=(ORGANISM-STATS+REGULATORY-INTERACTIONS)&orgids=(IN55195+I012T4+INKY1+IN366+IN370)) | [0](http://localhost:1555/comp-genomics?type=(ORGANISM-STATS+REGULATORY-INTERACTIONS)&orgids=IN55195) | [0](http://localhost:1555/comp-genomics?type=(ORGANISM-STATS+REGULATORY-INTERACTIONS)&orgids=I012T4) | [0](http://localhost:1555/comp-genomics?type=(ORGANISM-STATS+REGULATORY-INTERACTIONS)&orgids=INKY1) | [0](http://localhost:1555/comp-genomics?type=(ORGANISM-STATS+REGULATORY-INTERACTIONS)&orgids=IN366) | [0](http://localhost:1555/comp-genomics?type=(ORGANISM-STATS+REGULATORY-INTERACTIONS)&orgids=IN370) |
| [Transcription Units](http://localhost:1555/comp-genomics?type=(ORGANISM-STATS+TRANSCRIPTION-UNITS)&orgids=(IN55195+I012T4+INKY1+IN366+IN370)) | [622](http://localhost:1555/comp-genomics?type=(ORGANISM-STATS+TRANSCRIPTION-UNITS)&orgids=IN55195) | [618](http://localhost:1555/comp-genomics?type=(ORGANISM-STATS+TRANSCRIPTION-UNITS)&orgids=I012T4) | [768](http://localhost:1555/comp-genomics?type=(ORGANISM-STATS+TRANSCRIPTION-UNITS)&orgids=INKY1) | [676](http://localhost:1555/comp-genomics?type=(ORGANISM-STATS+TRANSCRIPTION-UNITS)&orgids=IN366) | [627](http://localhost:1555/comp-genomics?type=(ORGANISM-STATS+TRANSCRIPTION-UNITS)&orgids=IN370) |
| [Promoters](http://localhost:1555/comp-genomics?type=(ORGANISM-STATS+PROMOTERS)&orgids=(IN55195+I012T4+INKY1+IN366+IN370)) | [0](http://localhost:1555/comp-genomics?type=(ORGANISM-STATS+PROMOTERS)&orgids=IN55195) | [0](http://localhost:1555/comp-genomics?type=(ORGANISM-STATS+PROMOTERS)&orgids=I012T4) | [0](http://localhost:1555/comp-genomics?type=(ORGANISM-STATS+PROMOTERS)&orgids=INKY1) | [0](http://localhost:1555/comp-genomics?type=(ORGANISM-STATS+PROMOTERS)&orgids=IN366) | [0](http://localhost:1555/comp-genomics?type=(ORGANISM-STATS+PROMOTERS)&orgids=IN370) |
| [Transcription Factor Binding Sites](http://localhost:1555/comp-genomics?type=(ORGANISM-STATS+BINDING-SITES)&orgids=(IN55195+I012T4+INKY1+IN366+IN370)) | [0](http://localhost:1555/comp-genomics?type=(ORGANISM-STATS+BINDING-SITES)&orgids=IN55195) | [0](http://localhost:1555/comp-genomics?type=(ORGANISM-STATS+BINDING-SITES)&orgids=I012T4) | [0](http://localhost:1555/comp-genomics?type=(ORGANISM-STATS+BINDING-SITES)&orgids=INKY1) | [0](http://localhost:1555/comp-genomics?type=(ORGANISM-STATS+BINDING-SITES)&orgids=IN366) | [0](http://localhost:1555/comp-genomics?type=(ORGANISM-STATS+BINDING-SITES)&orgids=IN370) |
| [Prophages](http://localhost:1555/comp-genomics?type=(ORGANISM-STATS+PROPHAGES)&orgids=(IN55195+I012T4+INKY1+IN366+IN370)) | [0](http://localhost:1555/comp-genomics?type=(ORGANISM-STATS+PROPHAGES)&orgids=IN55195) | [0](http://localhost:1555/comp-genomics?type=(ORGANISM-STATS+PROPHAGES)&orgids=I012T4) | [0](http://localhost:1555/comp-genomics?type=(ORGANISM-STATS+PROPHAGES)&orgids=INKY1) | [0](http://localhost:1555/comp-genomics?type=(ORGANISM-STATS+PROPHAGES)&orgids=IN366) | [0](http://localhost:1555/comp-genomics?type=(ORGANISM-STATS+PROPHAGES)&orgids=IN370) |
| [Cryptic Prophages](http://localhost:1555/comp-genomics?type=(ORGANISM-STATS+CRYPTIC-PROPHAGES)&orgids=(IN55195+I012T4+INKY1+IN366+IN370)) | [0](http://localhost:1555/comp-genomics?type=(ORGANISM-STATS+CRYPTIC-PROPHAGES)&orgids=IN55195) | [0](http://localhost:1555/comp-genomics?type=(ORGANISM-STATS+CRYPTIC-PROPHAGES)&orgids=I012T4) | [0](http://localhost:1555/comp-genomics?type=(ORGANISM-STATS+CRYPTIC-PROPHAGES)&orgids=INKY1) | [0](http://localhost:1555/comp-genomics?type=(ORGANISM-STATS+CRYPTIC-PROPHAGES)&orgids=IN366) | [0](http://localhost:1555/comp-genomics?type=(ORGANISM-STATS+CRYPTIC-PROPHAGES)&orgids=IN370) |
| [REP Elements](http://localhost:1555/comp-genomics?type=(ORGANISM-STATS+REP-ELEMENTS)&orgids=(IN55195+I012T4+INKY1+IN366+IN370)) | [0](http://localhost:1555/comp-genomics?type=(ORGANISM-STATS+REP-ELEMENTS)&orgids=IN55195) | [0](http://localhost:1555/comp-genomics?type=(ORGANISM-STATS+REP-ELEMENTS)&orgids=I012T4) | [0](http://localhost:1555/comp-genomics?type=(ORGANISM-STATS+REP-ELEMENTS)&orgids=INKY1) | [0](http://localhost:1555/comp-genomics?type=(ORGANISM-STATS+REP-ELEMENTS)&orgids=IN366) | [0](http://localhost:1555/comp-genomics?type=(ORGANISM-STATS+REP-ELEMENTS)&orgids=IN370) |
| [Transposons](http://localhost:1555/comp-genomics?type=(ORGANISM-STATS+TRANSPOSONS)&orgids=(IN55195+I012T4+INKY1+IN366+IN370)) | [0](http://localhost:1555/comp-genomics?type=(ORGANISM-STATS+TRANSPOSONS)&orgids=IN55195) | [0](http://localhost:1555/comp-genomics?type=(ORGANISM-STATS+TRANSPOSONS)&orgids=I012T4) | [0](http://localhost:1555/comp-genomics?type=(ORGANISM-STATS+TRANSPOSONS)&orgids=INKY1) | [0](http://localhost:1555/comp-genomics?type=(ORGANISM-STATS+TRANSPOSONS)&orgids=IN366) | [0](http://localhost:1555/comp-genomics?type=(ORGANISM-STATS+TRANSPOSONS)&orgids=IN370) |
| [Phage Attachment Sites](http://localhost:1555/comp-genomics?type=(ORGANISM-STATS+PHAGE-ATTACHMENT-SITES)&orgids=(IN55195+I012T4+INKY1+IN366+IN370)) | [0](http://localhost:1555/comp-genomics?type=(ORGANISM-STATS+PHAGE-ATTACHMENT-SITES)&orgids=IN55195) | [0](http://localhost:1555/comp-genomics?type=(ORGANISM-STATS+PHAGE-ATTACHMENT-SITES)&orgids=I012T4) | [0](http://localhost:1555/comp-genomics?type=(ORGANISM-STATS+PHAGE-ATTACHMENT-SITES)&orgids=INKY1) | [0](http://localhost:1555/comp-genomics?type=(ORGANISM-STATS+PHAGE-ATTACHMENT-SITES)&orgids=IN366) | [0](http://localhost:1555/comp-genomics?type=(ORGANISM-STATS+PHAGE-ATTACHMENT-SITES)&orgids=IN370) |
| [Publications](http://localhost:1555/comp-genomics?type=(ORGANISM-STATS+PUBLICATIONS)&orgids=(IN55195+I012T4+INKY1+IN366+IN370)) | [811](http://localhost:1555/comp-genomics?type=(ORGANISM-STATS+PUBLICATIONS)&orgids=IN55195) | [1,276](http://localhost:1555/comp-genomics?type=(ORGANISM-STATS+PUBLICATIONS)&orgids=I012T4) | [1,285](http://localhost:1555/comp-genomics?type=(ORGANISM-STATS+PUBLICATIONS)&orgids=INKY1) | [814](http://localhost:1555/comp-genomics?type=(ORGANISM-STATS+PUBLICATIONS)&orgids=IN366) | [797](http://localhost:1555/comp-genomics?type=(ORGANISM-STATS+PUBLICATIONS)&orgids=IN370) |
| [Total GO term annotations](http://localhost:1555/comp-genomics?type=(ORGANISM-STATS+GO-TERMS-ANNOTS)&orgids=(IN55195+I012T4+INKY1+IN366+IN370)) | 0 | 0 | 0 | 0 | 0 |

**Reactions**

**Supplemental Table 5B: Breakdown of Reactions by Type**

This table counts reactions based on the types of their substrates. Clicking on any row, column or cell will show the complete list of reactions included in that category.

| [**Reaction Type**](http://localhost:1555/comp-genomics?type=RXN-TYPES&orgids=(IN55195+I012T4+INKY1+IN366+IN370)) | [*L.iners* ATCC 55195](http://localhost:1555/IN55195/organism-summary) | [*L.iners* I012T4](http://localhost:1555/I012T4/organism-summary) | [*L.iners* KY](http://localhost:1555/INKY1/organism-summary) | [*L.iners* pt1](http://localhost:1555/IN366/organism-summary) | [*L.iners* pt2](http://localhost:1555/IN370/organism-summary) |
| --- | --- | --- | --- | --- | --- |
| [Reactions in which all substrates are small molecules](http://localhost:1555/comp-genomics?type=(RXN-TYPES+SMALL-MOLECULE)&orgids=(IN55195+I012T4+INKY1+IN366+IN370)) | [542](http://localhost:1555/comp-genomics?type=(RXN-TYPES+SMALL-MOLECULE)&orgids=IN55195) | [528](http://localhost:1555/comp-genomics?type=(RXN-TYPES+SMALL-MOLECULE)&orgids=I012T4) | [488](http://localhost:1555/comp-genomics?type=(RXN-TYPES+SMALL-MOLECULE)&orgids=INKY1) | [558](http://localhost:1555/comp-genomics?type=(RXN-TYPES+SMALL-MOLECULE)&orgids=IN366) | [501](http://localhost:1555/comp-genomics?type=(RXN-TYPES+SMALL-MOLECULE)&orgids=IN370) |
| [Reactions of proteins with small molecules](http://localhost:1555/comp-genomics?type=(RXN-TYPES+PROTEIN-SMALL-MOLECULE-REACTION)&orgids=(IN55195+I012T4+INKY1+IN366+IN370)) | [78](http://localhost:1555/comp-genomics?type=(RXN-TYPES+PROTEIN-SMALL-MOLECULE-REACTION)&orgids=IN55195) | [73](http://localhost:1555/comp-genomics?type=(RXN-TYPES+PROTEIN-SMALL-MOLECULE-REACTION)&orgids=I012T4) | [73](http://localhost:1555/comp-genomics?type=(RXN-TYPES+PROTEIN-SMALL-MOLECULE-REACTION)&orgids=INKY1) | [78](http://localhost:1555/comp-genomics?type=(RXN-TYPES+PROTEIN-SMALL-MOLECULE-REACTION)&orgids=IN366) | [77](http://localhost:1555/comp-genomics?type=(RXN-TYPES+PROTEIN-SMALL-MOLECULE-REACTION)&orgids=IN370) |
| [Reactions in which all substrates are proteins](http://localhost:1555/comp-genomics?type=(RXN-TYPES+PROTEIN-REACTION)&orgids=(IN55195+I012T4+INKY1+IN366+IN370)) | [6](http://localhost:1555/comp-genomics?type=(RXN-TYPES+PROTEIN-REACTION)&orgids=IN55195) | [6](http://localhost:1555/comp-genomics?type=(RXN-TYPES+PROTEIN-REACTION)&orgids=I012T4) | [7](http://localhost:1555/comp-genomics?type=(RXN-TYPES+PROTEIN-REACTION)&orgids=INKY1) | [6](http://localhost:1555/comp-genomics?type=(RXN-TYPES+PROTEIN-REACTION)&orgids=IN366) | [8](http://localhost:1555/comp-genomics?type=(RXN-TYPES+PROTEIN-REACTION)&orgids=IN370) |
| [Reactions in which one substrate is a tRNA](http://localhost:1555/comp-genomics?type=(RXN-TYPES+TRNA-REACTION)&orgids=(IN55195+I012T4+INKY1+IN366+IN370)) | [6](http://localhost:1555/comp-genomics?type=(RXN-TYPES+TRNA-REACTION)&orgids=IN55195) | [6](http://localhost:1555/comp-genomics?type=(RXN-TYPES+TRNA-REACTION)&orgids=I012T4) | [6](http://localhost:1555/comp-genomics?type=(RXN-TYPES+TRNA-REACTION)&orgids=INKY1) | [6](http://localhost:1555/comp-genomics?type=(RXN-TYPES+TRNA-REACTION)&orgids=IN366) | [6](http://localhost:1555/comp-genomics?type=(RXN-TYPES+TRNA-REACTION)&orgids=IN370) |
| [Transport reactions](http://localhost:1555/comp-genomics?type=(RXN-TYPES+TRANSPORT)&orgids=(IN55195+I012T4+INKY1+IN366+IN370)) | [39](http://localhost:1555/comp-genomics?type=(RXN-TYPES+TRANSPORT)&orgids=IN55195) | [63](http://localhost:1555/comp-genomics?type=(RXN-TYPES+TRANSPORT)&orgids=I012T4) | [31](http://localhost:1555/comp-genomics?type=(RXN-TYPES+TRANSPORT)&orgids=INKY1) | [38](http://localhost:1555/comp-genomics?type=(RXN-TYPES+TRANSPORT)&orgids=IN366) | [33](http://localhost:1555/comp-genomics?type=(RXN-TYPES+TRANSPORT)&orgids=IN370) |
| [Other reactions](http://localhost:1555/comp-genomics?type=(RXN-TYPES+OTHER)&orgids=(IN55195+I012T4+INKY1+IN366+IN370)) | [115](http://localhost:1555/comp-genomics?type=(RXN-TYPES+OTHER)&orgids=IN55195) | [112](http://localhost:1555/comp-genomics?type=(RXN-TYPES+OTHER)&orgids=I012T4) | [112](http://localhost:1555/comp-genomics?type=(RXN-TYPES+OTHER)&orgids=INKY1) | [113](http://localhost:1555/comp-genomics?type=(RXN-TYPES+OTHER)&orgids=IN366) | [110](http://localhost:1555/comp-genomics?type=(RXN-TYPES+OTHER)&orgids=IN370) |
| [Total number of reactions](http://localhost:1555/comp-genomics?type=(RXN-TYPES+ALL)&orgids=(IN55195+I012T4+INKY1+IN366+IN370)) | [786](http://localhost:1555/comp-genomics?type=(RXN-TYPES+ALL)&orgids=IN55195) | [788](http://localhost:1555/comp-genomics?type=(RXN-TYPES+ALL)&orgids=I012T4) | [717](http://localhost:1555/comp-genomics?type=(RXN-TYPES+ALL)&orgids=INKY1) | [799](http://localhost:1555/comp-genomics?type=(RXN-TYPES+ALL)&orgids=IN366) | [735](http://localhost:1555/comp-genomics?type=(RXN-TYPES+ALL)&orgids=IN370) |

**Supplemental Table 5C: Reactions of Small Molecule Metabolism**

For this table, we show statistics for that subset of reactions that can be classified as reactions of small-molecule metabolism (SMM Reactions). We define SMM Reactions to include all reactions in which all substrates are small molecules, as well as all reactions (enzyme-catalyzed or not) involved in metabolic pathways. This category does include some reactions whose substrates are macromolecules (such as reactions of ACP), because these reactions are parts of pathways, but it does not include reactions of macromolecules in general. It includes reactions that are pathway holes (i.e. for which no enzyme has been identified but which are inferred to be present because they are part of a pathway). Transport reactions are not included in this category.

|  | [*L.iners* ATCC 55195](http://localhost:1555/IN55195/organism-summary) | [*L.iners* I012T4](http://localhost:1555/I012T4/organism-summary) | [*L.iners* KY](http://localhost:1555/INKY1/organism-summary) | [*L.iners* pt1](http://localhost:1555/IN366/organism-summary) | [*L.iners* pt2](http://localhost:1555/IN370/organism-summary) |
| --- | --- | --- | --- | --- | --- |
| [Total number of SMM Reactions](http://localhost:1555/comp-genomics?type=(METAB-RXN-STATS+ALL-METAB-RXNS)&orgids=(IN55195+I012T4+INKY1+IN366+IN370)) | [616](http://localhost:1555/comp-genomics?type=(METAB-RXN-STATS+ALL-METAB-RXNS)&orgids=IN55195) | [599](http://localhost:1555/comp-genomics?type=(METAB-RXN-STATS+ALL-METAB-RXNS)&orgids=I012T4) | [560](http://localhost:1555/comp-genomics?type=(METAB-RXN-STATS+ALL-METAB-RXNS)&orgids=INKY1) | [634](http://localhost:1555/comp-genomics?type=(METAB-RXN-STATS+ALL-METAB-RXNS)&orgids=IN366) | [572](http://localhost:1555/comp-genomics?type=(METAB-RXN-STATS+ALL-METAB-RXNS)&orgids=IN370) |
| [SMM Reactions with identified enzymes](http://localhost:1555/comp-genomics?type=(METAB-RXN-STATS+RXNS-W-ENZYMES)&orgids=(IN55195+I012T4+INKY1+IN366+IN370)) | [449](http://localhost:1555/comp-genomics?type=(METAB-RXN-STATS+RXNS-W-ENZYMES)&orgids=IN55195) | [435](http://localhost:1555/comp-genomics?type=(METAB-RXN-STATS+RXNS-W-ENZYMES)&orgids=I012T4) | [415](http://localhost:1555/comp-genomics?type=(METAB-RXN-STATS+RXNS-W-ENZYMES)&orgids=INKY1) | [467](http://localhost:1555/comp-genomics?type=(METAB-RXN-STATS+RXNS-W-ENZYMES)&orgids=IN366) | [421](http://localhost:1555/comp-genomics?type=(METAB-RXN-STATS+RXNS-W-ENZYMES)&orgids=IN370) |
| [SMM Reactions in more than one pathway](http://localhost:1555/comp-genomics?type=(METAB-RXN-STATS+RXNS-IN-MULTIPLE-PWYS)&orgids=(IN55195+I012T4+INKY1+IN366+IN370)) | [44](http://localhost:1555/comp-genomics?type=(METAB-RXN-STATS+RXNS-IN-MULTIPLE-PWYS)&orgids=IN55195) | [40](http://localhost:1555/comp-genomics?type=(METAB-RXN-STATS+RXNS-IN-MULTIPLE-PWYS)&orgids=I012T4) | [42](http://localhost:1555/comp-genomics?type=(METAB-RXN-STATS+RXNS-IN-MULTIPLE-PWYS)&orgids=INKY1) | [45](http://localhost:1555/comp-genomics?type=(METAB-RXN-STATS+RXNS-IN-MULTIPLE-PWYS)&orgids=IN366) | [42](http://localhost:1555/comp-genomics?type=(METAB-RXN-STATS+RXNS-IN-MULTIPLE-PWYS)&orgids=IN370) |

**Supplemental Table 5D: Breakdown of SMM Reactions by Top-Level EC Category**

This table shows the distribution of reactions in the database across the 6 top-level categories identified by the Enzyme Commission. Included in this table are all reactions in the database which have been assigned either full or partial EC numbers, and for which an enzyme has been identified (that is, these statistics do not include pathway holes).

| [**EC Category**](http://localhost:1555/comp-genomics?type=EC-DIST&orgids=(IN55195+I012T4+INKY1+IN366+IN370)) | [*L.iners* ATCC 55195](http://localhost:1555/IN55195/organism-summary) | [*L.iners* I012T4](http://localhost:1555/I012T4/organism-summary) | [*L.iners* KY](http://localhost:1555/INKY1/organism-summary) | [*L.iners* pt1](http://localhost:1555/IN366/organism-summary) | [*L.iners* pt2](http://localhost:1555/IN370/organism-summary) |
| --- | --- | --- | --- | --- | --- |
| [1 -- Oxidoreductases](http://localhost:1555/comp-genomics?type=(EC-DIST+EC-1)&orgids=(IN55195+I012T4+INKY1+IN366+IN370)) | [49 (9%)](http://localhost:1555/comp-genomics?type=(EC-DIST+EC-1)&orgids=IN55195) | [49 (9%)](http://localhost:1555/comp-genomics?type=(EC-DIST+EC-1)&orgids=I012T4) | [52 (10%)](http://localhost:1555/comp-genomics?type=(EC-DIST+EC-1)&orgids=INKY1) | [52 (9%)](http://localhost:1555/comp-genomics?type=(EC-DIST+EC-1)&orgids=IN366) | [52 (10%)](http://localhost:1555/comp-genomics?type=(EC-DIST+EC-1)&orgids=IN370) |
| [2 -- Transferases](http://localhost:1555/comp-genomics?type=(EC-DIST+EC-2)&orgids=(IN55195+I012T4+INKY1+IN366+IN370)) | [192 (36%)](http://localhost:1555/comp-genomics?type=(EC-DIST+EC-2)&orgids=IN55195) | [175 (34%)](http://localhost:1555/comp-genomics?type=(EC-DIST+EC-2)&orgids=I012T4) | [166 (33%)](http://localhost:1555/comp-genomics?type=(EC-DIST+EC-2)&orgids=INKY1) | [191 (35%)](http://localhost:1555/comp-genomics?type=(EC-DIST+EC-2)&orgids=IN366) | [172 (34%)](http://localhost:1555/comp-genomics?type=(EC-DIST+EC-2)&orgids=IN370) |
| [3 -- Hydrolases](http://localhost:1555/comp-genomics?type=(EC-DIST+EC-3)&orgids=(IN55195+I012T4+INKY1+IN366+IN370)) | [181 (34%)](http://localhost:1555/comp-genomics?type=(EC-DIST+EC-3)&orgids=IN55195) | [180 (35%)](http://localhost:1555/comp-genomics?type=(EC-DIST+EC-3)&orgids=I012T4) | [169 (34%)](http://localhost:1555/comp-genomics?type=(EC-DIST+EC-3)&orgids=INKY1) | [190 (34%)](http://localhost:1555/comp-genomics?type=(EC-DIST+EC-3)&orgids=IN366) | [177 (35%)](http://localhost:1555/comp-genomics?type=(EC-DIST+EC-3)&orgids=IN370) |
| [4 -- Lyases](http://localhost:1555/comp-genomics?type=(EC-DIST+EC-4)&orgids=(IN55195+I012T4+INKY1+IN366+IN370)) | [23 (4%)](http://localhost:1555/comp-genomics?type=(EC-DIST+EC-4)&orgids=IN55195) | [22 (4%)](http://localhost:1555/comp-genomics?type=(EC-DIST+EC-4)&orgids=I012T4) | [21 (4%)](http://localhost:1555/comp-genomics?type=(EC-DIST+EC-4)&orgids=INKY1) | [23 (4%)](http://localhost:1555/comp-genomics?type=(EC-DIST+EC-4)&orgids=IN366) | [18 (4%)](http://localhost:1555/comp-genomics?type=(EC-DIST+EC-4)&orgids=IN370) |
| [5 -- Isomerases](http://localhost:1555/comp-genomics?type=(EC-DIST+EC-5)&orgids=(IN55195+I012T4+INKY1+IN366+IN370)) | [35 (6%)](http://localhost:1555/comp-genomics?type=(EC-DIST+EC-5)&orgids=IN55195) | [36 (7%)](http://localhost:1555/comp-genomics?type=(EC-DIST+EC-5)&orgids=I012T4) | [34 (7%)](http://localhost:1555/comp-genomics?type=(EC-DIST+EC-5)&orgids=INKY1) | [36 (7%)](http://localhost:1555/comp-genomics?type=(EC-DIST+EC-5)&orgids=IN366) | [34 (7%)](http://localhost:1555/comp-genomics?type=(EC-DIST+EC-5)&orgids=IN370) |
| [6 -- Ligases](http://localhost:1555/comp-genomics?type=(EC-DIST+EC-6)&orgids=(IN55195+I012T4+INKY1+IN366+IN370)) | [59 (11%)](http://localhost:1555/comp-genomics?type=(EC-DIST+EC-6)&orgids=IN55195) | [57 (11%)](http://localhost:1555/comp-genomics?type=(EC-DIST+EC-6)&orgids=I012T4) | [56 (11%)](http://localhost:1555/comp-genomics?type=(EC-DIST+EC-6)&orgids=INKY1) | [59 (11%)](http://localhost:1555/comp-genomics?type=(EC-DIST+EC-6)&orgids=IN366) | [55 (11%)](http://localhost:1555/comp-genomics?type=(EC-DIST+EC-6)&orgids=IN370) |
| [Total reactions with full or partial EC Numbers](http://localhost:1555/comp-genomics?type=(EC-DIST+TOTAL)&orgids=(IN55195+I012T4+INKY1+IN366+IN370)) | 539 | 519 | 498 | 551 | 508 |

**Supplemental Table 5E: Shared Reactions**

This table counts the reactions that are shared between pairs of organisms. The number in parentheses represent the Jaccard similarity coefficient for the reactions.

| [**Reactions Shared by Organism Pairs**](http://localhost:1555/comp-genomics?type=REACTION-SHARED&orgids=(IN55195+I012T4+INKY1+IN366+IN370)) | [*L.iners* ATCC 55195](http://localhost:1555/IN55195/organism-summary) | [*L.iners* I012T4](http://localhost:1555/I012T4/organism-summary) | [*L.iners* KY](http://localhost:1555/INKY1/organism-summary) | [*L.iners* pt1](http://localhost:1555/IN366/organism-summary) | [*L.iners* pt2](http://localhost:1555/IN370/organism-summary) |
| --- | --- | --- | --- | --- | --- |
| [*Lactobacillus iners* ATCC 55195](http://localhost:1555/comp-genomics?type=(REACTION-SHARED+IN55195)&orgids=(IN55195+I012T4+INKY1+IN366+IN370)) | [786 (1.000)](http://localhost:1555/comp-genomics?type=(REACTION-SHARED+IN55195)&orgids=IN55195) | [718 (0.839)](http://localhost:1555/comp-genomics?type=(REACTION-SHARED+IN55195)&orgids=(IN55195+I012T4)) | [693 (0.856)](http://localhost:1555/comp-genomics?type=(REACTION-SHARED+IN55195)&orgids=(IN55195+INKY1)) | [777 (0.962)](http://localhost:1555/comp-genomics?type=(REACTION-SHARED+IN55195)&orgids=(IN55195+IN366)) | [718 (0.894)](http://localhost:1555/comp-genomics?type=(REACTION-SHARED+IN55195)&orgids=(IN55195+IN370)) |
| [*Lactobacillus iners* I012T4](http://localhost:1555/comp-genomics?type=(REACTION-SHARED+I012T4)&orgids=(IN55195+I012T4+INKY1+IN366+IN370)) | [718 (0.839)](http://localhost:1555/comp-genomics?type=(REACTION-SHARED+I012T4)&orgids=(I012T4+IN55195)) | [788 (1.000)](http://localhost:1555/comp-genomics?type=(REACTION-SHARED+I012T4)&orgids=I012T4) | [700 (0.870)](http://localhost:1555/comp-genomics?type=(REACTION-SHARED+I012T4)&orgids=(I012T4+INKY1)) | [719 (0.828)](http://localhost:1555/comp-genomics?type=(REACTION-SHARED+I012T4)&orgids=(I012T4+IN366)) | [664 (0.773)](http://localhost:1555/comp-genomics?type=(REACTION-SHARED+I012T4)&orgids=(I012T4+IN370)) |
| [*Lactobacillus iners KY*](http://localhost:1555/comp-genomics?type=(REACTION-SHARED+INKY1)&orgids=(IN55195+I012T4+INKY1+IN366+IN370)) | [693 (0.856)](http://localhost:1555/comp-genomics?type=(REACTION-SHARED+INKY1)&orgids=(INKY1+IN55195)) | [700 (0.870)](http://localhost:1555/comp-genomics?type=(REACTION-SHARED+INKY1)&orgids=(INKY1+I012T4)) | [717 (1.000)](http://localhost:1555/comp-genomics?type=(REACTION-SHARED+INKY1)&orgids=INKY1) | [693 (0.842)](http://localhost:1555/comp-genomics?type=(REACTION-SHARED+INKY1)&orgids=(INKY1+IN366)) | [645 (0.799)](http://localhost:1555/comp-genomics?type=(REACTION-SHARED+INKY1)&orgids=(INKY1+IN370)) |
| [*Lactobacillus iners* pt1](http://localhost:1555/comp-genomics?type=(REACTION-SHARED+IN366)&orgids=(IN55195+I012T4+INKY1+IN366+IN370)) | [777 (0.962)](http://localhost:1555/comp-genomics?type=(REACTION-SHARED+IN366)&orgids=(IN366+IN55195)) | [719 (0.828)](http://localhost:1555/comp-genomics?type=(REACTION-SHARED+IN366)&orgids=(IN366+I012T4)) | [693 (0.842)](http://localhost:1555/comp-genomics?type=(REACTION-SHARED+IN366)&orgids=(IN366+INKY1)) | [799 (1.000)](http://localhost:1555/comp-genomics?type=(REACTION-SHARED+IN366)&orgids=IN366) | [728 (0.903)](http://localhost:1555/comp-genomics?type=(REACTION-SHARED+IN366)&orgids=(IN366+IN370)) |
| [*Lactobacillus iners* pt2](http://localhost:1555/comp-genomics?type=(REACTION-SHARED+IN370)&orgids=(IN55195+I012T4+INKY1+IN366+IN370)) | [718 (0.894)](http://localhost:1555/comp-genomics?type=(REACTION-SHARED+IN370)&orgids=(IN370+IN55195)) | [664 (0.773)](http://localhost:1555/comp-genomics?type=(REACTION-SHARED+IN370)&orgids=(IN370+I012T4)) | [645 (0.799)](http://localhost:1555/comp-genomics?type=(REACTION-SHARED+IN370)&orgids=(IN370+INKY1)) | [728 (0.903)](http://localhost:1555/comp-genomics?type=(REACTION-SHARED+IN370)&orgids=(IN370+IN366)) | [735 (1.000)](http://localhost:1555/comp-genomics?type=(REACTION-SHARED+IN370)&orgids=IN370) |

**Supplemental Table 5F: Unique Reactions**

| **Unique Reactions in Organism**: Unique Reactions | *L.iners* ATCC 55195 | *L.iners* I012T4 | *L.iners* KY | *L.iners* pt1 | *L.iners* pt2 |
| --- | --- | --- | --- | --- | --- |
| [α-D-galactose 6-phosphate ↔ β-D-galactose 6-phosphate](http://localhost:1555/compare-frame-in-orgs?type=NIL&object=RXN-14816&orgids=(IN55195+I012T4+INKY1+IN366+IN370)) | [✓](http://localhost:1555/IN55195/NEW-IMAGE?type=REACTION&object=RXN-14816&orgids=(IN55195%20I012T4%20INKY1%20IN366%20IN370)) |  |  |  |  |
| [2 L-ascorbate + an oxidized unknown electron carrier + 2 H^+^ → 2 monodehydroascorbate radical + a reduced unknown two electron carrier](http://localhost:1555/compare-frame-in-orgs?type=NIL&object=RXN-10981&orgids=(IN55195+I012T4+INKY1+IN366+IN370)) |  | [✓](http://localhost:1555/I012T4/NEW-IMAGE?type=REACTION&object=RXN-10981&orgids=(IN55195%20I012T4%20INKY1%20IN366%20IN370)) |  |  |  |
| [2 L-ascorbate + hydrogen peroxide → L-ascorbate + dehydroascorbate + 2 H_2_O](http://localhost:1555/compare-frame-in-orgs?type=NIL&object=RXN-12440&orgids=(IN55195+I012T4+INKY1+IN366+IN370)) |  | [✓](http://localhost:1555/I012T4/NEW-IMAGE?type=REACTION&object=RXN-12440&orgids=(IN55195%20I012T4%20INKY1%20IN366%20IN370)) |  |  |  |
| [2 L-ascorbate + hydrogen peroxide + 2 H^+^ → 2 monodehydroascorbate radical + 2 H_2_O](http://localhost:1555/compare-frame-in-orgs?type=NIL&object=RXN-3521&orgids=(IN55195+I012T4+INKY1+IN366+IN370)) |  | [✓](http://localhost:1555/I012T4/NEW-IMAGE?type=REACTION&object=RXN-3521&orgids=(IN55195%20I012T4%20INKY1%20IN366%20IN370)) |  |  |  |
| [2 monodehydroascorbate radical → L-ascorbate + dehydroascorbate + 2 H^+^](http://localhost:1555/compare-frame-in-orgs?type=NIL&object=RXN-3523&orgids=(IN55195+I012T4+INKY1+IN366+IN370)) |  | [✓](http://localhost:1555/I012T4/NEW-IMAGE?type=REACTION&object=RXN-3523&orgids=(IN55195%20I012T4%20INKY1%20IN366%20IN370)) |  |  |  |
| [*N*-(5-phosphoribosyl)-anthranilate + diphosphate = anthranilate + 5-phospho-α-D-ribose 1-diphosphate](http://localhost:1555/compare-frame-in-orgs?type=NIL&object=PRTRANS-RXN&orgids=(IN55195+I012T4+INKY1+IN366+IN370)) |  | [✓](http://localhost:1555/I012T4/NEW-IMAGE?type=REACTION&object=PRTRANS-RXN&orgids=(IN55195%20I012T4%20INKY1%20IN366%20IN370)) |  |  |  |
| [*S*-adenosyl-L-methionine + a 5-(aminomethyl)-2-thiouridine^34^ in tRNA → *S*-adenosyl-L-homocysteine + a 5-[(methylamino)methyl]-2-thiouridine^34^ in tRNA + H^+^](http://localhost:1555/compare-frame-in-orgs?type=NIL&object=RXN0-5144&orgids=(IN55195+I012T4+INKY1+IN366+IN370)) |  | [✓](http://localhost:1555/I012T4/NEW-IMAGE?type=REACTION&object=RXN0-5144&orgids=(IN55195%20I012T4%20INKY1%20IN366%20IN370)) |  |  |  |
| [*sn*-glycerol 3-phosphate + dioxygen → hydrogen peroxide + glycerone phosphate](http://localhost:1555/compare-frame-in-orgs?type=NIL&object=GLYCEROL-3-PHOSPHATE-OXIDASE-RXN&orgids=(IN55195+I012T4+INKY1+IN366+IN370)) |  |  |  | [✓](http://localhost:1555/IN366/NEW-IMAGE?type=REACTION&object=GLYCEROL-3-PHOSPHATE-OXIDASE-RXN&orgids=(IN55195%20I012T4%20INKY1%20IN366%20IN370)) |  |
| [*sn*-glycerol 3-phosphate_[in]_ + a menaquinone_[membrane]_ → glycerone phosphate_[in]_ + a menaquinol_[membrane]_](http://localhost:1555/compare-frame-in-orgs?type=NIL&object=RXN-15740&orgids=(IN55195+I012T4+INKY1+IN366+IN370)) |  |  |  | [✓](http://localhost:1555/IN366/NEW-IMAGE?type=REACTION&object=RXN-15740&orgids=(IN55195%20I012T4%20INKY1%20IN366%20IN370)) |  |
| [a 2-thiouridine^34^ in tRNA + GTP + a 5,10-methylenetetrahydrofolate + glycine + an oxidized unknown electron carrier + H_2_O → a 5-carboxymethylaminomethyl-2-thiouridine^34^ in tRNA + GDP + a 7,8-dihydrofolate + a reduced unknown two electron carrier + phosphate + 2 H^+^](http://localhost:1555/compare-frame-in-orgs?type=NIL&object=RXN-18710&orgids=(IN55195+I012T4+INKY1+IN366+IN370)) |  | [✓](http://localhost:1555/I012T4/NEW-IMAGE?type=REACTION&object=RXN-18710&orgids=(IN55195%20I012T4%20INKY1%20IN366%20IN370)) |  |  |  |
| [a 5-carboxymethylaminomethyl-2-thiouridine^34^ in tRNA + FAD + H_2_O + 2 H^+^ → a 5-(aminomethyl)-2-thiouridine^34^ in tRNA + glyoxylate + FADH_2_](http://localhost:1555/compare-frame-in-orgs?type=NIL&object=RXN0-7081&orgids=(IN55195+I012T4+INKY1+IN366+IN370)) |  | [✓](http://localhost:1555/I012T4/NEW-IMAGE?type=REACTION&object=RXN0-7081&orgids=(IN55195%20I012T4%20INKY1%20IN366%20IN370)) |  |  |  |
| [a 5-methylaminomethyl-2-(*Se*-phospho)selenouridine^34^ in tRNA + H_2_O → a 5-methylaminomethyl-2-selenouridine^34^ in tRNA + phosphate](http://localhost:1555/compare-frame-in-orgs?type=NIL&object=RXN-20757&orgids=(IN55195+I012T4+INKY1+IN366+IN370)) |  | [✓](http://localhost:1555/I012T4/NEW-IMAGE?type=REACTION&object=RXN-20757&orgids=(IN55195%20I012T4%20INKY1%20IN366%20IN370)) |  |  |  |
| [a branched-chain amino acid_[extracellular space]_ + H^+^_[extracellular space]_ → a branched-chain amino acid_[cytosol]_ + H^+^_[cytosol]_](http://localhost:1555/compare-frame-in-orgs?type=NIL&object=TRANS-RXN1-7&orgids=(IN55195+I012T4+INKY1+IN366+IN370)) |  | [✓](http://localhost:1555/I012T4/NEW-IMAGE?type=REACTION&object=TRANS-RXN1-7&orgids=(IN55195%20I012T4%20INKY1%20IN366%20IN370)) |  |  |  |
| [a dipeptide_[extracellular space]_ + a tripeptide_[extracellular space]_ → a dipeptide_[cytosol]_ + a tripeptide_[cytosol]_](http://localhost:1555/compare-frame-in-orgs?type=NIL&object=TRANS-RXN1-12&orgids=(IN55195+I012T4+INKY1+IN366+IN370)) |  | [✓](http://localhost:1555/I012T4/NEW-IMAGE?type=REACTION&object=TRANS-RXN1-12&orgids=(IN55195%20I012T4%20INKY1%20IN366%20IN370)) |  |  |  |
| [a metal cation_[extracellular space]_ → a metal cation_[cytosol]_](http://localhost:1555/compare-frame-in-orgs?type=NIL&object=TRANS-RXN1-4&orgids=(IN55195+I012T4+INKY1+IN366+IN370)) |  | [✓](http://localhost:1555/I012T4/NEW-IMAGE?type=REACTION&object=TRANS-RXN1-4&orgids=(IN55195%20I012T4%20INKY1%20IN366%20IN370)) |  |  |  |
| [a metal cation_[extracellular space]_ + ATP + H_2_O → a metal cation_[cytosol]_ + ADP + phosphate + H^+^](http://localhost:1555/compare-frame-in-orgs?type=NIL&object=RXN1-1&orgids=(IN55195+I012T4+INKY1+IN366+IN370)) |  | [✓](http://localhost:1555/I012T4/NEW-IMAGE?type=REACTION&object=RXN1-1&orgids=(IN55195%20I012T4%20INKY1%20IN366%20IN370)) |  |  |  |
| [a peptide_[extracellular space]_ + ATP + H_2_O → a peptide_[cytosol]_ + ADP + phosphate + H^+^](http://localhost:1555/compare-frame-in-orgs?type=NIL&object=RXN1-8&orgids=(IN55195+I012T4+INKY1+IN366+IN370)) |  | [✓](http://localhost:1555/I012T4/NEW-IMAGE?type=REACTION&object=RXN1-8&orgids=(IN55195%20I012T4%20INKY1%20IN366%20IN370)) |  |  |  |
| [a polysaccharide_[extracellular space]_ → a polysaccharide_[cytosol]_](http://localhost:1555/compare-frame-in-orgs?type=NIL&object=TRANS-RXN1-5&orgids=(IN55195+I012T4+INKY1+IN366+IN370)) |  | [✓](http://localhost:1555/I012T4/NEW-IMAGE?type=REACTION&object=TRANS-RXN1-5&orgids=(IN55195%20I012T4%20INKY1%20IN366%20IN370)) |  |  |  |
| [a purine_[extracellular space]_ → a purine_[cytosol]_](http://localhost:1555/compare-frame-in-orgs?type=NIL&object=TRANS-RXN1-3&orgids=(IN55195+I012T4+INKY1+IN366+IN370)) |  | [✓](http://localhost:1555/I012T4/NEW-IMAGE?type=REACTION&object=TRANS-RXN1-3&orgids=(IN55195%20I012T4%20INKY1%20IN366%20IN370)) |  |  |  |
| [a sugar_[extracellular space]_ → a sugar_[cytosol]_](http://localhost:1555/compare-frame-in-orgs?type=NIL&object=TRANS-RXN1-6&orgids=(IN55195+I012T4+INKY1+IN366+IN370)) |  | [✓](http://localhost:1555/I012T4/NEW-IMAGE?type=REACTION&object=TRANS-RXN1-6&orgids=(IN55195%20I012T4%20INKY1%20IN366%20IN370)) |  |  |  |
| [a sugar_[extracellular space]_ + ATP + H_2_O → a sugar_[cytosol]_ + ADP + phosphate + H^+^](http://localhost:1555/compare-frame-in-orgs?type=NIL&object=RXN1-2&orgids=(IN55195+I012T4+INKY1+IN366+IN370)) |  | [✓](http://localhost:1555/I012T4/NEW-IMAGE?type=REACTION&object=RXN1-2&orgids=(IN55195%20I012T4%20INKY1%20IN366%20IN370)) |  |  |  |
| [a [TusA sulfur-carrier protein]-S-sulfanyl-L-cysteine + a [TusD]-L-cysteine → a [TusA]-L-cysteine + a [TusD sulfur-carrier protein]-S-sulfanyl-L-cysteine](http://localhost:1555/compare-frame-in-orgs?type=NIL&object=RXN-18709&orgids=(IN55195+I012T4+INKY1+IN366+IN370)) |  | [✓](http://localhost:1555/I012T4/NEW-IMAGE?type=REACTION&object=RXN-18709&orgids=(IN55195%20I012T4%20INKY1%20IN366%20IN370)) |  |  |  |
| [a [TusD sulfur-carrier protein]-S-sulfanyl-L-cysteine + a [TusE sulfur carrier protein]-L-cysteine → a [TusD]-L-cysteine + a [TusE sulfur carrier protein]-*S*-sulfanylcysteine](http://localhost:1555/compare-frame-in-orgs?type=NIL&object=RXN-18708&orgids=(IN55195+I012T4+INKY1+IN366+IN370)) |  | [✓](http://localhost:1555/I012T4/NEW-IMAGE?type=REACTION&object=RXN-18708&orgids=(IN55195%20I012T4%20INKY1%20IN366%20IN370)) |  |  |  |
| [an amino acid_[extracellular space]_ + ATP + H_2_O → an amino acid_[cytosol]_ + ADP + phosphate + H^+^](http://localhost:1555/compare-frame-in-orgs?type=NIL&object=RXN1-4&orgids=(IN55195+I012T4+INKY1+IN366+IN370)) |  | [✓](http://localhost:1555/I012T4/NEW-IMAGE?type=REACTION&object=RXN1-4&orgids=(IN55195%20I012T4%20INKY1%20IN366%20IN370)) |  |  |  |
| [an amino acid_[extracellular space]_ + H^+^_[extracellular space]_ → an amino acid_[cytosol]_ + H^+^_[cytosol]_](http://localhost:1555/compare-frame-in-orgs?type=NIL&object=TRANS-RXN1-1&orgids=(IN55195+I012T4+INKY1+IN366+IN370)) |  | [✓](http://localhost:1555/I012T4/NEW-IMAGE?type=REACTION&object=TRANS-RXN1-1&orgids=(IN55195%20I012T4%20INKY1%20IN366%20IN370)) |  |  |  |
| [an oligopeptide_[extracellular space]_ + ATP + H_2_O → an oligopeptide_[cytosol]_ + ADP + phosphate + H^+^](http://localhost:1555/compare-frame-in-orgs?type=NIL&object=3.6.3.23-RXN&orgids=(IN55195+I012T4+INKY1+IN366+IN370)) |  | [✓](http://localhost:1555/I012T4/NEW-IMAGE?type=REACTION&object=3.6.3.23-RXN&orgids=(IN55195%20I012T4%20INKY1%20IN366%20IN370)) |  |  |  |
| [an [HPr protein]-*N^π^*-phospho-L-histidine + α-D-galactopyranose_[out]_ → α-D-galactose 6-phosphate_[in]_ + an [HPr]-L-histidine](http://localhost:1555/compare-frame-in-orgs?type=NIL&object=RXN-15170&orgids=(IN55195+I012T4+INKY1+IN366+IN370)) | [✓](http://localhost:1555/IN55195/NEW-IMAGE?type=REACTION&object=RXN-15170&orgids=(IN55195%20I012T4%20INKY1%20IN366%20IN370)) |  |  |  |  |
| [an [HPr protein]-*N^π^*-phospho-L-histidine + β-D-galactopyranose_[out]_ → β-D-galactose 6-phosphate_[in]_ + an [HPr]-L-histidine](http://localhost:1555/compare-frame-in-orgs?type=NIL&object=RXN-15171&orgids=(IN55195+I012T4+INKY1+IN366+IN370)) | [✓](http://localhost:1555/IN55195/NEW-IMAGE?type=REACTION&object=RXN-15171&orgids=(IN55195%20I012T4%20INKY1%20IN366%20IN370)) |  |  |  |  |
| [an [HPr protein]-*N^π^*-phospho-L-histidine + *aldehydo*-D-galactose_[out]_ → *aldehydo*-D-galactose 6-phosphate_[in]_ + an [HPr]-L-histidine](http://localhost:1555/compare-frame-in-orgs?type=NIL&object=RXN-15168&orgids=(IN55195+I012T4+INKY1+IN366+IN370)) | [✓](http://localhost:1555/IN55195/NEW-IMAGE?type=REACTION&object=RXN-15168&orgids=(IN55195%20I012T4%20INKY1%20IN366%20IN370)) |  |  |  |  |
| [an [HPr protein]-*N^π^*-phospho-L-histidine + D-fructofuranose_[out]_ → D-fructofuranose 1-phosphate_[in]_ + an [HPr]-L-histidine](http://localhost:1555/compare-frame-in-orgs?type=NIL&object=RXN-15151&orgids=(IN55195+I012T4+INKY1+IN366+IN370)) |  |  | [✓](http://localhost:1555/INKY1/NEW-IMAGE?type=REACTION&object=RXN-15151&orgids=(IN55195%20I012T4%20INKY1%20IN366%20IN370)) |  |  |
| [an [HPr protein]-*N^π^*-phospho-L-histidine + D-fructopyranose_[out]_ → D-fructopyranose 1-phosphate_[in]_ + an [HPr]-L-histidine](http://localhost:1555/compare-frame-in-orgs?type=NIL&object=RXN-15155&orgids=(IN55195+I012T4+INKY1+IN366+IN370)) |  |  | [✓](http://localhost:1555/INKY1/NEW-IMAGE?type=REACTION&object=RXN-15155&orgids=(IN55195%20I012T4%20INKY1%20IN366%20IN370)) |  |  |
| [an [HPr protein]-*N^π^*-phospho-L-histidine + D-fructose_[extracellular space]_ → D-fructose 1-phosphate_[cytosol]_ + an [HPr]-L-histidine](http://localhost:1555/compare-frame-in-orgs?type=NIL&object=RXN-15084&orgids=(IN55195+I012T4+INKY1+IN366+IN370)) |  |  | [✓](http://localhost:1555/INKY1/NEW-IMAGE?type=REACTION&object=RXN-15084&orgids=(IN55195%20I012T4%20INKY1%20IN366%20IN370)) |  |  |
| [an [HPr protein]-*N^π^*-phospho-L-histidine + D-galactopyranose_[out]_ → D-galactopyranose 6-phosphate_[in]_ + an [HPr]-L-histidine](http://localhost:1555/compare-frame-in-orgs?type=NIL&object=RXN-15169&orgids=(IN55195+I012T4+INKY1+IN366+IN370)) | [✓](http://localhost:1555/IN55195/NEW-IMAGE?type=REACTION&object=RXN-15169&orgids=(IN55195%20I012T4%20INKY1%20IN366%20IN370)) |  |  |  |  |
| [an [HPr protein]-*N^π^*-phospho-L-histidine + D-galactose_[out]_ → a D-galactose 6-phosphate_[in]_ + an [HPr]-L-histidine](http://localhost:1555/compare-frame-in-orgs?type=NIL&object=RXN-15093&orgids=(IN55195+I012T4+INKY1+IN366+IN370)) | [✓](http://localhost:1555/IN55195/NEW-IMAGE?type=REACTION&object=RXN-15093&orgids=(IN55195%20I012T4%20INKY1%20IN366%20IN370)) |  |  |  |  |
| [an [HPr protein]-*N^π^*-phospho-L-histidine + D-glucose_[extracellular space]_ → D-glucose 6-phosphate_[cytosol]_ + an [HPr]-L-histidine](http://localhost:1555/compare-frame-in-orgs?type=NIL&object=RXN-15083&orgids=(IN55195+I012T4+INKY1+IN366+IN370)) |  | [✓](http://localhost:1555/I012T4/NEW-IMAGE?type=REACTION&object=RXN-15083&orgids=(IN55195%20I012T4%20INKY1%20IN366%20IN370)) |  |  |  |
| [an [L-cysteine desulfurase]-*S*-sulfanyl-L-cysteine + a [TusA]-L-cysteine → an [L-cysteine desulfurase]-L-cysteine + a [TusA sulfur-carrier protein]-S-sulfanyl-L-cysteine](http://localhost:1555/compare-frame-in-orgs?type=NIL&object=RXN-18707&orgids=(IN55195+I012T4+INKY1+IN366+IN370)) |  | [✓](http://localhost:1555/I012T4/NEW-IMAGE?type=REACTION&object=RXN-18707&orgids=(IN55195%20I012T4%20INKY1%20IN366%20IN370)) |  |  |  |
| [ATP + *sn*-glycerol 3-phosphate_[extracellular space]_ + H_2_O → ADP + *sn*-glycerol 3-phosphate_[cytosol]_ + phosphate + H^+^](http://localhost:1555/compare-frame-in-orgs?type=NIL&object=ABC-34-RXN&orgids=(IN55195+I012T4+INKY1+IN366+IN370)) |  | [✓](http://localhost:1555/I012T4/NEW-IMAGE?type=REACTION&object=ABC-34-RXN&orgids=(IN55195%20I012T4%20INKY1%20IN366%20IN370)) |  |  |  |
| [ATP + L-methionine_[extracellular space]_ + H_2_O → ADP + L-methionine_[cytosol]_ + phosphate + H^+^](http://localhost:1555/compare-frame-in-orgs?type=NIL&object=RXN0-4522&orgids=(IN55195+I012T4+INKY1+IN366+IN370)) |  | [✓](http://localhost:1555/I012T4/NEW-IMAGE?type=REACTION&object=RXN0-4522&orgids=(IN55195%20I012T4%20INKY1%20IN366%20IN370)) |  |  |  |
| [ATP + Mn^2+^_[extracellular space]_ + H_2_O → ADP + Mn^2+^_[cytosol]_ + phosphate + H^+^](http://localhost:1555/compare-frame-in-orgs?type=NIL&object=3.6.3.35-RXN&orgids=(IN55195+I012T4+INKY1+IN366+IN370)) |  | [✓](http://localhost:1555/I012T4/NEW-IMAGE?type=REACTION&object=3.6.3.35-RXN&orgids=(IN55195%20I012T4%20INKY1%20IN366%20IN370)) |  |  |  |
| [ATP + phosphate_[extracellular space]_ + H_2_O → ADP + phosphate_[cytosol]_ + phosphate_[cytosol]_ + H^+^](http://localhost:1555/compare-frame-in-orgs?type=NIL&object=RXN1-9&orgids=(IN55195+I012T4+INKY1+IN366+IN370)) |  | [✓](http://localhost:1555/I012T4/NEW-IMAGE?type=REACTION&object=RXN1-9&orgids=(IN55195%20I012T4%20INKY1%20IN366%20IN370)) |  |  |  |
| [chloride_[extracellular space]_ → chloride_[cytosol]_](http://localhost:1555/compare-frame-in-orgs?type=NIL&object=TRANS-RXN1-13&orgids=(IN55195+I012T4+INKY1+IN366+IN370)) |  | [✓](http://localhost:1555/I012T4/NEW-IMAGE?type=REACTION&object=TRANS-RXN1-13&orgids=(IN55195%20I012T4%20INKY1%20IN366%20IN370)) |  |  |  |
| [choline_[extracellular space]_ + ATP + H_2_O → choline_[cytosol]_ + ADP + phosphate + H^+^](http://localhost:1555/compare-frame-in-orgs?type=NIL&object=RXN1-6&orgids=(IN55195+I012T4+INKY1+IN366+IN370)) |  | [✓](http://localhost:1555/I012T4/NEW-IMAGE?type=REACTION&object=RXN1-6&orgids=(IN55195%20I012T4%20INKY1%20IN366%20IN370)) |  |  |  |
| [Co^2+^_[extracellular space]_ + ATP + H_2_O → Co^2+^_[cytosol]_ + ADP + phosphate + H^+^](http://localhost:1555/compare-frame-in-orgs?type=NIL&object=RXN1-10&orgids=(IN55195+I012T4+INKY1+IN366+IN370)) |  | [✓](http://localhost:1555/I012T4/NEW-IMAGE?type=REACTION&object=RXN1-10&orgids=(IN55195%20I012T4%20INKY1%20IN366%20IN370)) |  |  |  |
| [cytosine_[extracellular space]_ → cytosine_[cytosol]_](http://localhost:1555/compare-frame-in-orgs?type=NIL&object=TRANS-RXN1-2&orgids=(IN55195+I012T4+INKY1+IN366+IN370)) |  | [✓](http://localhost:1555/I012T4/NEW-IMAGE?type=REACTION&object=TRANS-RXN1-2&orgids=(IN55195%20I012T4%20INKY1%20IN366%20IN370)) |  |  |  |
| [D-xylose_[extracellular space]_ → D-xylose_[cytosol]_](http://localhost:1555/compare-frame-in-orgs?type=NIL&object=TRANS-RXN1-11&orgids=(IN55195+I012T4+INKY1+IN366+IN370)) |  | [✓](http://localhost:1555/I012T4/NEW-IMAGE?type=REACTION&object=TRANS-RXN1-11&orgids=(IN55195%20I012T4%20INKY1%20IN366%20IN370)) |  |  |  |
| [dehydroascorbate (bicyclic form) → 2,3-didehydro-L-gulonate + H^+^](http://localhost:1555/compare-frame-in-orgs?type=NIL&object=RXN-12861&orgids=(IN55195+I012T4+INKY1+IN366+IN370)) |  | [✓](http://localhost:1555/I012T4/NEW-IMAGE?type=REACTION&object=RXN-12861&orgids=(IN55195%20I012T4%20INKY1%20IN366%20IN370)) |  |  |  |
| [dehydroascorbate + H_2_O → 2,3-didehydro-L-gulonate](http://localhost:1555/compare-frame-in-orgs?type=NIL&object=RXN-21400&orgids=(IN55195+I012T4+INKY1+IN366+IN370)) |  | [✓](http://localhost:1555/I012T4/NEW-IMAGE?type=REACTION&object=RXN-21400&orgids=(IN55195%20I012T4%20INKY1%20IN366%20IN370)) |  |  |  |
| [dehydroascorbate + H_2_O + H^+^ → dehydroascorbate (bicyclic form)](http://localhost:1555/compare-frame-in-orgs?type=NIL&object=RXN-12862&orgids=(IN55195+I012T4+INKY1+IN366+IN370)) |  | [✓](http://localhost:1555/I012T4/NEW-IMAGE?type=REACTION&object=RXN-12862&orgids=(IN55195%20I012T4%20INKY1%20IN366%20IN370)) |  |  |  |
| [geranyl diphosphate + a 5-[(methylamino)methyl]-2-thiouridine^34^ in tRNA → a 5-methylaminomethyl-2-(*S*-geranyl)thiouridine^34^ in tRNA + diphosphate](http://localhost:1555/compare-frame-in-orgs?type=NIL&object=RXN-20755&orgids=(IN55195+I012T4+INKY1+IN366+IN370)) |  | [✓](http://localhost:1555/I012T4/NEW-IMAGE?type=REACTION&object=RXN-20755&orgids=(IN55195%20I012T4%20INKY1%20IN366%20IN370)) |  |  |  |
| [glutamate_[extracellular space]_ + 4-aminobutanoate_[cytosol]_ → glutamate_[cytosol]_ + 4-aminobutanoate_[extracellular space]_](http://localhost:1555/compare-frame-in-orgs?type=NIL&object=TRANS-RXN1-10&orgids=(IN55195+I012T4+INKY1+IN366+IN370)) |  | [✓](http://localhost:1555/I012T4/NEW-IMAGE?type=REACTION&object=TRANS-RXN1-10&orgids=(IN55195%20I012T4%20INKY1%20IN366%20IN370)) |  |  |  |
| [glycerol + ATP → *sn*-glycerol 3-phosphate + ADP + H^+^](http://localhost:1555/compare-frame-in-orgs?type=NIL&object=GLYCEROL-KIN-RXN&orgids=(IN55195+I012T4+INKY1+IN366+IN370)) |  |  |  | [✓](http://localhost:1555/IN366/NEW-IMAGE?type=REACTION&object=GLYCEROL-KIN-RXN&orgids=(IN55195%20I012T4%20INKY1%20IN366%20IN370)) |  |
| [glycerol_[extracellular space]_ + H^+^_[extracellular space]_ → glycerol_[cytosol]_ + H^+^_[cytosol]_](http://localhost:1555/compare-frame-in-orgs?type=NIL&object=TRANS-RXN1-8&orgids=(IN55195+I012T4+INKY1+IN366+IN370)) |  | [✓](http://localhost:1555/I012T4/NEW-IMAGE?type=REACTION&object=TRANS-RXN1-8&orgids=(IN55195%20I012T4%20INKY1%20IN366%20IN370)) |  |  |  |
| [glycerol_[out]_ → glycerol_[in]_](http://localhost:1555/compare-frame-in-orgs?type=NIL&object=TRANS-RXN-131&orgids=(IN55195+I012T4+INKY1+IN366+IN370)) |  |  |  | [✓](http://localhost:1555/IN366/NEW-IMAGE?type=REACTION&object=TRANS-RXN-131&orgids=(IN55195%20I012T4%20INKY1%20IN366%20IN370)) |  |
| [glycine betaine_[extracellular space]_ + ATP + H_2_O → glycine betaine_[cytosol]_ + ADP + phosphate + H^+^](http://localhost:1555/compare-frame-in-orgs?type=NIL&object=TRANS-RXN-283&orgids=(IN55195+I012T4+INKY1+IN366+IN370)) |  | [✓](http://localhost:1555/I012T4/NEW-IMAGE?type=REACTION&object=TRANS-RXN-283&orgids=(IN55195%20I012T4%20INKY1%20IN366%20IN370)) |  |  |  |
| [L-ascorbate_[extracellular space]_ + H^+^_[extracellular space]_ → L-ascorbate_[cytosol]_ + H^+^_[cytosol]_](http://localhost:1555/compare-frame-in-orgs?type=NIL&object=TRANS-RXN-208&orgids=(IN55195+I012T4+INKY1+IN366+IN370)) |  | [✓](http://localhost:1555/I012T4/NEW-IMAGE?type=REACTION&object=TRANS-RXN-208&orgids=(IN55195%20I012T4%20INKY1%20IN366%20IN370)) |  |  |  |
| [L-carnitine_[extracellular space]_ + ATP + H_2_O → L-carnitine_[cytosol]_ + ADP + phosphate + H^+^](http://localhost:1555/compare-frame-in-orgs?type=NIL&object=RXN1-7&orgids=(IN55195+I012T4+INKY1+IN366+IN370)) |  | [✓](http://localhost:1555/I012T4/NEW-IMAGE?type=REACTION&object=RXN1-7&orgids=(IN55195%20I012T4%20INKY1%20IN366%20IN370)) |  |  |  |
| [Mg^2+^_[extracellular space]_ → Mg^2+^_[cytosol]_](http://localhost:1555/compare-frame-in-orgs?type=NIL&object=TRANS-RXN-141&orgids=(IN55195+I012T4+INKY1+IN366+IN370)) |  | [✓](http://localhost:1555/I012T4/NEW-IMAGE?type=REACTION&object=TRANS-RXN-141&orgids=(IN55195%20I012T4%20INKY1%20IN366%20IN370)) |  |  |  |
| [Na^+^_[extracellular space]_ + ATP + H_2_O → Na^+^_[cytosol]_ + ADP + phosphate + H^+^](http://localhost:1555/compare-frame-in-orgs?type=NIL&object=RXN1-3&orgids=(IN55195+I012T4+INKY1+IN366+IN370)) |  | [✓](http://localhost:1555/I012T4/NEW-IMAGE?type=REACTION&object=RXN1-3&orgids=(IN55195%20I012T4%20INKY1%20IN366%20IN370)) |  |  |  |
| [phosphate_[cytosol]_ ← phosphate_[extracellular space]_](http://localhost:1555/compare-frame-in-orgs?type=NIL&object=TRANS-RXN0-470&orgids=(IN55195+I012T4+INKY1+IN366+IN370)) |  | [✓](http://localhost:1555/I012T4/NEW-IMAGE?type=REACTION&object=TRANS-RXN0-470&orgids=(IN55195%20I012T4%20INKY1%20IN366%20IN370)) |  |  |  |
| [selenophosphate + a 5-methylaminomethyl-2-(*S*-geranyl)thiouridine^34^ in tRNA → a 5-methylaminomethyl-2-(*Se*-phospho)selenouridine^34^ in tRNA + (2E)-3,7-dimethylocta-2,6-diene-1-thiol](http://localhost:1555/compare-frame-in-orgs?type=NIL&object=RXN-20756&orgids=(IN55195+I012T4+INKY1+IN366+IN370)) |  | [✓](http://localhost:1555/I012T4/NEW-IMAGE?type=REACTION&object=RXN-20756&orgids=(IN55195%20I012T4%20INKY1%20IN366%20IN370)) |  |  |  |
| [Und-PP-{β-D-GlcNAc-(1→4)-Mur2Ac-L-Ala-γ-D-iGln-[*N*^6^-(Gly_5_)]-L-Lys-D-Ala-D-Ala}_(n+1)_ + a peptidoglycan internal segment (*S. aureus*) → a peptidoglycan with D,D cross-link (S. aureus) + D-alanine](http://localhost:1555/compare-frame-in-orgs?type=NIL&object=RXN-11065&orgids=(IN55195+I012T4+INKY1+IN366+IN370)) |  |  |  |  | [✓](http://localhost:1555/IN370/NEW-IMAGE?type=REACTION&object=RXN-11065&orgids=(IN55195%20I012T4%20INKY1%20IN366%20IN370)) |
| [uracil_[cytosol]_ ← uracil_[extracellular space]_](http://localhost:1555/compare-frame-in-orgs?type=NIL&object=TRANS-RXN0-560&orgids=(IN55195+I012T4+INKY1+IN366+IN370)) |  | [✓](http://localhost:1555/I012T4/NEW-IMAGE?type=REACTION&object=TRANS-RXN0-560&orgids=(IN55195%20I012T4%20INKY1%20IN366%20IN370)) |  |  |  |
| [uracil_[extracellular space]_ + H^+^_[extracellular space]_ ↔ uracil_[cytosol]_ + H^+^_[cytosol]_](http://localhost:1555/compare-frame-in-orgs?type=NIL&object=TRANS-RXN-132&orgids=(IN55195+I012T4+INKY1+IN366+IN370)) |  | [✓](http://localhost:1555/I012T4/NEW-IMAGE?type=REACTION&object=TRANS-RXN-132&orgids=(IN55195%20I012T4%20INKY1%20IN366%20IN370)) |  |  |  |
| [xanthine_[extracellular space]_ + uracil_[extracellular space]_ → xanthine_[cytosol]_ + uracil_[cytosol]_](http://localhost:1555/compare-frame-in-orgs?type=NIL&object=TRANS-RXN1-9&orgids=(IN55195+I012T4+INKY1+IN366+IN370)) |  | [✓](http://localhost:1555/I012T4/NEW-IMAGE?type=REACTION&object=TRANS-RXN1-9&orgids=(IN55195%20I012T4%20INKY1%20IN366%20IN370)) |  |  |  |
| [Zn^2+^_[extracellular space]_ + ATP + H_2_O → Zn^2+^_[cytosol]_ + ADP + phosphate + H^+^](http://localhost:1555/compare-frame-in-orgs?type=NIL&object=ABC-63-RXN&orgids=(IN55195+I012T4+INKY1+IN366+IN370)) |  | [✓](http://localhost:1555/I012T4/NEW-IMAGE?type=REACTION&object=ABC-63-RXN&orgids=(IN55195%20I012T4%20INKY1%20IN366%20IN370)) |  |  |  |
| Total | [6](http://localhost:1555/comp-genomics?type=(REACTION-UNIQUE+UNIQUE-REACTIONS)&orgids=IN55195) | [51](http://localhost:1555/comp-genomics?type=(REACTION-UNIQUE+UNIQUE-REACTIONS)&orgids=I012T4) | [3](http://localhost:1555/comp-genomics?type=(REACTION-UNIQUE+UNIQUE-REACTIONS)&orgids=INKY1) | [4](http://localhost:1555/comp-genomics?type=(REACTION-UNIQUE+UNIQUE-REACTIONS)&orgids=IN366) | [1](http://localhost:1555/comp-genomics?type=(REACTION-UNIQUE+UNIQUE-REACTIONS)&orgids=IN370) |

**Pathways**

**Supplemental Table 5G: Pathway Comparison by Pathway Class**

This table presents statistics on the number of pathways present in each pathway class. The two largest top-level classes, Biosynthesis and Degradation/Utilization/Assimilation, are broken down further to show the distribution of pathways among their next-level subclasses. The vast majority of pathways are assigned to only a single class. However, a small number may be assigned to more than one class; such pathways would be double-counted, making the Total line in this table different from the totals in the Shared Pathways table.

| [**Pathway Class**](http://localhost:1555/comp-genomics?type=PWY-CLASS-DIST&orgids=(IN55195+I012T4+INKY1+IN366+IN370)) | [*L.iners* ATCC 55195](http://localhost:1555/IN55195/organism-summary) | [*L.iners* I012T4](http://localhost:1555/I012T4/organism-summary) | [*L.iners* KY](http://localhost:1555/INKY1/organism-summary) | [*L.iners* pt1](http://localhost:1555/IN366/organism-summary) | [*L.iners* pt2](http://localhost:1555/IN370/organism-summary) |
| --- | --- | --- | --- | --- | --- |
| [Biosynthesis](http://localhost:1555/comp-genomics?type=(PWY-CLASS-DIST+%7cBiosynthesis%7c)&orgids=(IN55195+I012T4+INKY1+IN366+IN370)) | 79 | 76 | 74 | 80 | 72 |
| [Amine and Polyamine Biosynthesis](http://localhost:1555/comp-genomics?type=(PWY-CLASS-DIST+%7cPolyamine-Biosynthesis%7c)&orgids=(IN55195+I012T4+INKY1+IN366+IN370)" \t "_blank) | [0](http://localhost:1555/comp-genomics?type=(PWY-CLASS-DIST+%7cPolyamine-Biosynthesis%7c)&orgids=IN55195) | [0](http://localhost:1555/comp-genomics?type=(PWY-CLASS-DIST+%7cPolyamine-Biosynthesis%7c)&orgids=I012T4) | [0](http://localhost:1555/comp-genomics?type=(PWY-CLASS-DIST+%7cPolyamine-Biosynthesis%7c)&orgids=INKY1) | [0](http://localhost:1555/comp-genomics?type=(PWY-CLASS-DIST+%7cPolyamine-Biosynthesis%7c)&orgids=IN366) | [0](http://localhost:1555/comp-genomics?type=(PWY-CLASS-DIST+%7cPolyamine-Biosynthesis%7c)&orgids=IN370) |
| [Amino Acid Biosynthesis](http://localhost:1555/comp-genomics?type=(PWY-CLASS-DIST+%7cAmino-Acid-Biosynthesis%7c)&orgids=(IN55195+I012T4+INKY1+IN366+IN370)" \t "_blank) | [7](http://localhost:1555/comp-genomics?type=(PWY-CLASS-DIST+%7cAmino-Acid-Biosynthesis%7c)&orgids=IN55195) | [6](http://localhost:1555/comp-genomics?type=(PWY-CLASS-DIST+%7cAmino-Acid-Biosynthesis%7c)&orgids=I012T4) | [5](http://localhost:1555/comp-genomics?type=(PWY-CLASS-DIST+%7cAmino-Acid-Biosynthesis%7c)&orgids=INKY1) | [7](http://localhost:1555/comp-genomics?type=(PWY-CLASS-DIST+%7cAmino-Acid-Biosynthesis%7c)&orgids=IN366) | [7](http://localhost:1555/comp-genomics?type=(PWY-CLASS-DIST+%7cAmino-Acid-Biosynthesis%7c)&orgids=IN370) |
| [Aminoacyl-tRNA Charging](http://localhost:1555/comp-genomics?type=(PWY-CLASS-DIST+%7cAminoacyl-tRNAs-Charging%7c)&orgids=(IN55195+I012T4+INKY1+IN366+IN370)" \t "_blank) | [3](http://localhost:1555/comp-genomics?type=(PWY-CLASS-DIST+%7cAminoacyl-tRNAs-Charging%7c)&orgids=IN55195) | [2](http://localhost:1555/comp-genomics?type=(PWY-CLASS-DIST+%7cAminoacyl-tRNAs-Charging%7c)&orgids=I012T4) | [2](http://localhost:1555/comp-genomics?type=(PWY-CLASS-DIST+%7cAminoacyl-tRNAs-Charging%7c)&orgids=INKY1) | [3](http://localhost:1555/comp-genomics?type=(PWY-CLASS-DIST+%7cAminoacyl-tRNAs-Charging%7c)&orgids=IN366) | [3](http://localhost:1555/comp-genomics?type=(PWY-CLASS-DIST+%7cAminoacyl-tRNAs-Charging%7c)&orgids=IN370) |
| [Aromatic Compound Biosynthesis](http://localhost:1555/comp-genomics?type=(PWY-CLASS-DIST+AROMATIC-COMPOUNDS-BIOSYN)&orgids=(IN55195+I012T4+INKY1+IN366+IN370)" \t "_blank) | [3](http://localhost:1555/comp-genomics?type=(PWY-CLASS-DIST+AROMATIC-COMPOUNDS-BIOSYN)&orgids=IN55195) | [3](http://localhost:1555/comp-genomics?type=(PWY-CLASS-DIST+AROMATIC-COMPOUNDS-BIOSYN)&orgids=I012T4) | [3](http://localhost:1555/comp-genomics?type=(PWY-CLASS-DIST+AROMATIC-COMPOUNDS-BIOSYN)&orgids=INKY1) | [3](http://localhost:1555/comp-genomics?type=(PWY-CLASS-DIST+AROMATIC-COMPOUNDS-BIOSYN)&orgids=IN366) | [3](http://localhost:1555/comp-genomics?type=(PWY-CLASS-DIST+AROMATIC-COMPOUNDS-BIOSYN)&orgids=IN370) |
| [Carbohydrate Biosynthesis](http://localhost:1555/comp-genomics?type=(PWY-CLASS-DIST+%7cCarbohydrates-Biosynthesis%7c)&orgids=(IN55195+I012T4+INKY1+IN366+IN370)" \t "_blank) | [4](http://localhost:1555/comp-genomics?type=(PWY-CLASS-DIST+%7cCarbohydrates-Biosynthesis%7c)&orgids=IN55195) | [4](http://localhost:1555/comp-genomics?type=(PWY-CLASS-DIST+%7cCarbohydrates-Biosynthesis%7c)&orgids=I012T4) | [4](http://localhost:1555/comp-genomics?type=(PWY-CLASS-DIST+%7cCarbohydrates-Biosynthesis%7c)&orgids=INKY1) | [4](http://localhost:1555/comp-genomics?type=(PWY-CLASS-DIST+%7cCarbohydrates-Biosynthesis%7c)&orgids=IN366) | [4](http://localhost:1555/comp-genomics?type=(PWY-CLASS-DIST+%7cCarbohydrates-Biosynthesis%7c)&orgids=IN370) |
| [Cell Structure Biosynthesis](http://localhost:1555/comp-genomics?type=(PWY-CLASS-DIST+%7cCell-Structure-Biosynthesis%7c)&orgids=(IN55195+I012T4+INKY1+IN366+IN370)" \t "_blank) | [3](http://localhost:1555/comp-genomics?type=(PWY-CLASS-DIST+%7cCell-Structure-Biosynthesis%7c)&orgids=IN55195) | [3](http://localhost:1555/comp-genomics?type=(PWY-CLASS-DIST+%7cCell-Structure-Biosynthesis%7c)&orgids=I012T4) | [3](http://localhost:1555/comp-genomics?type=(PWY-CLASS-DIST+%7cCell-Structure-Biosynthesis%7c)&orgids=INKY1) | [3](http://localhost:1555/comp-genomics?type=(PWY-CLASS-DIST+%7cCell-Structure-Biosynthesis%7c)&orgids=IN366) | [1](http://localhost:1555/comp-genomics?type=(PWY-CLASS-DIST+%7cCell-Structure-Biosynthesis%7c)&orgids=IN370) |
| [Cofactor, Carrier, and Vitamin Biosynthesis](http://localhost:1555/comp-genomics?type=(PWY-CLASS-DIST+%7cCofactor-Biosynthesis%7c)&orgids=(IN55195+I012T4+INKY1+IN366+IN370)" \t "_blank) | [24](http://localhost:1555/comp-genomics?type=(PWY-CLASS-DIST+%7cCofactor-Biosynthesis%7c)&orgids=IN55195) | [24](http://localhost:1555/comp-genomics?type=(PWY-CLASS-DIST+%7cCofactor-Biosynthesis%7c)&orgids=I012T4) | [24](http://localhost:1555/comp-genomics?type=(PWY-CLASS-DIST+%7cCofactor-Biosynthesis%7c)&orgids=INKY1) | [24](http://localhost:1555/comp-genomics?type=(PWY-CLASS-DIST+%7cCofactor-Biosynthesis%7c)&orgids=IN366) | [20](http://localhost:1555/comp-genomics?type=(PWY-CLASS-DIST+%7cCofactor-Biosynthesis%7c)&orgids=IN370) |
| [Fatty Acid and Lipid Biosynthesis](http://localhost:1555/comp-genomics?type=(PWY-CLASS-DIST+%7cLipid-Biosynthesis%7c)&orgids=(IN55195+I012T4+INKY1+IN366+IN370)" \t "_blank) | [6](http://localhost:1555/comp-genomics?type=(PWY-CLASS-DIST+%7cLipid-Biosynthesis%7c)&orgids=IN55195) | [4](http://localhost:1555/comp-genomics?type=(PWY-CLASS-DIST+%7cLipid-Biosynthesis%7c)&orgids=I012T4) | [4](http://localhost:1555/comp-genomics?type=(PWY-CLASS-DIST+%7cLipid-Biosynthesis%7c)&orgids=INKY1) | [7](http://localhost:1555/comp-genomics?type=(PWY-CLASS-DIST+%7cLipid-Biosynthesis%7c)&orgids=IN366) | [6](http://localhost:1555/comp-genomics?type=(PWY-CLASS-DIST+%7cLipid-Biosynthesis%7c)&orgids=IN370) |
| [Metabolic Regulator Biosynthesis](http://localhost:1555/comp-genomics?type=(PWY-CLASS-DIST+%7cMetabolic-Regulators%7c)&orgids=(IN55195+I012T4+INKY1+IN366+IN370)" \t "_blank) | [1](http://localhost:1555/comp-genomics?type=(PWY-CLASS-DIST+%7cMetabolic-Regulators%7c)&orgids=IN55195) | [1](http://localhost:1555/comp-genomics?type=(PWY-CLASS-DIST+%7cMetabolic-Regulators%7c)&orgids=I012T4) | [1](http://localhost:1555/comp-genomics?type=(PWY-CLASS-DIST+%7cMetabolic-Regulators%7c)&orgids=INKY1) | [1](http://localhost:1555/comp-genomics?type=(PWY-CLASS-DIST+%7cMetabolic-Regulators%7c)&orgids=IN366) | [1](http://localhost:1555/comp-genomics?type=(PWY-CLASS-DIST+%7cMetabolic-Regulators%7c)&orgids=IN370) |
| [Nucleoside and Nucleotide Biosynthesis](http://localhost:1555/comp-genomics?type=(PWY-CLASS-DIST+%7cNucleotide-Biosynthesis%7c)&orgids=(IN55195+I012T4+INKY1+IN366+IN370)" \t "_blank) | [19](http://localhost:1555/comp-genomics?type=(PWY-CLASS-DIST+%7cNucleotide-Biosynthesis%7c)&orgids=IN55195) | [19](http://localhost:1555/comp-genomics?type=(PWY-CLASS-DIST+%7cNucleotide-Biosynthesis%7c)&orgids=I012T4) | [19](http://localhost:1555/comp-genomics?type=(PWY-CLASS-DIST+%7cNucleotide-Biosynthesis%7c)&orgids=INKY1) | [19](http://localhost:1555/comp-genomics?type=(PWY-CLASS-DIST+%7cNucleotide-Biosynthesis%7c)&orgids=IN366) | [19](http://localhost:1555/comp-genomics?type=(PWY-CLASS-DIST+%7cNucleotide-Biosynthesis%7c)&orgids=IN370) |
| [Other Biosynthesis](http://localhost:1555/comp-genomics?type=(PWY-CLASS-DIST+%7cOther-biosynthesis%7c)&orgids=(IN55195+I012T4+INKY1+IN366+IN370)" \t "_blank) | [0](http://localhost:1555/comp-genomics?type=(PWY-CLASS-DIST+%7cOther-biosynthesis%7c)&orgids=IN55195) | [0](http://localhost:1555/comp-genomics?type=(PWY-CLASS-DIST+%7cOther-biosynthesis%7c)&orgids=I012T4) | [0](http://localhost:1555/comp-genomics?type=(PWY-CLASS-DIST+%7cOther-biosynthesis%7c)&orgids=INKY1) | [0](http://localhost:1555/comp-genomics?type=(PWY-CLASS-DIST+%7cOther-biosynthesis%7c)&orgids=IN366) | [0](http://localhost:1555/comp-genomics?type=(PWY-CLASS-DIST+%7cOther-biosynthesis%7c)&orgids=IN370) |
| [Polyprenyl Biosynthesis](http://localhost:1555/comp-genomics?type=(PWY-CLASS-DIST+%7cPolyprenyl-Biosynthesis%7c)&orgids=(IN55195+I012T4+INKY1+IN366+IN370)" \t "_blank) | [2](http://localhost:1555/comp-genomics?type=(PWY-CLASS-DIST+%7cPolyprenyl-Biosynthesis%7c)&orgids=IN55195) | [2](http://localhost:1555/comp-genomics?type=(PWY-CLASS-DIST+%7cPolyprenyl-Biosynthesis%7c)&orgids=I012T4) | [2](http://localhost:1555/comp-genomics?type=(PWY-CLASS-DIST+%7cPolyprenyl-Biosynthesis%7c)&orgids=INKY1) | [2](http://localhost:1555/comp-genomics?type=(PWY-CLASS-DIST+%7cPolyprenyl-Biosynthesis%7c)&orgids=IN366) | [2](http://localhost:1555/comp-genomics?type=(PWY-CLASS-DIST+%7cPolyprenyl-Biosynthesis%7c)&orgids=IN370) |
| [Secondary Metabolite Biosynthesis](http://localhost:1555/comp-genomics?type=(PWY-CLASS-DIST+SECONDARY-METABOLITE-BIOSYNTHESIS)&orgids=(IN55195+I012T4+INKY1+IN366+IN370)" \t "_blank) | [1](http://localhost:1555/comp-genomics?type=(PWY-CLASS-DIST+SECONDARY-METABOLITE-BIOSYNTHESIS)&orgids=IN55195) | [1](http://localhost:1555/comp-genomics?type=(PWY-CLASS-DIST+SECONDARY-METABOLITE-BIOSYNTHESIS)&orgids=I012T4) | [1](http://localhost:1555/comp-genomics?type=(PWY-CLASS-DIST+SECONDARY-METABOLITE-BIOSYNTHESIS)&orgids=INKY1) | [1](http://localhost:1555/comp-genomics?type=(PWY-CLASS-DIST+SECONDARY-METABOLITE-BIOSYNTHESIS)&orgids=IN366) | [1](http://localhost:1555/comp-genomics?type=(PWY-CLASS-DIST+SECONDARY-METABOLITE-BIOSYNTHESIS)&orgids=IN370) |
| [Storage Compound Biosynthesis](http://localhost:1555/comp-genomics?type=(PWY-CLASS-DIST+%7cStorage-Compounds-Biosynthesis%7c)&orgids=(IN55195+I012T4+INKY1+IN366+IN370)" \t "_blank) | [0](http://localhost:1555/comp-genomics?type=(PWY-CLASS-DIST+%7cStorage-Compounds-Biosynthesis%7c)&orgids=IN55195) | [0](http://localhost:1555/comp-genomics?type=(PWY-CLASS-DIST+%7cStorage-Compounds-Biosynthesis%7c)&orgids=I012T4) | [0](http://localhost:1555/comp-genomics?type=(PWY-CLASS-DIST+%7cStorage-Compounds-Biosynthesis%7c)&orgids=INKY1) | [0](http://localhost:1555/comp-genomics?type=(PWY-CLASS-DIST+%7cStorage-Compounds-Biosynthesis%7c)&orgids=IN366) | [0](http://localhost:1555/comp-genomics?type=(PWY-CLASS-DIST+%7cStorage-Compounds-Biosynthesis%7c)&orgids=IN370) |
| [Tetrapyrrole Biosynthesis](http://localhost:1555/comp-genomics?type=(PWY-CLASS-DIST+%7cTetrapyrrole-Biosynthesis%7c)&orgids=(IN55195+I012T4+INKY1+IN366+IN370)" \t "_blank) | [1](http://localhost:1555/comp-genomics?type=(PWY-CLASS-DIST+%7cTetrapyrrole-Biosynthesis%7c)&orgids=IN55195) | [1](http://localhost:1555/comp-genomics?type=(PWY-CLASS-DIST+%7cTetrapyrrole-Biosynthesis%7c)&orgids=I012T4) | [1](http://localhost:1555/comp-genomics?type=(PWY-CLASS-DIST+%7cTetrapyrrole-Biosynthesis%7c)&orgids=INKY1) | [1](http://localhost:1555/comp-genomics?type=(PWY-CLASS-DIST+%7cTetrapyrrole-Biosynthesis%7c)&orgids=IN366) | [1](http://localhost:1555/comp-genomics?type=(PWY-CLASS-DIST+%7cTetrapyrrole-Biosynthesis%7c)&orgids=IN370) |
| [Generation of Precursor Metabolites and Energy](http://localhost:1555/comp-genomics?type=(PWY-CLASS-DIST+%7cEnergy-Metabolism%7c)&orgids=(IN55195+I012T4+INKY1+IN366+IN370)) | [6](http://localhost:1555/comp-genomics?type=(PWY-CLASS-DIST+%7cEnergy-Metabolism%7c)&orgids=IN55195) | [6](http://localhost:1555/comp-genomics?type=(PWY-CLASS-DIST+%7cEnergy-Metabolism%7c)&orgids=I012T4) | [6](http://localhost:1555/comp-genomics?type=(PWY-CLASS-DIST+%7cEnergy-Metabolism%7c)&orgids=INKY1) | [7](http://localhost:1555/comp-genomics?type=(PWY-CLASS-DIST+%7cEnergy-Metabolism%7c)&orgids=IN366) | [6](http://localhost:1555/comp-genomics?type=(PWY-CLASS-DIST+%7cEnergy-Metabolism%7c)&orgids=IN370) |
| [Metabolic Clusters](http://localhost:1555/comp-genomics?type=(PWY-CLASS-DIST+%7cMetabolic-Clusters%7c)&orgids=(IN55195+I012T4+INKY1+IN366+IN370)) | [6](http://localhost:1555/comp-genomics?type=(PWY-CLASS-DIST+%7cMetabolic-Clusters%7c)&orgids=IN55195) | [6](http://localhost:1555/comp-genomics?type=(PWY-CLASS-DIST+%7cMetabolic-Clusters%7c)&orgids=I012T4) | [6](http://localhost:1555/comp-genomics?type=(PWY-CLASS-DIST+%7cMetabolic-Clusters%7c)&orgids=INKY1) | [6](http://localhost:1555/comp-genomics?type=(PWY-CLASS-DIST+%7cMetabolic-Clusters%7c)&orgids=IN366) | [6](http://localhost:1555/comp-genomics?type=(PWY-CLASS-DIST+%7cMetabolic-Clusters%7c)&orgids=IN370) |
| [Bioluminescence](http://localhost:1555/comp-genomics?type=(PWY-CLASS-DIST+%7cBioluminescence%7c)&orgids=(IN55195+I012T4+INKY1+IN366+IN370)) | [0](http://localhost:1555/comp-genomics?type=(PWY-CLASS-DIST+%7cBioluminescence%7c)&orgids=IN55195) | [0](http://localhost:1555/comp-genomics?type=(PWY-CLASS-DIST+%7cBioluminescence%7c)&orgids=I012T4) | [0](http://localhost:1555/comp-genomics?type=(PWY-CLASS-DIST+%7cBioluminescence%7c)&orgids=INKY1) | [0](http://localhost:1555/comp-genomics?type=(PWY-CLASS-DIST+%7cBioluminescence%7c)&orgids=IN366) | [0](http://localhost:1555/comp-genomics?type=(PWY-CLASS-DIST+%7cBioluminescence%7c)&orgids=IN370) |
| [Detoxification](http://localhost:1555/comp-genomics?type=(PWY-CLASS-DIST+%7cDetoxification%7c)&orgids=(IN55195+I012T4+INKY1+IN366+IN370)) | [0](http://localhost:1555/comp-genomics?type=(PWY-CLASS-DIST+%7cDetoxification%7c)&orgids=IN55195) | [2](http://localhost:1555/comp-genomics?type=(PWY-CLASS-DIST+%7cDetoxification%7c)&orgids=I012T4) | [2](http://localhost:1555/comp-genomics?type=(PWY-CLASS-DIST+%7cDetoxification%7c)&orgids=INKY1) | [0](http://localhost:1555/comp-genomics?type=(PWY-CLASS-DIST+%7cDetoxification%7c)&orgids=IN366) | [0](http://localhost:1555/comp-genomics?type=(PWY-CLASS-DIST+%7cDetoxification%7c)&orgids=IN370) |
| [Transport](http://localhost:1555/comp-genomics?type=(PWY-CLASS-DIST+%7cTransport-Pathways%7c)&orgids=(IN55195+I012T4+INKY1+IN366+IN370)) | [0](http://localhost:1555/comp-genomics?type=(PWY-CLASS-DIST+%7cTransport-Pathways%7c)&orgids=IN55195) | [0](http://localhost:1555/comp-genomics?type=(PWY-CLASS-DIST+%7cTransport-Pathways%7c)&orgids=I012T4) | [0](http://localhost:1555/comp-genomics?type=(PWY-CLASS-DIST+%7cTransport-Pathways%7c)&orgids=INKY1) | [0](http://localhost:1555/comp-genomics?type=(PWY-CLASS-DIST+%7cTransport-Pathways%7c)&orgids=IN366) | [0](http://localhost:1555/comp-genomics?type=(PWY-CLASS-DIST+%7cTransport-Pathways%7c)&orgids=IN370) |
| [Macromolecule Modification](http://localhost:1555/comp-genomics?type=(PWY-CLASS-DIST+%7cMacromolecule-Modification%7c)&orgids=(IN55195+I012T4+INKY1+IN366+IN370)) | [4](http://localhost:1555/comp-genomics?type=(PWY-CLASS-DIST+%7cMacromolecule-Modification%7c)&orgids=IN55195) | [5](http://localhost:1555/comp-genomics?type=(PWY-CLASS-DIST+%7cMacromolecule-Modification%7c)&orgids=I012T4) | [5](http://localhost:1555/comp-genomics?type=(PWY-CLASS-DIST+%7cMacromolecule-Modification%7c)&orgids=INKY1) | [4](http://localhost:1555/comp-genomics?type=(PWY-CLASS-DIST+%7cMacromolecule-Modification%7c)&orgids=IN366) | [5](http://localhost:1555/comp-genomics?type=(PWY-CLASS-DIST+%7cMacromolecule-Modification%7c)&orgids=IN370) |
| [Activation/Inactivation/Interconversion](http://localhost:1555/comp-genomics?type=(PWY-CLASS-DIST+%7cActivation-Inactivation-Interconversion%7c)&orgids=(IN55195+I012T4+INKY1+IN366+IN370)) | [1](http://localhost:1555/comp-genomics?type=(PWY-CLASS-DIST+%7cActivation-Inactivation-Interconversion%7c)&orgids=IN55195) | [1](http://localhost:1555/comp-genomics?type=(PWY-CLASS-DIST+%7cActivation-Inactivation-Interconversion%7c)&orgids=I012T4) | [1](http://localhost:1555/comp-genomics?type=(PWY-CLASS-DIST+%7cActivation-Inactivation-Interconversion%7c)&orgids=INKY1) | [1](http://localhost:1555/comp-genomics?type=(PWY-CLASS-DIST+%7cActivation-Inactivation-Interconversion%7c)&orgids=IN366) | [1](http://localhost:1555/comp-genomics?type=(PWY-CLASS-DIST+%7cActivation-Inactivation-Interconversion%7c)&orgids=IN370) |
| [Degradation/Utilization/Assimilation](http://localhost:1555/comp-genomics?type=(PWY-CLASS-DIST+%7cDegradation%7c)&orgids=(IN55195+I012T4+INKY1+IN366+IN370)) | 28 | 24 | 24 | 30 | 25 |
| [Alcohol Degradation](http://localhost:1555/comp-genomics?type=(PWY-CLASS-DIST+%7cAlcohol-Degradation%7c)&orgids=(IN55195+I012T4+INKY1+IN366+IN370)" \t "_blank) | [4](http://localhost:1555/comp-genomics?type=(PWY-CLASS-DIST+%7cAlcohol-Degradation%7c)&orgids=IN55195) | [4](http://localhost:1555/comp-genomics?type=(PWY-CLASS-DIST+%7cAlcohol-Degradation%7c)&orgids=I012T4) | [4](http://localhost:1555/comp-genomics?type=(PWY-CLASS-DIST+%7cAlcohol-Degradation%7c)&orgids=INKY1) | [5](http://localhost:1555/comp-genomics?type=(PWY-CLASS-DIST+%7cAlcohol-Degradation%7c)&orgids=IN366) | [3](http://localhost:1555/comp-genomics?type=(PWY-CLASS-DIST+%7cAlcohol-Degradation%7c)&orgids=IN370) |
| [Aldehyde Degradation](http://localhost:1555/comp-genomics?type=(PWY-CLASS-DIST+%7cAldehyde-Degradation%7c)&orgids=(IN55195+I012T4+INKY1+IN366+IN370)" \t "_blank) | [0](http://localhost:1555/comp-genomics?type=(PWY-CLASS-DIST+%7cAldehyde-Degradation%7c)&orgids=IN55195) | [0](http://localhost:1555/comp-genomics?type=(PWY-CLASS-DIST+%7cAldehyde-Degradation%7c)&orgids=I012T4) | [0](http://localhost:1555/comp-genomics?type=(PWY-CLASS-DIST+%7cAldehyde-Degradation%7c)&orgids=INKY1) | [0](http://localhost:1555/comp-genomics?type=(PWY-CLASS-DIST+%7cAldehyde-Degradation%7c)&orgids=IN366) | [0](http://localhost:1555/comp-genomics?type=(PWY-CLASS-DIST+%7cAldehyde-Degradation%7c)&orgids=IN370) |
| [Amine and Polyamine Degradation](http://localhost:1555/comp-genomics?type=(PWY-CLASS-DIST+AMINE-DEG)&orgids=(IN55195+I012T4+INKY1+IN366+IN370)" \t "_blank) | [2](http://localhost:1555/comp-genomics?type=(PWY-CLASS-DIST+AMINE-DEG)&orgids=IN55195) | [2](http://localhost:1555/comp-genomics?type=(PWY-CLASS-DIST+AMINE-DEG)&orgids=I012T4) | [1](http://localhost:1555/comp-genomics?type=(PWY-CLASS-DIST+AMINE-DEG)&orgids=INKY1) | [2](http://localhost:1555/comp-genomics?type=(PWY-CLASS-DIST+AMINE-DEG)&orgids=IN366) | [2](http://localhost:1555/comp-genomics?type=(PWY-CLASS-DIST+AMINE-DEG)&orgids=IN370) |
| [Amino Acid Degradation](http://localhost:1555/comp-genomics?type=(PWY-CLASS-DIST+%7cAmino-Acid-Degradation%7c)&orgids=(IN55195+I012T4+INKY1+IN366+IN370)" \t "_blank) | [6](http://localhost:1555/comp-genomics?type=(PWY-CLASS-DIST+%7cAmino-Acid-Degradation%7c)&orgids=IN55195) | [4](http://localhost:1555/comp-genomics?type=(PWY-CLASS-DIST+%7cAmino-Acid-Degradation%7c)&orgids=I012T4) | [5](http://localhost:1555/comp-genomics?type=(PWY-CLASS-DIST+%7cAmino-Acid-Degradation%7c)&orgids=INKY1) | [6](http://localhost:1555/comp-genomics?type=(PWY-CLASS-DIST+%7cAmino-Acid-Degradation%7c)&orgids=IN366) | [5](http://localhost:1555/comp-genomics?type=(PWY-CLASS-DIST+%7cAmino-Acid-Degradation%7c)&orgids=IN370) |
| [Aromatic Compound Degradation](http://localhost:1555/comp-genomics?type=(PWY-CLASS-DIST+AROMATIC-COMPOUNDS-DEGRADATION)&orgids=(IN55195+I012T4+INKY1+IN366+IN370)" \t "_blank) | [0](http://localhost:1555/comp-genomics?type=(PWY-CLASS-DIST+AROMATIC-COMPOUNDS-DEGRADATION)&orgids=IN55195) | [0](http://localhost:1555/comp-genomics?type=(PWY-CLASS-DIST+AROMATIC-COMPOUNDS-DEGRADATION)&orgids=I012T4) | [0](http://localhost:1555/comp-genomics?type=(PWY-CLASS-DIST+AROMATIC-COMPOUNDS-DEGRADATION)&orgids=INKY1) | [0](http://localhost:1555/comp-genomics?type=(PWY-CLASS-DIST+AROMATIC-COMPOUNDS-DEGRADATION)&orgids=IN366) | [0](http://localhost:1555/comp-genomics?type=(PWY-CLASS-DIST+AROMATIC-COMPOUNDS-DEGRADATION)&orgids=IN370) |
| [C1 Compound Utilization and Assimilation](http://localhost:1555/comp-genomics?type=(PWY-CLASS-DIST+C1-COMPOUNDS)&orgids=(IN55195+I012T4+INKY1+IN366+IN370)" \t "_blank) | [1](http://localhost:1555/comp-genomics?type=(PWY-CLASS-DIST+C1-COMPOUNDS)&orgids=IN55195) | [1](http://localhost:1555/comp-genomics?type=(PWY-CLASS-DIST+C1-COMPOUNDS)&orgids=I012T4) | [1](http://localhost:1555/comp-genomics?type=(PWY-CLASS-DIST+C1-COMPOUNDS)&orgids=INKY1) | [1](http://localhost:1555/comp-genomics?type=(PWY-CLASS-DIST+C1-COMPOUNDS)&orgids=IN366) | [1](http://localhost:1555/comp-genomics?type=(PWY-CLASS-DIST+C1-COMPOUNDS)&orgids=IN370) |
| [Carbohydrate Degradation](http://localhost:1555/comp-genomics?type=(PWY-CLASS-DIST+%7cCarbohydrates-Degradation%7c)&orgids=(IN55195+I012T4+INKY1+IN366+IN370)" \t "_blank) | [5](http://localhost:1555/comp-genomics?type=(PWY-CLASS-DIST+%7cCarbohydrates-Degradation%7c)&orgids=IN55195) | [4](http://localhost:1555/comp-genomics?type=(PWY-CLASS-DIST+%7cCarbohydrates-Degradation%7c)&orgids=I012T4) | [4](http://localhost:1555/comp-genomics?type=(PWY-CLASS-DIST+%7cCarbohydrates-Degradation%7c)&orgids=INKY1) | [5](http://localhost:1555/comp-genomics?type=(PWY-CLASS-DIST+%7cCarbohydrates-Degradation%7c)&orgids=IN366) | [4](http://localhost:1555/comp-genomics?type=(PWY-CLASS-DIST+%7cCarbohydrates-Degradation%7c)&orgids=IN370) |
| [Carboxylate Degradation](http://localhost:1555/comp-genomics?type=(PWY-CLASS-DIST+CARBOXYLATES-DEG)&orgids=(IN55195+I012T4+INKY1+IN366+IN370)" \t "_blank) | [2](http://localhost:1555/comp-genomics?type=(PWY-CLASS-DIST+CARBOXYLATES-DEG)&orgids=IN55195) | [2](http://localhost:1555/comp-genomics?type=(PWY-CLASS-DIST+CARBOXYLATES-DEG)&orgids=I012T4) | [2](http://localhost:1555/comp-genomics?type=(PWY-CLASS-DIST+CARBOXYLATES-DEG)&orgids=INKY1) | [2](http://localhost:1555/comp-genomics?type=(PWY-CLASS-DIST+CARBOXYLATES-DEG)&orgids=IN366) | [2](http://localhost:1555/comp-genomics?type=(PWY-CLASS-DIST+CARBOXYLATES-DEG)&orgids=IN370) |
| [Chlorinated Compound Degradation](http://localhost:1555/comp-genomics?type=(PWY-CLASS-DIST+CHLORINATED-COMPOUNDS-DEG)&orgids=(IN55195+I012T4+INKY1+IN366+IN370)" \t "_blank) | [0](http://localhost:1555/comp-genomics?type=(PWY-CLASS-DIST+CHLORINATED-COMPOUNDS-DEG)&orgids=IN55195) | [0](http://localhost:1555/comp-genomics?type=(PWY-CLASS-DIST+CHLORINATED-COMPOUNDS-DEG)&orgids=I012T4) | [0](http://localhost:1555/comp-genomics?type=(PWY-CLASS-DIST+CHLORINATED-COMPOUNDS-DEG)&orgids=INKY1) | [0](http://localhost:1555/comp-genomics?type=(PWY-CLASS-DIST+CHLORINATED-COMPOUNDS-DEG)&orgids=IN366) | [0](http://localhost:1555/comp-genomics?type=(PWY-CLASS-DIST+CHLORINATED-COMPOUNDS-DEG)&orgids=IN370) |
| [Cofactor, Prosthetic Group, Electron Carrier Degradation](http://localhost:1555/comp-genomics?type=(PWY-CLASS-DIST+COFACTOR-DEGRADATION)&orgids=(IN55195+I012T4+INKY1+IN366+IN370)" \t "_blank) | [0](http://localhost:1555/comp-genomics?type=(PWY-CLASS-DIST+COFACTOR-DEGRADATION)&orgids=IN55195) | [0](http://localhost:1555/comp-genomics?type=(PWY-CLASS-DIST+COFACTOR-DEGRADATION)&orgids=I012T4) | [0](http://localhost:1555/comp-genomics?type=(PWY-CLASS-DIST+COFACTOR-DEGRADATION)&orgids=INKY1) | [0](http://localhost:1555/comp-genomics?type=(PWY-CLASS-DIST+COFACTOR-DEGRADATION)&orgids=IN366) | [0](http://localhost:1555/comp-genomics?type=(PWY-CLASS-DIST+COFACTOR-DEGRADATION)&orgids=IN370) |
| [Degradation/Utilization/Assimilation - Other](http://localhost:1555/comp-genomics?type=(PWY-CLASS-DIST+%7cOther-Degradation%7c)&orgids=(IN55195+I012T4+INKY1+IN366+IN370)" \t "_blank) | [0](http://localhost:1555/comp-genomics?type=(PWY-CLASS-DIST+%7cOther-Degradation%7c)&orgids=IN55195) | [0](http://localhost:1555/comp-genomics?type=(PWY-CLASS-DIST+%7cOther-Degradation%7c)&orgids=I012T4) | [0](http://localhost:1555/comp-genomics?type=(PWY-CLASS-DIST+%7cOther-Degradation%7c)&orgids=INKY1) | [0](http://localhost:1555/comp-genomics?type=(PWY-CLASS-DIST+%7cOther-Degradation%7c)&orgids=IN366) | [0](http://localhost:1555/comp-genomics?type=(PWY-CLASS-DIST+%7cOther-Degradation%7c)&orgids=IN370) |
| [Fatty Acid and Lipid Degradation](http://localhost:1555/comp-genomics?type=(PWY-CLASS-DIST+%7cFatty-Acid-and-Lipid-Degradation%7c)&orgids=(IN55195+I012T4+INKY1+IN366+IN370)" \t "_blank) | [0](http://localhost:1555/comp-genomics?type=(PWY-CLASS-DIST+%7cFatty-Acid-and-Lipid-Degradation%7c)&orgids=IN55195) | [0](http://localhost:1555/comp-genomics?type=(PWY-CLASS-DIST+%7cFatty-Acid-and-Lipid-Degradation%7c)&orgids=I012T4) | [0](http://localhost:1555/comp-genomics?type=(PWY-CLASS-DIST+%7cFatty-Acid-and-Lipid-Degradation%7c)&orgids=INKY1) | [1](http://localhost:1555/comp-genomics?type=(PWY-CLASS-DIST+%7cFatty-Acid-and-Lipid-Degradation%7c)&orgids=IN366) | [0](http://localhost:1555/comp-genomics?type=(PWY-CLASS-DIST+%7cFatty-Acid-and-Lipid-Degradation%7c)&orgids=IN370) |
| [Hormone Degradation](http://localhost:1555/comp-genomics?type=(PWY-CLASS-DIST+HORMONE-DEG)&orgids=(IN55195+I012T4+INKY1+IN366+IN370)" \t "_blank) | [0](http://localhost:1555/comp-genomics?type=(PWY-CLASS-DIST+HORMONE-DEG)&orgids=IN55195) | [0](http://localhost:1555/comp-genomics?type=(PWY-CLASS-DIST+HORMONE-DEG)&orgids=I012T4) | [0](http://localhost:1555/comp-genomics?type=(PWY-CLASS-DIST+HORMONE-DEG)&orgids=INKY1) | [0](http://localhost:1555/comp-genomics?type=(PWY-CLASS-DIST+HORMONE-DEG)&orgids=IN366) | [0](http://localhost:1555/comp-genomics?type=(PWY-CLASS-DIST+HORMONE-DEG)&orgids=IN370) |
| [Inorganic Nutrient Metabolism](http://localhost:1555/comp-genomics?type=(PWY-CLASS-DIST+%7cNoncarbon-Nutrients%7c)&orgids=(IN55195+I012T4+INKY1+IN366+IN370)" \t "_blank) | [2](http://localhost:1555/comp-genomics?type=(PWY-CLASS-DIST+%7cNoncarbon-Nutrients%7c)&orgids=IN55195) | [2](http://localhost:1555/comp-genomics?type=(PWY-CLASS-DIST+%7cNoncarbon-Nutrients%7c)&orgids=I012T4) | [2](http://localhost:1555/comp-genomics?type=(PWY-CLASS-DIST+%7cNoncarbon-Nutrients%7c)&orgids=INKY1) | [2](http://localhost:1555/comp-genomics?type=(PWY-CLASS-DIST+%7cNoncarbon-Nutrients%7c)&orgids=IN366) | [2](http://localhost:1555/comp-genomics?type=(PWY-CLASS-DIST+%7cNoncarbon-Nutrients%7c)&orgids=IN370) |
| [Nucleoside and Nucleotide Degradation](http://localhost:1555/comp-genomics?type=(PWY-CLASS-DIST+NUCLEO-DEG)&orgids=(IN55195+I012T4+INKY1+IN366+IN370)" \t "_blank) | [3](http://localhost:1555/comp-genomics?type=(PWY-CLASS-DIST+NUCLEO-DEG)&orgids=IN55195) | [3](http://localhost:1555/comp-genomics?type=(PWY-CLASS-DIST+NUCLEO-DEG)&orgids=I012T4) | [3](http://localhost:1555/comp-genomics?type=(PWY-CLASS-DIST+NUCLEO-DEG)&orgids=INKY1) | [3](http://localhost:1555/comp-genomics?type=(PWY-CLASS-DIST+NUCLEO-DEG)&orgids=IN366) | [3](http://localhost:1555/comp-genomics?type=(PWY-CLASS-DIST+NUCLEO-DEG)&orgids=IN370) |
| [Polymeric Compound Degradation](http://localhost:1555/comp-genomics?type=(PWY-CLASS-DIST+%7cPolymer-Degradation%7c)&orgids=(IN55195+I012T4+INKY1+IN366+IN370)" \t "_blank) | [0](http://localhost:1555/comp-genomics?type=(PWY-CLASS-DIST+%7cPolymer-Degradation%7c)&orgids=IN55195) | [0](http://localhost:1555/comp-genomics?type=(PWY-CLASS-DIST+%7cPolymer-Degradation%7c)&orgids=I012T4) | [0](http://localhost:1555/comp-genomics?type=(PWY-CLASS-DIST+%7cPolymer-Degradation%7c)&orgids=INKY1) | [0](http://localhost:1555/comp-genomics?type=(PWY-CLASS-DIST+%7cPolymer-Degradation%7c)&orgids=IN366) | [0](http://localhost:1555/comp-genomics?type=(PWY-CLASS-DIST+%7cPolymer-Degradation%7c)&orgids=IN370) |
| [Protein Degradation](http://localhost:1555/comp-genomics?type=(PWY-CLASS-DIST+%7cProtein-Degradation%7c)&orgids=(IN55195+I012T4+INKY1+IN366+IN370)" \t "_blank) | [0](http://localhost:1555/comp-genomics?type=(PWY-CLASS-DIST+%7cProtein-Degradation%7c)&orgids=IN55195) | [0](http://localhost:1555/comp-genomics?type=(PWY-CLASS-DIST+%7cProtein-Degradation%7c)&orgids=I012T4) | [0](http://localhost:1555/comp-genomics?type=(PWY-CLASS-DIST+%7cProtein-Degradation%7c)&orgids=INKY1) | [0](http://localhost:1555/comp-genomics?type=(PWY-CLASS-DIST+%7cProtein-Degradation%7c)&orgids=IN366) | [0](http://localhost:1555/comp-genomics?type=(PWY-CLASS-DIST+%7cProtein-Degradation%7c)&orgids=IN370) |
| [Secondary Metabolite Degradation](http://localhost:1555/comp-genomics?type=(PWY-CLASS-DIST+SECONDARY-METABOLITE-DEGRADATION)&orgids=(IN55195+I012T4+INKY1+IN366+IN370)" \t "_blank) | [2](http://localhost:1555/comp-genomics?type=(PWY-CLASS-DIST+SECONDARY-METABOLITE-DEGRADATION)&orgids=IN55195) | [1](http://localhost:1555/comp-genomics?type=(PWY-CLASS-DIST+SECONDARY-METABOLITE-DEGRADATION)&orgids=I012T4) | [1](http://localhost:1555/comp-genomics?type=(PWY-CLASS-DIST+SECONDARY-METABOLITE-DEGRADATION)&orgids=INKY1) | [2](http://localhost:1555/comp-genomics?type=(PWY-CLASS-DIST+SECONDARY-METABOLITE-DEGRADATION)&orgids=IN366) | [2](http://localhost:1555/comp-genomics?type=(PWY-CLASS-DIST+SECONDARY-METABOLITE-DEGRADATION)&orgids=IN370) |
| [Glycan Pathways](http://localhost:1555/comp-genomics?type=(PWY-CLASS-DIST+%7cGlycan-Pathways%7c)&orgids=(IN55195+I012T4+INKY1+IN366+IN370)) | [0](http://localhost:1555/comp-genomics?type=(PWY-CLASS-DIST+%7cGlycan-Pathways%7c)&orgids=IN55195) | [0](http://localhost:1555/comp-genomics?type=(PWY-CLASS-DIST+%7cGlycan-Pathways%7c)&orgids=I012T4) | [0](http://localhost:1555/comp-genomics?type=(PWY-CLASS-DIST+%7cGlycan-Pathways%7c)&orgids=INKY1) | [0](http://localhost:1555/comp-genomics?type=(PWY-CLASS-DIST+%7cGlycan-Pathways%7c)&orgids=IN366) | [0](http://localhost:1555/comp-genomics?type=(PWY-CLASS-DIST+%7cGlycan-Pathways%7c)&orgids=IN370) |
| [Signal transduction pathways](http://localhost:1555/comp-genomics?type=(PWY-CLASS-DIST+%7cSignaling-Pathways%7c)&orgids=(IN55195+I012T4+INKY1+IN366+IN370)) | [0](http://localhost:1555/comp-genomics?type=(PWY-CLASS-DIST+%7cSignaling-Pathways%7c)&orgids=IN55195) | [0](http://localhost:1555/comp-genomics?type=(PWY-CLASS-DIST+%7cSignaling-Pathways%7c)&orgids=I012T4) | [0](http://localhost:1555/comp-genomics?type=(PWY-CLASS-DIST+%7cSignaling-Pathways%7c)&orgids=INKY1) | [0](http://localhost:1555/comp-genomics?type=(PWY-CLASS-DIST+%7cSignaling-Pathways%7c)&orgids=IN366) | [0](http://localhost:1555/comp-genomics?type=(PWY-CLASS-DIST+%7cSignaling-Pathways%7c)&orgids=IN370) |
| [Total](http://localhost:1555/comp-genomics?type=(PWY-CLASS-DIST+TOTAL)&orgids=(IN55195+I012T4+INKY1+IN366+IN370)) | 114 | 110 | 109 | 117 | 106 |

**Supplemental Table 5H: Shared Pathways**

This table counts the pathways that are shared between pairs of organisms.The number in parentheses is for the pairwise pathways comparison between two organisms - the Jaccard similarity coefficient for the pathways.

Click on the first cell (Pathways Shared by Organism Pairs) to see a table listing all shared pathways.
Click on a number within a cell to see a listing of those shared pathways.

| [**Pathways Shared by Organism Pairs**](http://localhost:1555/comp-genomics?type=PATHWAY-SHARED&orgids=(IN55195+I012T4+INKY1+IN366+IN370)) | [*L.iners* ATCC 55195](http://localhost:1555/IN55195/organism-summary) | [*L.iners* I012T4](http://localhost:1555/I012T4/organism-summary) | [*L.iners* KY](http://localhost:1555/INKY1/organism-summary) | [*L.iners* pt1](http://localhost:1555/IN366/organism-summary) | [*L.iners* pt2](http://localhost:1555/IN370/organism-summary) |
| --- | --- | --- | --- | --- | --- |
| [*Lactobacillus iners* ATCC 55195](http://localhost:1555/comp-genomics?type=(PATHWAY-SHARED+IN55195)&orgids=(IN55195+I012T4+INKY1+IN366+IN370)) | [104 (1.000)](http://localhost:1555/comp-genomics?type=(PATHWAY-SHARED+IN55195)&orgids=IN55195) | [94 (0.855)](http://localhost:1555/comp-genomics?type=(PATHWAY-SHARED+IN55195)&orgids=(IN55195+I012T4)) | [94 (0.862)](http://localhost:1555/comp-genomics?type=(PATHWAY-SHARED+IN55195)&orgids=(IN55195+INKY1)) | [104 (0.972)](http://localhost:1555/comp-genomics?type=(PATHWAY-SHARED+IN55195)&orgids=(IN55195+IN366)) | [93 (0.869)](http://localhost:1555/comp-genomics?type=(PATHWAY-SHARED+IN55195)&orgids=(IN55195+IN370)) |
| [*Lactobacillus iners* I012T4](http://localhost:1555/comp-genomics?type=(PATHWAY-SHARED+I012T4)&orgids=(IN55195+I012T4+INKY1+IN366+IN370)) | [94 (0.855)](http://localhost:1555/comp-genomics?type=(PATHWAY-SHARED+I012T4)&orgids=(I012T4+IN55195)) | [100 (1.000)](http://localhost:1555/comp-genomics?type=(PATHWAY-SHARED+I012T4)&orgids=I012T4) | [95 (0.913)](http://localhost:1555/comp-genomics?type=(PATHWAY-SHARED+I012T4)&orgids=(I012T4+INKY1)) | [94 (0.832)](http://localhost:1555/comp-genomics?type=(PATHWAY-SHARED+I012T4)&orgids=(I012T4+IN366)) | [86 (0.782)](http://localhost:1555/comp-genomics?type=(PATHWAY-SHARED+I012T4)&orgids=(I012T4+IN370)) |
| [*Lactobacillus iners KY*](http://localhost:1555/comp-genomics?type=(PATHWAY-SHARED+INKY1)&orgids=(IN55195+I012T4+INKY1+IN366+IN370)) | [94 (0.862)](http://localhost:1555/comp-genomics?type=(PATHWAY-SHARED+INKY1)&orgids=(INKY1+IN55195)) | [95 (0.913)](http://localhost:1555/comp-genomics?type=(PATHWAY-SHARED+INKY1)&orgids=(INKY1+I012T4)) | [99 (1.000)](http://localhost:1555/comp-genomics?type=(PATHWAY-SHARED+INKY1)&orgids=INKY1) | [94 (0.839)](http://localhost:1555/comp-genomics?type=(PATHWAY-SHARED+INKY1)&orgids=(INKY1+IN366)) | [85 (0.773)](http://localhost:1555/comp-genomics?type=(PATHWAY-SHARED+INKY1)&orgids=(INKY1+IN370)) |
| [*Lactobacillus iners* pt1](http://localhost:1555/comp-genomics?type=(PATHWAY-SHARED+IN366)&orgids=(IN55195+I012T4+INKY1+IN366+IN370)) | [104 (0.972)](http://localhost:1555/comp-genomics?type=(PATHWAY-SHARED+IN366)&orgids=(IN366+IN55195)) | [94 (0.832)](http://localhost:1555/comp-genomics?type=(PATHWAY-SHARED+IN366)&orgids=(IN366+I012T4)) | [94 (0.839)](http://localhost:1555/comp-genomics?type=(PATHWAY-SHARED+IN366)&orgids=(IN366+INKY1)) | [107 (1.000)](http://localhost:1555/comp-genomics?type=(PATHWAY-SHARED+IN366)&orgids=IN366) | [94 (0.862)](http://localhost:1555/comp-genomics?type=(PATHWAY-SHARED+IN366)&orgids=(IN366+IN370)) |
| [*Lactobacillus iners* pt2](http://localhost:1555/comp-genomics?type=(PATHWAY-SHARED+IN370)&orgids=(IN55195+I012T4+INKY1+IN366+IN370)) | [93 (0.869)](http://localhost:1555/comp-genomics?type=(PATHWAY-SHARED+IN370)&orgids=(IN370+IN55195)) | [86 (0.782)](http://localhost:1555/comp-genomics?type=(PATHWAY-SHARED+IN370)&orgids=(IN370+I012T4)) | [85 (0.773)](http://localhost:1555/comp-genomics?type=(PATHWAY-SHARED+IN370)&orgids=(IN370+INKY1)) | [94 (0.862)](http://localhost:1555/comp-genomics?type=(PATHWAY-SHARED+IN370)&orgids=(IN370+IN366)) | [96 (1.000)](http://localhost:1555/comp-genomics?type=(PATHWAY-SHARED+IN370)&orgids=IN370) |

**Supplemental Table 5I: Unique Pathways**

| Unique Pathways in Organism: Unique Pathways | L.iners ATCC 55195 | L.iners I012T4 | L.iners KY | L.iners pt1 | L.iners pt2 |
| --- | --- | --- | --- | --- | --- |
| [glycerol degradation I](http://localhost:1555/compare-frame-in-orgs?type=NIL&object=PWY-4261&orgids=(IN55195+I012T4+INKY1+IN366+IN370)) |  |  |  | [✓](http://localhost:1555/IN366/NEW-IMAGE?type=PATHWAY&object=PWY-4261&orgids=(IN55195%20I012T4%20INKY1%20IN366%20IN370)) |  |
| [glycerol-3-phosphate to fumarate electron transfer](http://localhost:1555/compare-frame-in-orgs?type=NIL&object=PWY0-1582&orgids=(IN55195+I012T4+INKY1+IN366+IN370)) |  |  |  | [✓](http://localhost:1555/IN366/NEW-IMAGE?type=PATHWAY&object=PWY0-1582&orgids=(IN55195%20I012T4%20INKY1%20IN366%20IN370)) |  |
| [tRNA-uridine 2-thiolation and selenation (bacteria)](http://localhost:1555/compare-frame-in-orgs?type=NIL&object=PWY-7892&orgids=(IN55195+I012T4+INKY1+IN366+IN370)) |  | [✓](http://localhost:1555/I012T4/NEW-IMAGE?type=PATHWAY&object=PWY-7892&orgids=(IN55195%20I012T4%20INKY1%20IN366%20IN370)) |  |  |  |
| [Total](http://localhost:1555/comp-genomics?type=(PATHWAY-UNIQUE+UNIQUE-PATHWAYS)&orgids=(IN55195+I012T4+INKY1+IN366+IN370)) | [0](http://localhost:1555/comp-genomics?type=(PATHWAY-UNIQUE+UNIQUE-PATHWAYS)&orgids=IN55195) | [1](http://localhost:1555/comp-genomics?type=(PATHWAY-UNIQUE+UNIQUE-PATHWAYS)&orgids=I012T4) | [0](http://localhost:1555/comp-genomics?type=(PATHWAY-UNIQUE+UNIQUE-PATHWAYS)&orgids=INKY1) | [2](http://localhost:1555/comp-genomics?type=(PATHWAY-UNIQUE+UNIQUE-PATHWAYS)&orgids=IN366) | [0](http://localhost:1555/comp-genomics?type=(PATHWAY-UNIQUE+UNIQUE-PATHWAYS)&orgids=IN370) |

**Supplemental Table 5J: Pathway Holes**

A pathway hole is a reaction in a pathway for which no corresponding gene has been identified in the genome. Pathway holes may exist for a number of possible reasons: They may represent true enzymatic functions in the organism for which the gene has not yet been found, or they could represent false positive pathway predictions or cases in which the pathway in this organism differs slightly from the reference pathway in MetaCyc. This table counts all the pathway holes in each organism database, and classifies pathways based on their number of pathway holes.

| [**Pathway Holes**](http://localhost:1555/comp-genomics?type=PATHWAY-HOLES&orgids=(IN55195+I012T4+INKY1+IN366+IN370)) | [*L.iners* ATCC 55195](http://localhost:1555/IN55195/organism-summary) | [*L.iners* I012T4](http://localhost:1555/I012T4/organism-summary) | [*L.iners* KY](http://localhost:1555/INKY1/organism-summary) | [*L.iners* pt1](http://localhost:1555/IN366/organism-summary) | [*L.iners* pt2](http://localhost:1555/IN370/organism-summary) |
| --- | --- | --- | --- | --- | --- |
| [Number of Pathway Holes](http://localhost:1555/comp-genomics?type=(PATHWAY-HOLES+NUM-HOLES)&orgids=(IN55195+I012T4+INKY1+IN366+IN370)) | [98](http://localhost:1555/comp-genomics?type=(PATHWAY-HOLES+NUM-HOLES)&orgids=IN55195) | [103](http://localhost:1555/comp-genomics?type=(PATHWAY-HOLES+NUM-HOLES)&orgids=I012T4) | [93](http://localhost:1555/comp-genomics?type=(PATHWAY-HOLES+NUM-HOLES)&orgids=INKY1) | [97](http://localhost:1555/comp-genomics?type=(PATHWAY-HOLES+NUM-HOLES)&orgids=IN366) | [81](http://localhost:1555/comp-genomics?type=(PATHWAY-HOLES+NUM-HOLES)&orgids=IN370) |
| [Pathway Holes as a percentage of total reactions in pathways](http://localhost:1555/comp-genomics?type=(PATHWAY-HOLES+PERCENT-HOLES)&orgids=(IN55195+I012T4+INKY1+IN366+IN370)) | 33% | 35% | 34% | 32% | 31% |
| [Pathways with No Holes](http://localhost:1555/comp-genomics?type=(PATHWAY-HOLES+0)&orgids=(IN55195+I012T4+INKY1+IN366+IN370)) | [48](http://localhost:1555/comp-genomics?type=(PATHWAY-HOLES+0)&orgids=IN55195) | [48](http://localhost:1555/comp-genomics?type=(PATHWAY-HOLES+0)&orgids=I012T4) | [46](http://localhost:1555/comp-genomics?type=(PATHWAY-HOLES+0)&orgids=INKY1) | [51](http://localhost:1555/comp-genomics?type=(PATHWAY-HOLES+0)&orgids=IN366) | [47](http://localhost:1555/comp-genomics?type=(PATHWAY-HOLES+0)&orgids=IN370) |
| [Pathways with 1 Hole](http://localhost:1555/comp-genomics?type=(PATHWAY-HOLES+1)&orgids=(IN55195+I012T4+INKY1+IN366+IN370)) | [28](http://localhost:1555/comp-genomics?type=(PATHWAY-HOLES+1)&orgids=IN55195) | [23](http://localhost:1555/comp-genomics?type=(PATHWAY-HOLES+1)&orgids=I012T4) | [27](http://localhost:1555/comp-genomics?type=(PATHWAY-HOLES+1)&orgids=INKY1) | [29](http://localhost:1555/comp-genomics?type=(PATHWAY-HOLES+1)&orgids=IN366) | [27](http://localhost:1555/comp-genomics?type=(PATHWAY-HOLES+1)&orgids=IN370) |
| [Pathways with 2 Holes](http://localhost:1555/comp-genomics?type=(PATHWAY-HOLES+2)&orgids=(IN55195+I012T4+INKY1+IN366+IN370)) | [15](http://localhost:1555/comp-genomics?type=(PATHWAY-HOLES+2)&orgids=IN55195) | [16](http://localhost:1555/comp-genomics?type=(PATHWAY-HOLES+2)&orgids=I012T4) | [15](http://localhost:1555/comp-genomics?type=(PATHWAY-HOLES+2)&orgids=INKY1) | [14](http://localhost:1555/comp-genomics?type=(PATHWAY-HOLES+2)&orgids=IN366) | [8](http://localhost:1555/comp-genomics?type=(PATHWAY-HOLES+2)&orgids=IN370) |
| [Pathways with 3 Holes](http://localhost:1555/comp-genomics?type=(PATHWAY-HOLES+3)&orgids=(IN55195+I012T4+INKY1+IN366+IN370)) | [4](http://localhost:1555/comp-genomics?type=(PATHWAY-HOLES+3)&orgids=IN55195) | [2](http://localhost:1555/comp-genomics?type=(PATHWAY-HOLES+3)&orgids=I012T4) | [2](http://localhost:1555/comp-genomics?type=(PATHWAY-HOLES+3)&orgids=INKY1) | [4](http://localhost:1555/comp-genomics?type=(PATHWAY-HOLES+3)&orgids=IN366) | [4](http://localhost:1555/comp-genomics?type=(PATHWAY-HOLES+3)&orgids=IN370) |
| [Pathways with 4 Holes](http://localhost:1555/comp-genomics?type=(PATHWAY-HOLES+4)&orgids=(IN55195+I012T4+INKY1+IN366+IN370)) | [7](http://localhost:1555/comp-genomics?type=(PATHWAY-HOLES+4)&orgids=IN55195) | [8](http://localhost:1555/comp-genomics?type=(PATHWAY-HOLES+4)&orgids=I012T4) | [7](http://localhost:1555/comp-genomics?type=(PATHWAY-HOLES+4)&orgids=INKY1) | [7](http://localhost:1555/comp-genomics?type=(PATHWAY-HOLES+4)&orgids=IN366) | [9](http://localhost:1555/comp-genomics?type=(PATHWAY-HOLES+4)&orgids=IN370) |
| [Pathways with 5 Holes](http://localhost:1555/comp-genomics?type=(PATHWAY-HOLES+5)&orgids=(IN55195+I012T4+INKY1+IN366+IN370)) | [0](http://localhost:1555/comp-genomics?type=(PATHWAY-HOLES+5)&orgids=IN55195) | [0](http://localhost:1555/comp-genomics?type=(PATHWAY-HOLES+5)&orgids=I012T4) | [0](http://localhost:1555/comp-genomics?type=(PATHWAY-HOLES+5)&orgids=INKY1) | [0](http://localhost:1555/comp-genomics?type=(PATHWAY-HOLES+5)&orgids=IN366) | [0](http://localhost:1555/comp-genomics?type=(PATHWAY-HOLES+5)&orgids=IN370) |
| [Pathways with > 5 Holes](http://localhost:1555/comp-genomics?type=(PATHWAY-HOLES+MORE-THAN-5)&orgids=(IN55195+I012T4+INKY1+IN366+IN370)) | [2](http://localhost:1555/comp-genomics?type=(PATHWAY-HOLES+MORE-THAN-5)&orgids=IN55195) | [3](http://localhost:1555/comp-genomics?type=(PATHWAY-HOLES+MORE-THAN-5)&orgids=I012T4) | [2](http://localhost:1555/comp-genomics?type=(PATHWAY-HOLES+MORE-THAN-5)&orgids=INKY1) | [2](http://localhost:1555/comp-genomics?type=(PATHWAY-HOLES+MORE-THAN-5)&orgids=IN366) | [1](http://localhost:1555/comp-genomics?type=(PATHWAY-HOLES+MORE-THAN-5)&orgids=IN370) |
| [Total Pathways with Holes](http://localhost:1555/comp-genomics?type=(PATHWAY-HOLES+TOTAL)&orgids=(IN55195+I012T4+INKY1+IN366+IN370)) | [56](http://localhost:1555/comp-genomics?type=(PATHWAY-HOLES+TOTAL)&orgids=IN55195) | [52](http://localhost:1555/comp-genomics?type=(PATHWAY-HOLES+TOTAL)&orgids=I012T4) | [53](http://localhost:1555/comp-genomics?type=(PATHWAY-HOLES+TOTAL)&orgids=INKY1) | [56](http://localhost:1555/comp-genomics?type=(PATHWAY-HOLES+TOTAL)&orgids=IN366) | [49](http://localhost:1555/comp-genomics?type=(PATHWAY-HOLES+TOTAL)&orgids=IN370) |

**Supplemental Table 5K. Pathways involved in degradation of carbohydrate in studied *L. iners* strains.**

*L.iners* ATCC 55195 is a draft genome of *L.iners* ATCC 55195 strain isolated form a patient with bacterial vaginosis

*L.iners* I012T4 is the high quality draft genome of *L.iners* assembled from WGS sequencing data of a cervical swab of CC patient.

*L.iners* KY is the complete genome of *L.iners* isolated from a healthy individual

*L.iners* pt1 and *L.iners* pt2 are draft genomes of *L.iners*  isolated from cervical swabs of 2 CC patients.

The table was generated by Pathway Tools software.

In addition to reflecting differences in biology of different organisms, these statistics reflect completeness of the PGDBs for the strains, which is lowest in *L. iners* (91% versus 97%-100% in the rest).

| **Pathway Class**: Degradation/Utilization/Assimilation - Carbohydrate Degradation | *L.iners* ATCC 55195 | *L.iners* I012T4 | *L.iners* KY | *L.iners* pt1 | *L.iners* pt2 |
| --- | --- | --- | --- | --- | --- |
| [D-galactose degradation I (Leloir pathway)](http://localhost:1555/compare-frame-in-orgs?type=PATHWAY&object=PWY-6317&orgids=(IN55195+I012T4+INKY1+IN366+IN370)) | [✓](http://localhost:1555/IN55195/NEW-IMAGE?type=PATHWAY&object=PWY-6317&orgids=(IN55195%20I012T4%20INKY1%20IN366%20IN370)) | [✓](http://localhost:1555/I012T4/NEW-IMAGE?type=PATHWAY&object=PWY-6317&orgids=(IN55195%20I012T4%20INKY1%20IN366%20IN370)) | [✓](http://localhost:1555/INKY1/NEW-IMAGE?type=PATHWAY&object=PWY-6317&orgids=(IN55195%20I012T4%20INKY1%20IN366%20IN370)) | [✓](http://localhost:1555/IN366/NEW-IMAGE?type=PATHWAY&object=PWY-6317&orgids=(IN55195%20I012T4%20INKY1%20IN366%20IN370)) | [✓](http://localhost:1555/IN370/NEW-IMAGE?type=PATHWAY&object=PWY-6317&orgids=(IN55195%20I012T4%20INKY1%20IN366%20IN370)) |
| [D-mannose degradation I](http://localhost:1555/compare-frame-in-orgs?type=PATHWAY&object=MANNCAT-PWY&orgids=(IN55195+I012T4+INKY1+IN366+IN370)) | [✓](http://localhost:1555/IN55195/NEW-IMAGE?type=PATHWAY&object=MANNCAT-PWY&orgids=(IN55195%20I012T4%20INKY1%20IN366%20IN370)) | [✓](http://localhost:1555/I012T4/NEW-IMAGE?type=PATHWAY&object=MANNCAT-PWY&orgids=(IN55195%20I012T4%20INKY1%20IN366%20IN370)) | [✓](http://localhost:1555/INKY1/NEW-IMAGE?type=PATHWAY&object=MANNCAT-PWY&orgids=(IN55195%20I012T4%20INKY1%20IN366%20IN370)) | [✓](http://localhost:1555/IN366/NEW-IMAGE?type=PATHWAY&object=MANNCAT-PWY&orgids=(IN55195%20I012T4%20INKY1%20IN366%20IN370)) | [✓](http://localhost:1555/IN370/NEW-IMAGE?type=PATHWAY&object=MANNCAT-PWY&orgids=(IN55195%20I012T4%20INKY1%20IN366%20IN370)) |
| [fructose degradation](http://localhost:1555/compare-frame-in-orgs?type=PATHWAY&object=PWY0-1314&orgids=(IN55195+I012T4+INKY1+IN366+IN370)) | [✓](http://localhost:1555/IN55195/NEW-IMAGE?type=PATHWAY&object=PWY0-1314&orgids=(IN55195%20I012T4%20INKY1%20IN366%20IN370)) | [✓](http://localhost:1555/I012T4/NEW-IMAGE?type=PATHWAY&object=PWY0-1314&orgids=(IN55195%20I012T4%20INKY1%20IN366%20IN370)) | [✓](http://localhost:1555/INKY1/NEW-IMAGE?type=PATHWAY&object=PWY0-1314&orgids=(IN55195%20I012T4%20INKY1%20IN366%20IN370)) | [✓](http://localhost:1555/IN366/NEW-IMAGE?type=PATHWAY&object=PWY0-1314&orgids=(IN55195%20I012T4%20INKY1%20IN366%20IN370)) | [✓](http://localhost:1555/IN370/NEW-IMAGE?type=PATHWAY&object=PWY0-1314&orgids=(IN55195%20I012T4%20INKY1%20IN366%20IN370)) |
| [lactose and galactose degradation I](http://localhost:1555/compare-frame-in-orgs?type=PATHWAY&object=LACTOSECAT-PWY&orgids=(IN55195+I012T4+INKY1+IN366+IN370)) | [✓](http://localhost:1555/IN55195/NEW-IMAGE?type=PATHWAY&object=LACTOSECAT-PWY&orgids=(IN55195%20I012T4%20INKY1%20IN366%20IN370)) | [✓](http://localhost:1555/I012T4/NEW-IMAGE?type=PATHWAY&object=LACTOSECAT-PWY&orgids=(IN55195%20I012T4%20INKY1%20IN366%20IN370)) |  | [✓](http://localhost:1555/IN366/NEW-IMAGE?type=PATHWAY&object=LACTOSECAT-PWY&orgids=(IN55195%20I012T4%20INKY1%20IN366%20IN370)) |  |
| [maltose degradation](http://localhost:1555/compare-frame-in-orgs?type=PATHWAY&object=MALTOSECAT-PWY&orgids=(IN55195+I012T4+INKY1+IN366+IN370)) | [✓](http://localhost:1555/IN55195/NEW-IMAGE?type=PATHWAY&object=MALTOSECAT-PWY&orgids=(IN55195%20I012T4%20INKY1%20IN366%20IN370)) |  | [✓](http://localhost:1555/INKY1/NEW-IMAGE?type=PATHWAY&object=MALTOSECAT-PWY&orgids=(IN55195%20I012T4%20INKY1%20IN366%20IN370)) | [✓](http://localhost:1555/IN366/NEW-IMAGE?type=PATHWAY&object=MALTOSECAT-PWY&orgids=(IN55195%20I012T4%20INKY1%20IN366%20IN370)) | [✓](http://localhost:1555/IN370/NEW-IMAGE?type=PATHWAY&object=MALTOSECAT-PWY&orgids=(IN55195%20I012T4%20INKY1%20IN366%20IN370)) |

**Supplemental Table 5L. Cross-Species Comparison of lactose and galactose degradation I.**

The pathway was not predicted in *L.iners* KY and *L.iners* pt2. Only 1 of 2 enzymes, which are unique for the pathway was identified in [*L. iners* ATCC 55195](http://localhost:1555/IN55195/NEW-IMAGE?type=PATHWAY&object=LACTOSECAT-PWY&orgids=(IN55195%20I012T4%20INKY1%20IN366%20IN370)) strain.

|  | [L.iners ATCC 55195](http://localhost:1555/IN55195/NEW-IMAGE?type=PATHWAY&object=LACTOSECAT-PWY&orgids=(IN55195%20I012T4%20INKY1%20IN366%20IN370)) | [L.iners I012T4](http://localhost:1555/I012T4/NEW-IMAGE?type=PATHWAY&object=LACTOSECAT-PWY&orgids=(IN55195%20I012T4%20INKY1%20IN366%20IN370)) |  | [L.iners pt1](http://localhost:1555/IN366/NEW-IMAGE?type=PATHWAY&object=LACTOSECAT-PWY&orgids=(IN55195%20I012T4%20INKY1%20IN366%20IN370)) |
| --- | --- | --- | --- | --- |
| Pathway Evidence Glyph.  Reactions are labelled by green lines if enzyme was identified or black if not identified in the organism. Orange line labels enzymes that are iunique for the pathway | 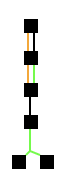 | 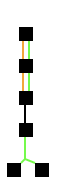 |  | 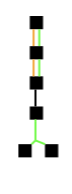 |
| Operons  encoding enzymes of the pathway | 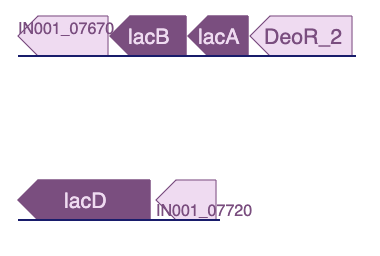 | 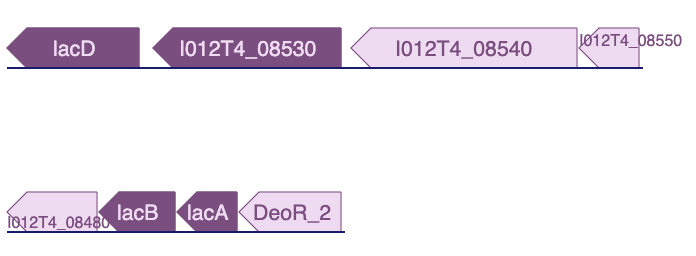 |  | 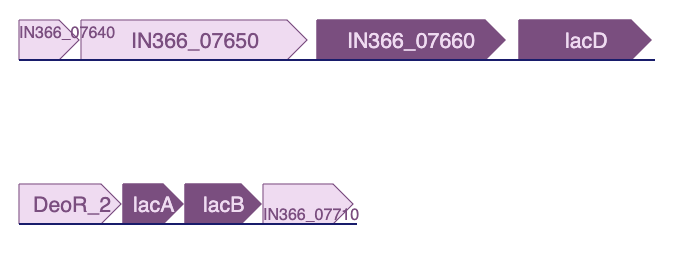 |
| [LACTOSE6P-HYDROXY-RXN](http://localhost:1555/META/NEW-IMAGE?type=REACTION&object=LACTOSE6P-HYDROXY-RXN&orgids=(IN55195%20I012T4%20INKY1%20IN366%20IN370)) | None | [6-phospho-beta-galactosidase](http://localhost:1555/gene?orgid=I012T4&id=I012T4_08530-MONOMER): [I012T4_08530](http://localhost:1555/gene?orgid=I012T4&id=I012T4_08530) |  | [6-phospho-beta-galactosidase](http://localhost:1555/gene?orgid=IN366&id=IN366_07660-MONOMER): [IN366_07660](http://localhost:1555/gene?orgid=IN366&id=IN366_07660) |
| [LACTOSE-6-PHOSPHATE-ISOMERASE-RXN](http://localhost:1555/META/NEW-IMAGE?type=REACTION&object=LACTOSE-6-PHOSPHATE-ISOMERASE-RXN&orgids=(IN55195%20I012T4%20INKY1%20IN366%20IN370)) | [galactose-6-phosphate isomerase subunit LacB](http://localhost:1555/gene?orgid=IN55195&id=IN001_07680-MONOMER): [lacB](http://localhost:1555/gene?orgid=IN55195&id=IN001_07680) [galactose-6-phosphate isomerase subunit LacA](http://localhost:1555/gene?orgid=IN55195&id=IN001_07690-MONOMER): [lacA](http://localhost:1555/gene?orgid=IN55195&id=IN001_07690) | [galactose-6-phosphate isomerase](http://localhost:1555/I012T4/NEW-IMAGE?type=ENZYME&object=CPLX1-12&orgids=(IN55195%20I012T4%20INKY1%20IN366%20IN370)): [lacB](http://localhost:1555/gene?orgid=I012T4&id=I012T4_08490), [lacA](http://localhost:1555/gene?orgid=I012T4&id=I012T4_08500) |  | [galactose-6-phosphate isomerase subunit LacB](http://localhost:1555/gene?orgid=IN366&id=IN366_07700-MONOMER): [lacB](http://localhost:1555/gene?orgid=IN366&id=IN366_07700) [galactose-6-phosphate isomerase subunit LacA](http://localhost:1555/gene?orgid=IN366&id=IN366_07690-MONOMER): [lacA](http://localhost:1555/gene?orgid=IN366&id=IN366_07690) |
| [EC 2.7.1.144](http://localhost:1555/META/NEW-IMAGE?type=REACTION&object=TAGAKIN-RXN&orgids=(IN55195%20I012T4%20INKY1%20IN366%20IN370)) | None | None |  | None |
| EC 4.1.2.40 | [tagatose-bisphosphate aldolase](http://localhost:1555/gene?orgid=IN55195&id=IN001_07710-MONOMER): [lacD](http://localhost:1555/gene?orgid=IN55195&id=IN001_07710) | [tagatose-bisphosphate aldolase](http://localhost:1555/gene?orgid=I012T4&id=I012T4_08520-MONOMER): [lacD](http://localhost:1555/gene?orgid=I012T4&id=I012T4_08520) |  | [tagatose-bisphosphate aldolase](http://localhost:1555/gene?orgid=IN366&id=IN366_07670-MONOMER): [lacD](http://localhost:1555/gene?orgid=IN366&id=IN366_07670) |
